# Frequency of neurological manifestations in COVID-19: a systematic review and meta-analysis of 350 studies

**DOI:** 10.1101/2021.04.20.21255780

**Authors:** Shubham Misra, Kavitha Kolappa, Manya Prasad, Divya Radhakrishnan, Kiran T Thakur, Tom Solomon, Benedict Daniel Michael, Andrea Sylvia Winkler, Ettore Beghi, Alla Guekht, Carlos A. Pardo, Greta Karen Wood, Sherry Hsiang-Yi Chou, Ericka L. Fink, Erich Schmutzhard, Amir Kheradmand, Fan Kee Hoo, Amit Kumar, Animesh Das, Achal K Srivastava, Ayush Agarwal, Tarun Dua, Kameshwar Prasad

## Abstract

**Objective:** To summarize the frequency of neurological manifestations reported in COVID-19 patients and investigate the association of these manifestations with disease severity and mortality.

**Design:** Systematic review and meta-analysis

**Eligibility criteria:** Studies enrolling consecutive COVID-19 patients (probable or confirmed) presenting with neurological manifestations.

**Data sources:** PubMed, Medline, Cochrane library, clinicaltrials.gov and EMBASE from 31^st^ December 2019 to 15^th^ December 2020.

**Data extraction and analysis:** Two authors independently screened titles and abstracts retrieved by literature search. Risk of bias was examined using Joanna Briggs Institute (JBI) scale. A random-effects meta-analysis was performed, and pooled prevalence and 95% Confidence Intervals (CI) were calculated for neurological manifestations. Odds ratio (OR) and 95%CI were calculated to determine the association of neurological manifestations with disease severity and mortality. Presence of heterogeneity was assessed using I-square, meta-regression, and subgroup analyses. Statistical analyses were conducted in R version 3.6.2.

**Results:** Of 2,455 citations, 350 studies were included in this review, providing data on 145,634 COVID-19 patients, 89% of whom were hospitalized. Forty-one neurological manifestations (24 symptoms and 17 diagnoses) were identified. Pooled prevalence of the most common neurological symptoms included: fatigue (32%), myalgia (20%), taste impairment (21%), smell impairment (19%) and headache (13%). A low risk of bias was observed in 85% of studies; studies with higher risk of bias yielded higher prevalence estimates. Stroke was the most common neurological diagnosis (pooled prevalence-2%). In COVID-19 patients aged >60, the pooled prevalence of acute confusion/delirium was 34% and the presence of any neurological manifestations in this age group was associated with mortality (OR 1.80; 95%CI 1.11 to 2.91).

**Conclusions:** Up to one-third of COVID-19 patients analysed in this review experienced at least one neurological manifestation. One in 50 patients experienced stroke. In those over 60, more than one-third had acute confusion/delirium; the presence of neurological manifestations in this group was associated with near doubling of mortality. Results must be interpreted keeping in view the limitations of observational studies and associated bias.

**Systematic review registration:** PROSPERO CRD42020181867.

**What is already known on this topic:** The frequency of neurological manifestations including fatigue, myalgia, taste and smell impairments, headache and dizziness in COVID-19 patients has been reported in a few systematic reviews and meta-analyses. However, considerable heterogeneity has been observed in terms of methodological quality of the studies, severity of the disease, mean age and hospitalization status of the patients. The evidence regarding the frequency of neurological diagnoses including stroke, encephalitis, Guillain Barré syndrome (GBS) is also limited to case reports and case series and no data exists thus far on the pooled prevalence estimates for neurological diagnoses in COVID-19 patients.

**What this study adds:** To the best of the authors’ knowledge, this is the largest systematic review and meta-analysis to date (including 350 studies with data on 145,634 cases) summarizing the evidence on the frequency of the full spectrum of neurological manifestations in COVID-19 patients in the overall, young and elderly populations. For the first time, our review reports the pooled prevalence of stroke in COVID-19 patients. Risk of bias, old age and disease severity were potential determinants of the frequency and nature of neurological manifestations as well as its association with mortality. Our review also highlights the need to develop reporting standards for studies describing the frequency of clinical features. Moreover, we note that this will be the first systematic review and meta-analysis on this subject to include studies reported in all languages.

## Introduction

One year into the COVID-19 pandemic, the disease caused by the severe acute respiratory syndrome coronavirus 2 (SARS-CoV-2) has left nearly no place on Earth untouched, with more than 137 million confirmed cases and 2.9 million deaths across 220 countries and territories to date.^1^ The disease spectrum has ranged from asymptomatic cases with or without mild respiratory symptoms to severe cases with respiratory failure and multi-organ dysfunction.^2–4^ Over the past year, reports of neurological manifestations associated with COVID-19 have increased rapidly, with the central nervous, peripheral nervous and musculoskeletal systems impacted.^5–7^ The World Health Organization (WHO) has recognized the clinical relevance of neurological manifestations of COVID-19.^8,9^

Prior systematic reviews have highlighted the frequency of common neurological symptoms observed in COVID-19 including fatigue, myalgia, impaired sense of smell or taste, and headaches.^10–12^ Additional neurological manifestations reported include dizziness, acute confusion/delirium, agitation, stroke, hypoxic ischaemic injury, seizures, coma, and encephalitis, amongst others.^11,12^ In some cases, neurological manifestations have been reported even without respiratory symptoms.^13–15^ However, considerable variations in the frequency of neurological manifestations have been reported with respect to different ethnic populations, age and hospitalization status of included patients, and severity of disease.^12^ Reports of neurological manifestations in COVID-19 patients are accumulating rapidly and this warrants updated meta-analyses with pooled prevalence estimates. Additionally, questions remain as to whether the presence of neurological manifestations are associated with increased mortality, as initial reports have suggested.^16^

We conducted a systematic review and meta-analysis to obtain pooled prevalence estimates and to understand the variations in the reported frequencies of various neurological manifestations (including symptoms and diagnoses) in COVID-19 patients across different age groups. This review updates the findings of the previously published systematic reviews on neurological symptoms, explores the reasons for heterogeneity of published studies and gives novel insights into the pooled prevalence of neurological diagnoses in COVID-19 patients. Our review further investigates the association of neurological manifestations with COVID-19 severity and mortality.

## Methodology

### Literature search

Our intention was to cover one year period to reflect on the cumulative knowledge about neurological manifestations seen in COVID-19. A comprehensive literature search was carried out for the period 31^st^ December 2019 to 15^th^ December 2020 in PubMed, EMBASE, MEDLINE, Google Scholar, Cochrane Library and ClinicalTrials.gov. The detailed search strategy is provided in the supplementary file.

### Population

The target population for this review includes patients clinically suspected to have COVID-19 confirmed by real-time reverse-transcription polymerase chain reaction (RT-PCR) detection, high throughput sequencing, SARS-CoV-2 viral culture in throat swab specimens, or SARS-CoV-2 antibody detection in blood samples. Studies had to report one or more new onset neurological manifestation(s) in a proportion of the included patients, temporally associated with SARS-CoV-2 infection and not explained by an alternate etiology. We also included studies reporting on probable cases of COVID-19, according to WHO case definitions,^17^ based on clinical presentation where diagnostic testing was not available.

### Intervention

None

### Comparator

The main objective of this review (describing the frequency of neurological manifestations) did not merit a comparator group; however, for the secondary objectives of determining whether neurological manifestations were associated with severity or mortality, the events were compared between severe and non-severe COVID-19 patients and between those with and without neurological manifestations.

### Outcomes

The primary outcome was the frequency of neurological manifestations in COVID-19 patients. Considering the lack of uniformity and reporting regarding classifications, definitions and diagnostic criteria of neurological manifestations across studies, we decided to accept study definitions as reported. For calculating pooled prevalence, both the numerator (number with neurological manifestations) and denominator (total number of patients with COVID-19 attending the facility) were required. Studies without the denominator were not included in the meta-analysis, and are listed in supplementary table 10 and table 11. Secondary outcomes included age specific variations in neurological manifestations and association of neurological manifestations with severity of and mortality in COVID-19.

### Inclusion and Exclusion Criteria

Our systematic review and meta-analysis was conducted according to the Preferred Reporting Items for Systematic Reviews and Meta-Analyses Protocol (PRISMA-P) 2015 guidelines^18^ (PRISMA checklist in supplement) and included observational studies on human subjects that: (1) included probable or confirmed COVID-19 subjects defined as above; (2) reported on neurological manifestations; (3) full texts available; and (4) published in indexed journals. There were no restrictions on language.

Studies meeting the above inclusion criteria were excluded if either of the following applied: (1) they did not specify inclusion of consecutive COVID-19 patients; (2) they were individual case-reports, or reviews, narratives or editorials. The protocol of this systematic review and meta-analysis was registered in PROSPERO (Registration ID: CRD42020181867) and there were no major deviations from the protocol.^19^

### Data extraction

Two authors independently screened titles and abstracts retrieved by the literature search against the eligibility criteria. Duplicate records were excluded. The following information was extracted from each eligible study: first author; year of publication; study design; sample size; mean or median age of subjects; comorbidities; clinical parameters; severity of COVID-19 infection; neurological symptoms and diagnoses. Hospitalization status of COVID-19 patients was extracted independently by two authors. The definitions and inter-rater agreements are given in supplementary p3, table 6 and table 7. Any disagreement between the reviewers was settled by consensus or by referring to a third author.

### Risk of bias assessment

To choose a tool for assessing risk of bias, five available tools were critically evaluated for coverage of selection and measurement bias, clarity of wording, suitability for descriptive case series, and ease of use. The Joanna Briggs Institute (JBI) tool was selected, and risk of bias assessment was conducted independently by two authors for all studies included in this review.^20^

### Statistical analysis

Age was represented by mean with standard deviation (SD) whereas sex was represented by number with percentages. Meta-analyses were performed if data on the frequency of a specified neurological manifestation were available from two or more studies. Pooled prevalence for each neurological manifestation along with its corresponding 95% confidence interval (CI) was calculated using a random-effects model. For pooling, prevalence estimates were transformed using the Freeman-Tukey double arcsine transformation for better approximation to normal distribution as required by the assumption of conventional meta-analytic model.^21^ The association between neurological symptoms and both severity of and mortality in COVID-19 were determined using the Odds Ratio (OR) and 95%CI. Subgroup analyses were also conducted to estimate the pooled prevalence of neurological symptoms and diagnoses in the elderly and young populations. The false discovery rate (FDR) was controlled using the Benjamini-Hochberg procedure. The presence of heterogeneity was assessed using I-square. Funnel plots were used to assess the risk of publication bias and the funnel plot asymmetry was assessed using the Egger’s regression test and Duval & Tweedie’s trim-and-fill test.

A meta-regression analysis was performed to further explore the source of heterogeneity for neurological symptoms and diagnoses wherein 10 or more studies were pooled. If risk of bias was found to be significantly associated with prevalence, then a subgroup analysis was conducted to stratify the outcome variables in terms of low, moderate and high risk of bias. P-values <0.05 were considered to be statistically significant. Statistical analyses were conducted in R version 3.6.2.^22^

#### Patient and Public Involvement

There was no patient or public involvement in our study.

## Results

Our search yielded 2,455 articles, of which 778 were screened by full text, and 350 met the inclusion criteria and were included in our systematic review and meta-analysis (figure 1). Of the 350 studies, 15 were included from non-English languages, for which the online English versions were retrieved. Twenty studies included only young COVID-19 patients (age <18 years) while 14 studies included only elderly patients (age> 60 years). The studies included data from 55 countries. Baseline characteristics of all the studies included in this systematic review and meta-analysis are provided in supplementary table 1 and table 2 (including countries classified by World Bank Income Category and WHO Regions).

**Figure 1:**
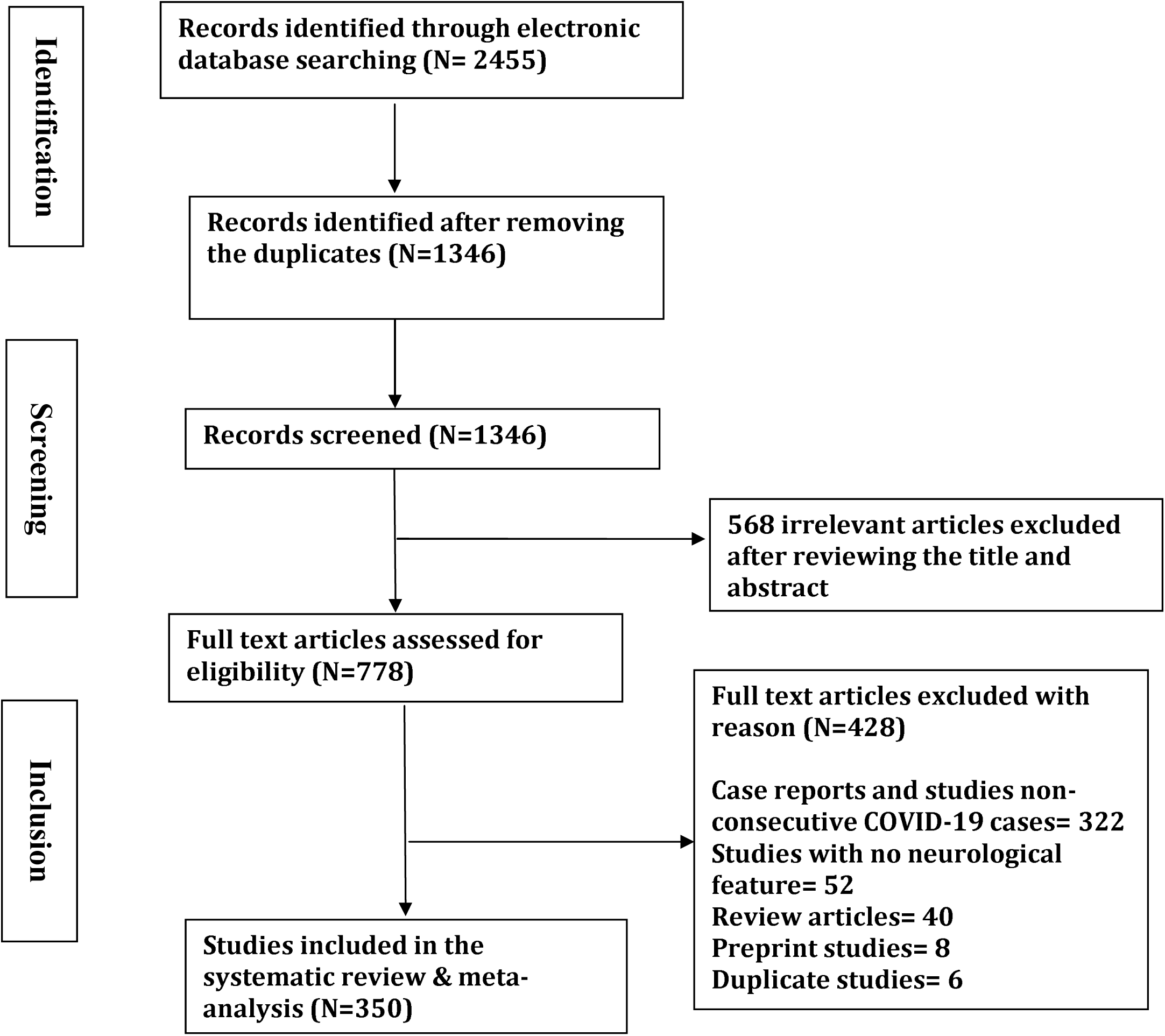
PRISMA flow diagram for the systematic review and meta-analysis

A total of 145,634 COVID-19 cases (54% males) were analysed in our systematic review. Hospitalization was reported for 129,825 cases (89%), 9,188 cases (6%) were non-hospitalized, and hospitalization status was not reported for 6,621 (5%). Pre-existing medical comorbidities were reported for hypertension 26% (95%CI 24 to 28%); diabetes 14% (95%CI 13 to 15%); cardiovascular/cerebrovascular disease (aggregated reporting) 18% (95%CI 12 to 23%); cardiovascular disease 10% (95%CI 9 to 11%); chronic obstructive pulmonary disease 5% (95%CI 4 to 6%); chronic kidney disease 5% (4-5%); cerebrovascular disease 4% (95%CI 3 to 4%); malignancy 4% (95%CI 3 to 4%) and chronic liver disease 3% (95%CI 3 to 4%).

Individual case reports and case series including neurological manifestations in non-consecutive COVID-19 cases or a specific subgroup of COVID-19 cases were present in an additional 322 studies detailed in the supplementary table 10 and table 11.

### Risk of bias assessment

Risk of bias scores utilizing the JBI tool were categorized into low (13-16), moderate (7-12) and high (0-6). Overall, 296 studies (84.6%) had a low risk of bias, 49 studies had moderate (14%) and five studies (1.4%) had a high risk of bias. Supplementary table 9 represents the risk of bias assessment done for studies included in our systematic review and meta-analysis.

### Neurological manifestations in COVID-19 patients

Twenty-four neurological symptoms from 305 articles are included in our systematic review and meta-analysis. Pooled prevalence (95%CI, Number of studies) of the most common neurological symptoms include: fatigue 32% (95%CI 30 to 35%, 169 studies); myalgia 20% (95%CI 18 to 23%, 207 studies); myalgia or fatigue 31% (95%CI 25 to 37%, 22 studies); headache 13% (95%CI 12 to 15%, 202 studies); dizziness 7% (95%CI 5 to 8%, 46 studies); headache and dizziness together 12% (95%CI 8 to 17%, nine studies); smell impairment 19% (95%CI 13 to 25%, 51 studies); taste impairment 21% (95%CI 15 to 29%, 38 studies); smell or taste impairment 18% (95%CI 10 to 28%, 14 studies); acute confusion/delirium 11% (95%CI 7 to 16%, 19 studies); disturbance of consciousness 7% (95%CI 5 to 10%, 25 studies) and agitation 45% (95%CI 3 to 93%, three studies). Pooled prevalence of neurological symptoms included in the meta-analysis are given in table 1 including the individual prevalence of neurological symptoms for which meta-analyses were not possible.

**Table 1:**
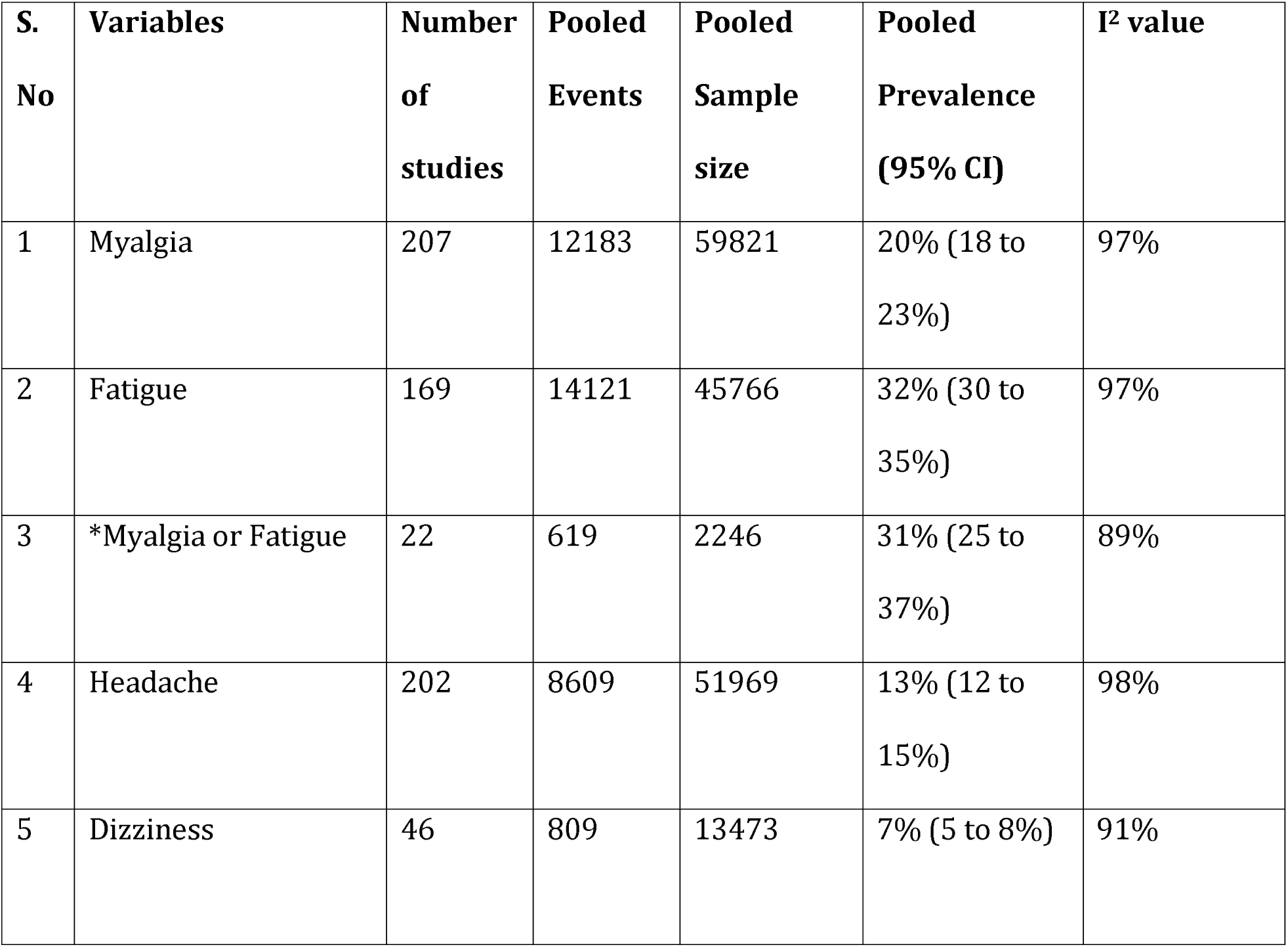
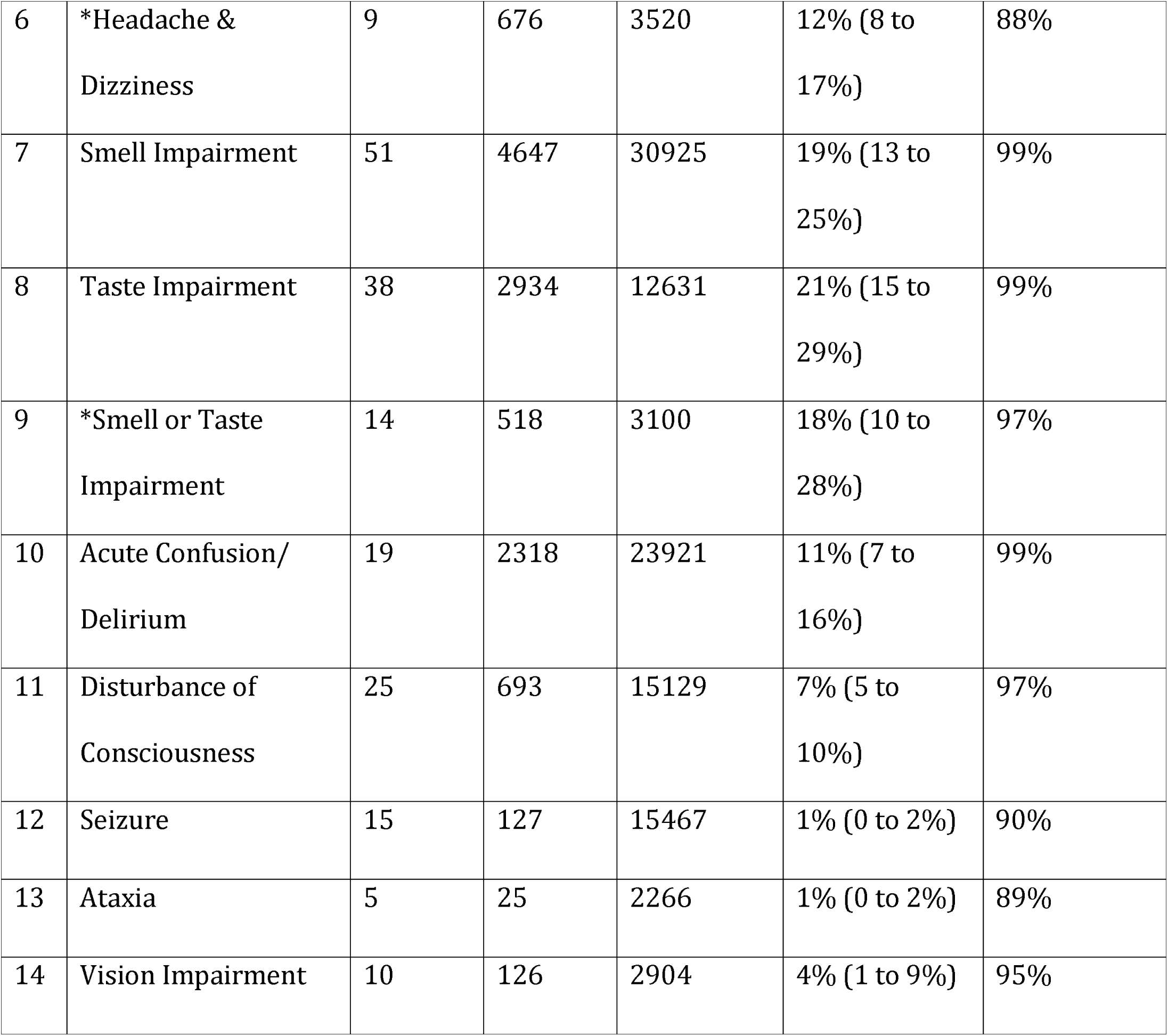
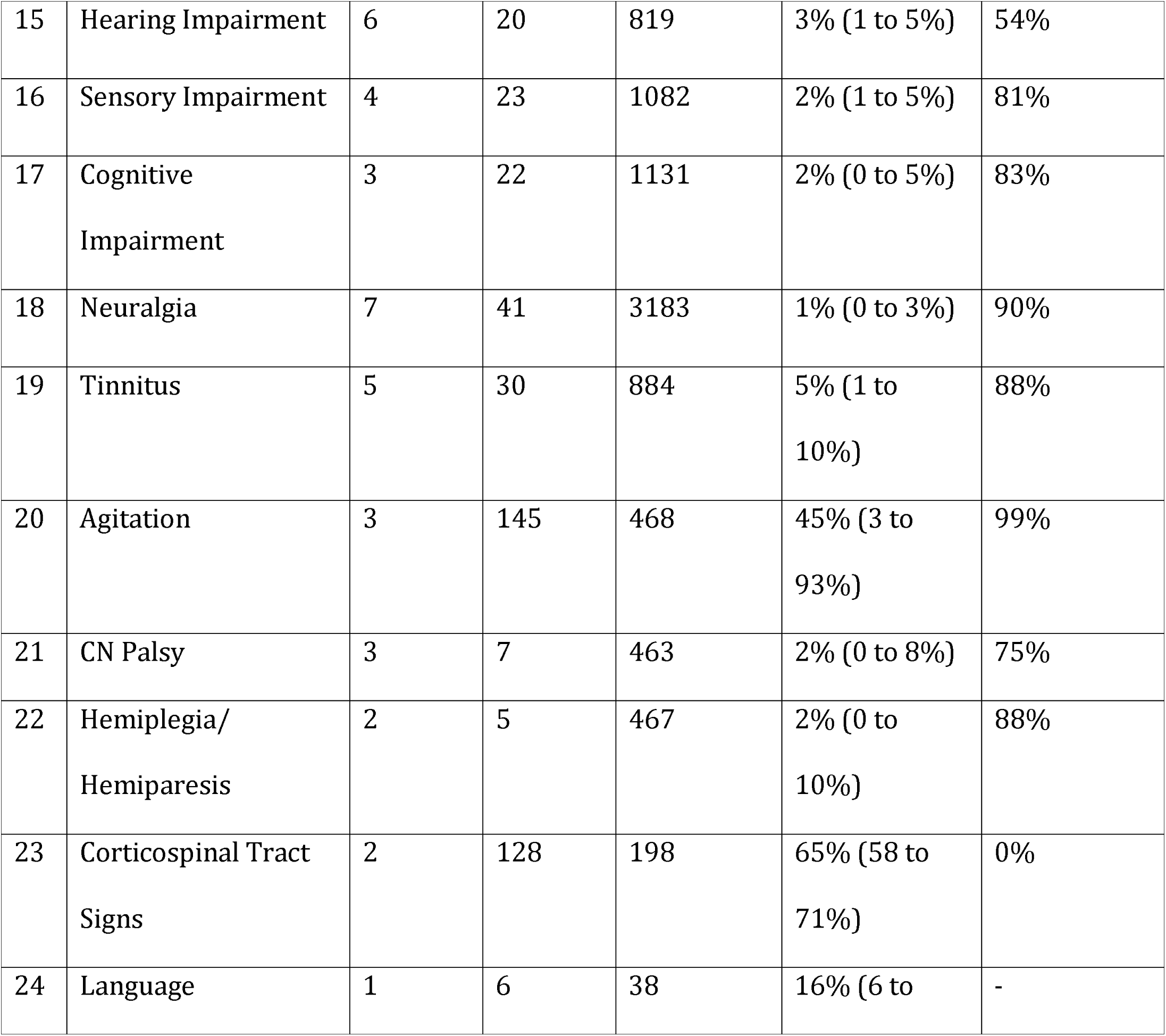
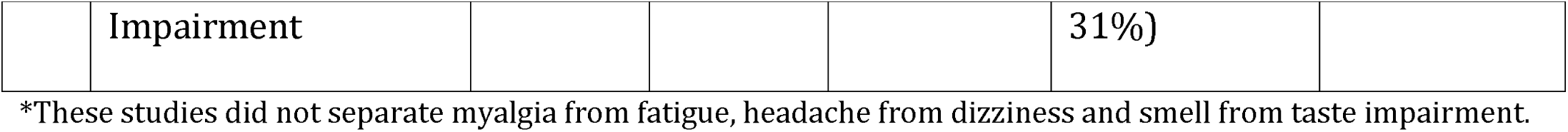
Prevalence of various neurological symptoms included in the systematic review and meta-analysis

Seventeen different neurological diagnoses in COVID-19 patients were observed in 33 studies. The highest number of studies (n=29) were pooled for stroke with a pooled prevalence of 2% (95%CI 1 to 2%) (figure 2). Acute cerebrovascular event was categorized into ischemic stroke or transient ischemic attack 1% (95%CI 1 to 2%, 29 studies), hemorrhagic stroke 0.31% (95%CI 0.15 to 0.50%, 21 studies) and cerebral venous thrombosis 0.12% (95%CI 0 to 2%, two studies). The pooled prevalence of neuropsychiatric disorders was 24% (95%CI 2 to 61%, three studies) while of skeletal muscle injury was 5% (95%CI 1 to 12%, four studies). Pooled prevalence of neurological diagnoses included in the meta-analysis are given in table 2 including the individual prevalence of various neurological diagnoses for which meta-analyses were not possible.

**Table 2:**
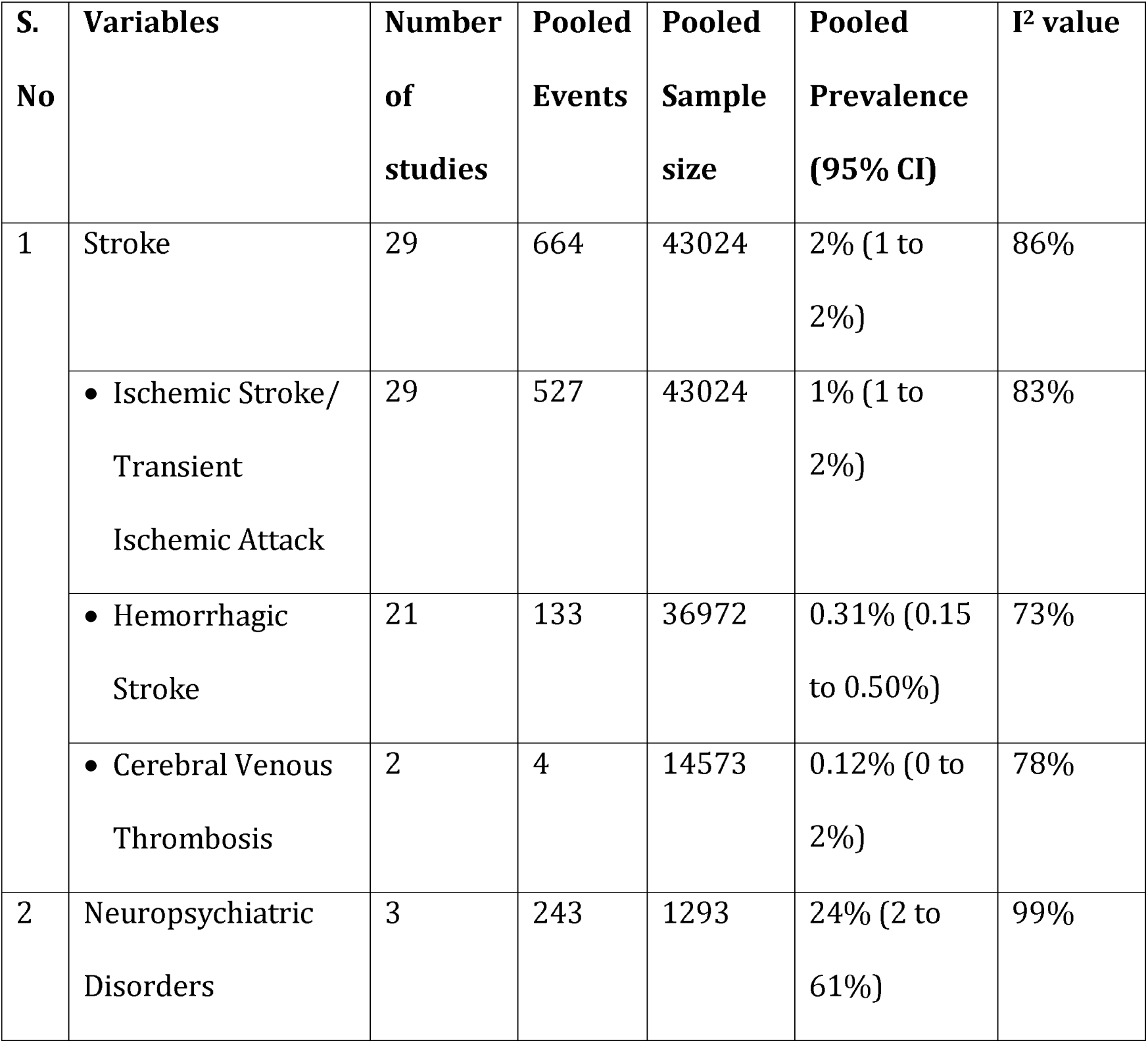
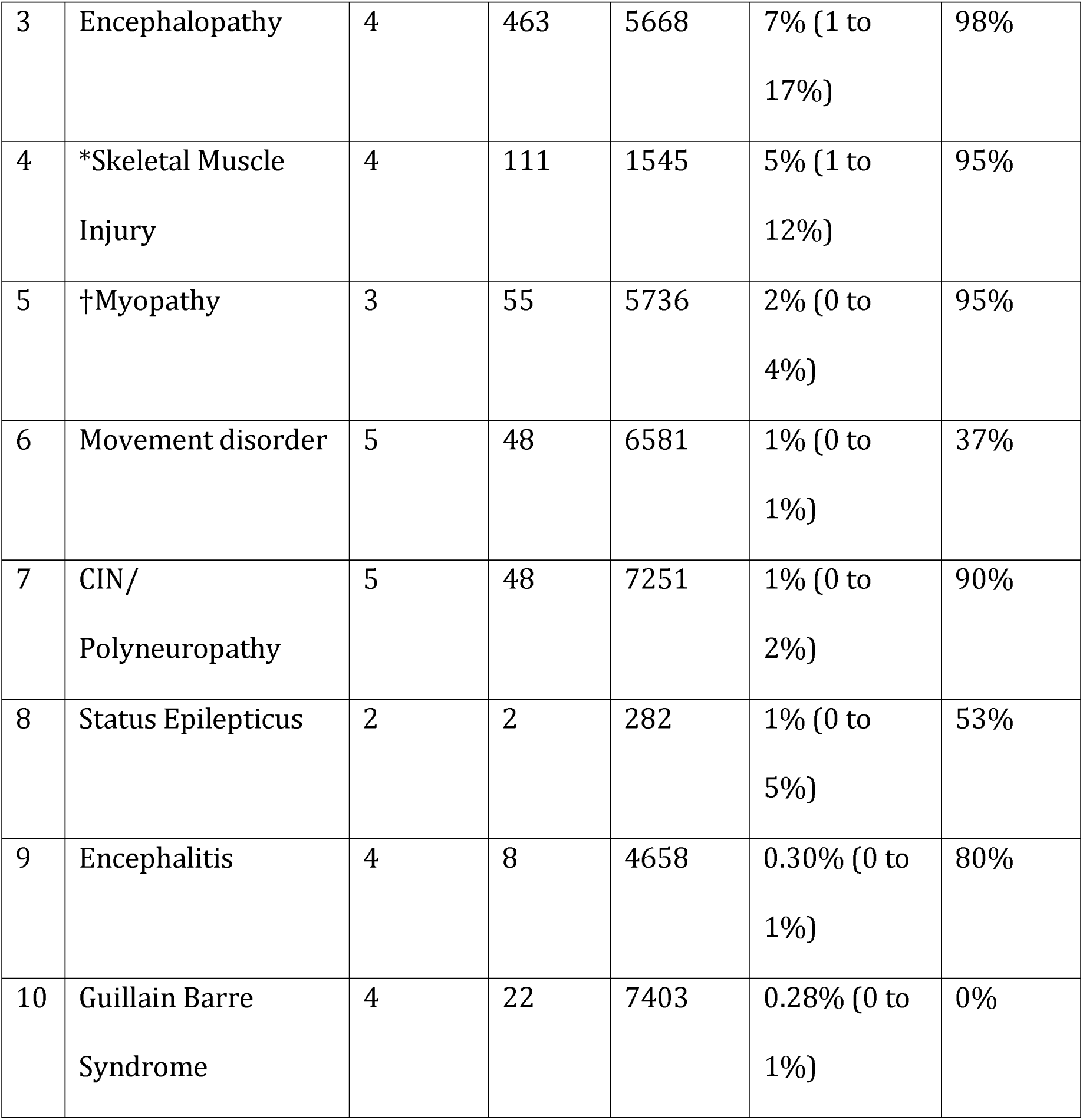
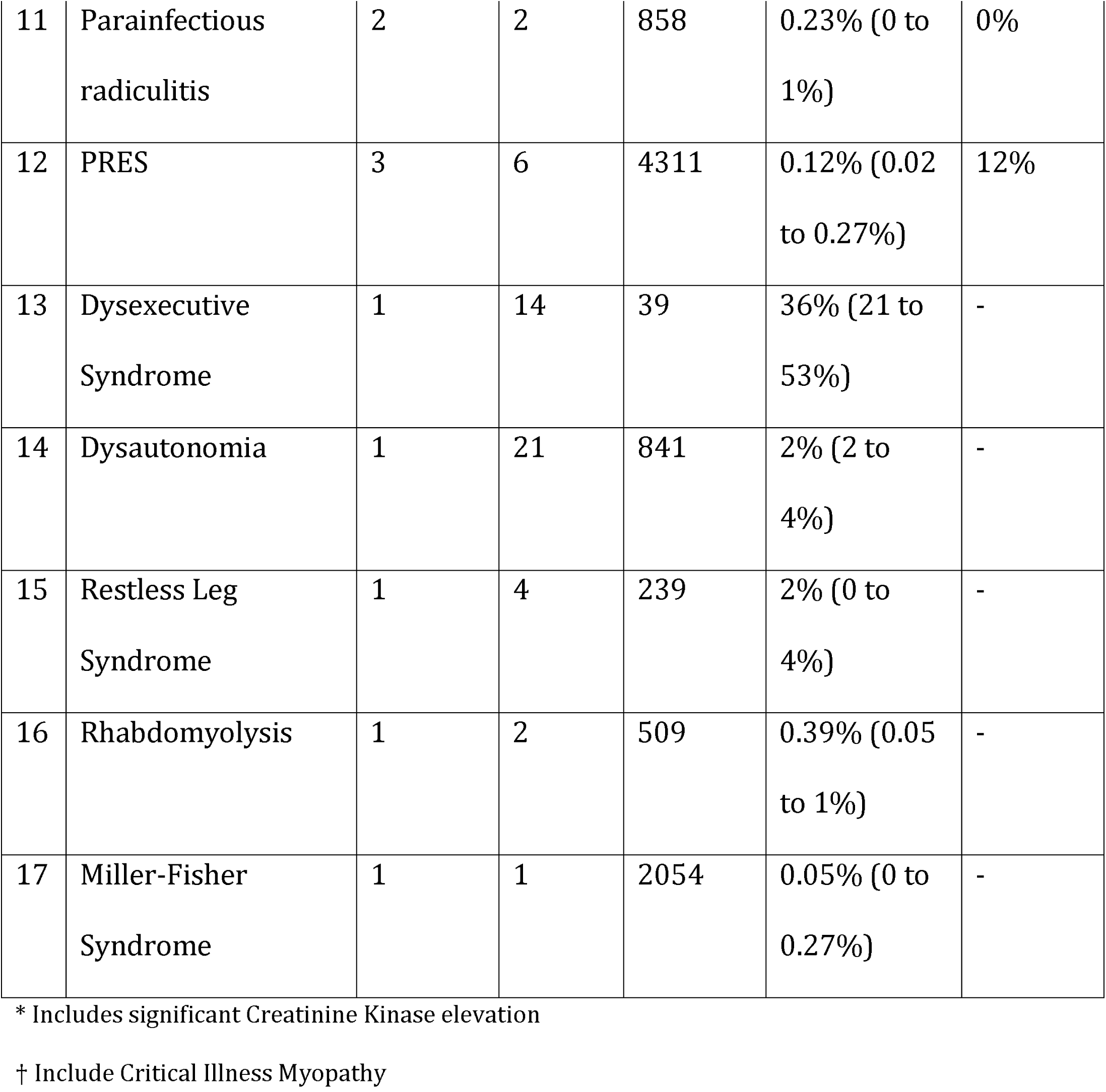
Prevalence of various neurological diagnoses included in the systematic review and meta-analysis

| **S. No** | **Variables** | **Number of studies** | **Pooled Events** | **Pooled Sample size** | **Pooled Prevalence (95% CI)** | **I<sup>2</sup> value** |
| --- | --- | --- | --- | --- | --- | --- |
| 1 | Stroke | 29 | 664 | 43024 | 2% (1 to 2%) | 86% |
|  | • Ischemic Stroke/<br>Transient Ischemic Attack | 29 | 527 | 43024 | 1% (1 to 2%) | 83% |
|  | • Hemorrhagic Stroke | 21 | 133 | 36972 | 0.31% (0.15 to 0.50%) | 73% |
|  | • Cerebral Venous Thrombosis | 2 | 4 | 14573 | 0.12% (0 to 2%) | 78% |
| 2 | Neuropsychiatric Disorders | 3 | 243 | 1293 | 24% (2 to 61%) | 99% |
| 3 | Encephalopathy | 4 | 463 | 5668 | 7% (1 to 17%) | 98% |
| 4 | *Skeletal Muscle Injury | 4 | 111 | 1545 | 5% (1 to 12%) | 95% |
| 5 | †Myopathy | 3 | 55 | 5736 | 2% (0 to 4%) | 95% |
| 6 | Movement disorder | 5 | 48 | 6581 | 1% (0 to 1%) | 37% |
| 7 | CIN/<br>Polyneuropathy | 5 | 48 | 7251 | 1% (0 to 2%) | 90% |
| 8 | Status Epilepticus | 2 | 2 | 282 | 1% (0 to 5%) | 53% |
| 9 | Encephalitis | 4 | 8 | 4658 | 0.30% (0 to 1%) | 80% |
| 10 | Guillain Barre Syndrome | 4 | 22 | 7403 | 0.28% (0 to 1%) | 0% |
| 11 | Parainfectious<br>radiculitis | 2 | 2 | 858 | 0.23% (0 to<br>1%) | 0% |
| 12 | PRES | 3 | 6 | 4311 | 0.12% (0.02<br>to 0.27%) | 12% |
| 13 | Dysexecutive<br>Syndrome | 1 | 14 | 39 | 36% (21 to<br>53%) | - |
| 14 | Dysautonomia | 1 | 21 | 841 | 2% (2 to<br>4%) | - |
| 15 | Restless Leg<br>Syndrome | 1 | 4 | 239 | 2% (0 to<br>4%) | - |
| 16 | Rhabdomyolysis | 1 | 2 | 509 | 0.39% (0.05<br>to 1%) | - |
| 17 | Miller-Fisher<br>Syndrome | 1 | 1 | 2054 | 0.05% (0 to<br>0.27%) | - |

\* Includes significant Creatinine Kinase elevation
† Include Critical Illness Myopathy

**Figure 2:**
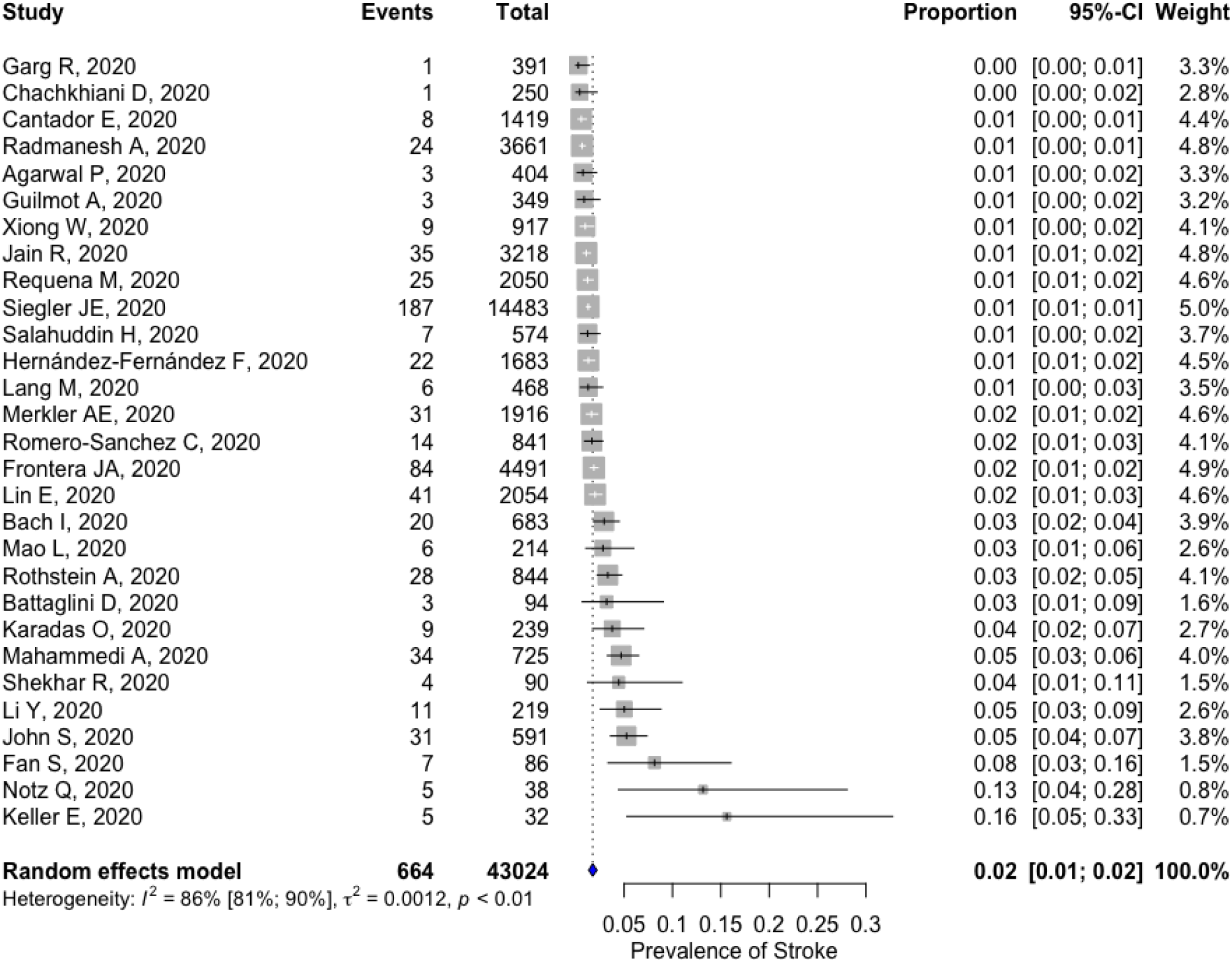
Forest plot depicting the pooled prevalence of stroke

### Neurological manifestations and severity of COVID-19

Forty-eight studies consisting of 2829 severe cases (classified as either severe or critical) and 7493 non-severe cases (classified as either mild or moderate) were analysed to determine the association between neurological manifestations and severity of COVID-19. Skeletal muscle injury/damage in two studies (OR 3.29; 95%CI 2.15 to 5.04), disturbance of consciousness (OR 5.68; 95%CI 2.08 to 15.50) in four studies and fatigue in 33 studies (OR 1.27; 95%CI 1.06 to 1.51) were significantly associated with severe COVID-19.

Patients with severe COVID-19 were less likely than those with mild disease to have alterations in smell (OR 0.44; 95%CI 0.28 to 0.68) in eight studies and taste (OR 0.62; 95%CI 0.42 to 0.91) in five studies. No significant association was found between other neurological manifestations with the severity of COVID-19. Table 3 represents the pooled association of various neurological manifestations with severity of COVID-19 including the individual association of various neurological diagnoses with severity of COVID-19 for which meta-analyses were not possible. No significant deviation was observed after controlling the FDR (supplementary table 3).

**Table 3:** Results from systematic review and meta-analysis of COVID-19 patients (severe and non-severe) developing neurological manifestations (Symptoms and Diagnosis)

| **S. No** | **Variables** | **No. of studies** | **Severe COVID19 (Event/Total)** | **Non-Severe COVID19 (Event/Total)** | **OR (95% CI)** | **P-value** |
| --- | --- | --- | --- | --- | --- | --- |
| 1 | Skeletal Muscle Injury | 2 | 69/417 | 36/638 | **3.29 (2.15 to 5.04)** | **<0.0001** |
| 2 | *Smell Impairment | 8 | 27/670 | 130/1355 | **0.44 (0.28 to 0.68)** | **0.0003** |
| 3 | Disturbance of Consciousness | 4 | 95/450 | 21/652 | **5.68 (2.08 to 15.50)** | **0.0007** |
| 4 | Fatigue | 33 | 916/1968 | 2067/5358 | **1.27 (1.06 to 1.51)** | **0.009** |
| 5 | *Taste Impairment | 5 | 38/617 | 138/1221 | **0.62 (0.42 to 0.91)** | **0.01** |
| 6 | Acute Confusion/<br>Delirium | 4 | 103/1031 | 84/1418 | 3.21 (0.82 to 12.59) | 0.09 |
| 7 | Stroke | 3 | 14/426 | 9/644 | 2.03 (0.83 to 4.96) | 0.12 |
| 8 | Seizure | 3 | 6/426 | 3/644 | 2.40 (0.63 to 9.19) | 0.20 |
| 9 | †Headache &<br>Dizziness | 3 | 24/131 | 64/454 | 1.29 (0.76 to 2.18) | 0.34 |
| 10 | Myalgia | 31 | 477/2146 | 1229/5859 | 1.08 (0.90 to 1.30) | 0.40 |
| 11 | Movement disorder | 2 | 2/463 | 2/887 | 2.02 (0.29 to 13.82) | 0.47 |
| 12 | †Myalgia or Fatigue | 4 | 52/102 | 82/262 | 1.51 (0.32 to 7.11) | 0.60 |
| 13 | Vision Impairment | 2 | 2/417 | 2/638 | 1.55 (0.23 to 10.70) | 0.65 |
| 14 | Headache | 32 | 293/2262 | 799/5998 | 1.05 (0.84 to 1.32) | 0.67 |
| 15 | Dizziness | 11 | 90/1084 | 185/2024 | 0.98 (0.65 to 1.47) | 0.92 |
| 16 | Neuropsychiatric<br>Disorder | 2 | 75/338 | 95/518 | 1.00 (0.30 to 3.31) | 0.99 |
| 17 | Myopathy | 1 | 22/329 | 4/512 | **9.10 (3.11 to 26.66)** | **<0.0001** |
| 18 | Neuralgia | 1 | 4/88 | 1/126 | 5.95 (0.65 to 54.19) | 0.11 |
| 19 | Dysautonomia | 1 | 6/329 | 15/512 | 0.62 (0.24 to 1.60) | 0.32 |
| 20 | Encephalopathy | 1 | 1/134 | 8/375 | 0.34 (0.04 to 2.78) | 0.32 |
| 21 | Rhabdomyolysis | 1 | 1/134 | 1/375 | 2.81 (0.17 to 45.28) | 0.46 |
| 22 | Encephalitis | 1 | 2/9 | 1/6 | 1.42 (0.10 to 20.44) | 0.79 |

\*Probably when severe COVID-19 patients reached hospital they were not in a position to give history for smell and taste impairment. But for completeness of data, we noted that recorded number of patients in smell and taste impairment were more in non-severe group than the severe group.
†These studies did not separate myalgia from fatigue and headache from dizziness.

#### Publication bias

The risk of bias was assessed for neurological manifestations for which 10 or more studies were pooled. The asymmetry of funnel plot was apparent and egger’s test was significant for dizziness (p=0.003), disturbance of consciousness (p=0.006) and stroke (p=0.002).

Twenty Egger’s tests were conducted out of which three were significant (p-value <0.05) and underwent trim and fill analysis. For these three manifestations we used Duval & Tweedie’s Trim-and-fill procedure and observed that studies with a lower prevalence were missing in dizziness, disturbance of consciousness and total stroke. The shape of the funnel plots was symmetric for the rest of the neurological manifestations assessed for the presence of publication bias.

While assessing the association between neurological manifestations and severity of COVID-19, we did not find the presence of significant publication bias for any manifestations assessed using the shape of the funnel plots and Egger’s regression test. The funnel plots and their respective p-values are depicted in supplementary figure 1.

### Meta-regression and subgroup analysis

While assessing the prevalence of neurological manifestations in COVID-19, risk of bias was significantly associated with pooled prevalence for acute confusion/delirium (p-value: 0.001; R^2^= 3.49%) and myalgia or fatigue (p-value: 0.0001; R^2^= 41.09%), where R^2^ is the degree of heterogeneity accounted for. A subgroup analysis by risk of bias for acute confusion/delirium demonstrated that two studies with moderate risk of bias had a pooled prevalence of 42% (95% CI 7 to 83%) while the pooled prevalence dropped to 11% (95% CI 7 to 16%) for 17 studies having a low risk of bias. A similar trend was observed while conducting the subgroup analysis for myalgia or fatigue. Upon stratification, a single study with high risk of bias had high prevalence of 68% (95% CI 48 to 85%), followed by six studies with moderate risk of bias 43% (95% CI 33 to 53%) while the lowest pooled prevalence was observed for 15 studies having a low risk of bias 24% (95% CI 19 to 29%). This subgroup analysis is represented in supplementary table 4 and table 5.

### Neurological symptoms in COVID-19 in the elderly and young sub-populations

Subgroup analyses were conducted on studies that included or disaggregated data on COVID-19 patients aged ≥60 years and those aged <18 years presenting with neurological symptoms. For the elderly, we found 13 studies solely reporting on older patients and two studies with disaggregated data, representing 3176 hospitalized patients presenting with 10 neurological symptoms, with the most prevalent (95%CI, Number of studies) as follows: acute confusion/delirium 34% (95%CI 23 to 46%, five studies); fatigue 20% (95%CI 11 to 31%, nine studies), myalgia 11% (95%CI 7 to 15%, 10 studies); dizziness 5% (95%CI 2 to 9%, three studies) and headache 5% (95%CI 2 to 8%, 10 studies). Table 4 represents the pooled prevalence of neurological symptoms in the elderly COVID-19 sub-population including the individual prevalence of neurological symptoms for which meta-analyses were not possible.

**Table 4:** Prevalence of various neurological symptoms included in the systematic review and meta-analysis of elderly population

| **S. No** | **Variables** | **Number of studies** | **Pooled Events** | **Pooled Sample size** | **Pooled Prevalence (95% CI)** | **I<sup>2</sup> value** |
| --- | --- | --- | --- | --- | --- | --- |
| 1 | Acute Confusion/Delirium | 5 | 433 | 1454 | 34% (23 to 46%) | 94% |
| 2 | Fatigue | 9 | 468 | 2186 | 20% (11 to 31%) | 96% |
| 3 | Myalgia | 10 | 302 | 2642 | 11% (7 to 15%) | 90% |
| 4 | Dizziness | 3 | 84 | 1211 | 5% (2 to 9%) | 83% |
| 5 | Headache | 10 | 161 | 2398 | 5% (2 to 8%) | 90% |
| 6 | *Myalgia or Fatigue | 1 | 60 | 118 | 51% (41 to 60%) | - |
| 7 | *Headache & Dizziness | 1 | 15 | 118 | 13% (7 to 20%) | - |
| 8 | Disturbance of<br>Consciousness | 1 | 10 | 218 | 5% (2 to 8%) | - |
| 9 | Taste Impairment | 1 | 4 | 76 | 5% (1 to 3%) | - |
| 10 | Smell Impairment | 1 | 1 | 76 | 1% (0 to 7%) | - |

\*These studies did not separate myalgia from fatigue and headache from dizziness.

For COVID-19 patients aged <18 years, we found 20 studies with 3051 hospitalized patients presenting with 10 neurological symptoms, with the most prevalent (95%CI, Number of studies) as follows: fatigue or myalgia 17% (95%CI 9 to 26%, two studies); smell or taste impairment 13% (95%CI 6 to 22%, two studies); headache 10% (95%CI 5 to 15%, 13 studies); fatigue 9% (95%CI 3 to 18%, nine studies); myalgia 7% (95%CI 2 to 15%, five studies); seizure 4% (95%CI 2 to 6%, two studies) and smell impairment 3% (95%CI 1 to 5%, five studies). Table 5 represents the pooled prevalence of neurological symptoms in the young COVID-19 sub-population including the individual prevalence of neurological symptoms for which meta-analyses were not possible.

**Table 5:** Prevalence of various neurological symptoms included in the systematic review and meta-analysis of young population

| **S. No** | **Variables** | **Number of studies** | **Pooled Events** | **Pooled Sample size** | **Pooled Prevalence (95% CI)** | **I<sup>2</sup> value** |
| --- | --- | --- | --- | --- | --- | --- |
| 1 | *Fatigue or Myalgia | 2 | 16 | 89 | 17% (9 to 26%) | 0% |
| 2 | *Smell or Taste Impairment | 2 | 9 | 68 | 13% (6 to 22%) | 0% |
| 3 | Headache | 13 | 174 | 1615 | 10% (5 to 15%) | 86% |
| 4 | Fatigue | 9 | 168 | 1431 | 9% (3 to 18%) | 93% |
| 5 | Myalgia | 5 | 57 | 1422 | 7% (2 to 15%) | 92% |
| 6 | Seizure | 2 | 30 | 694 | 4% (2 to 6%) | 0% |
| 7 | Smell Impairment | 5 | 11 | 346 | 3% (1 to 5%) | 0% |
| 8 | Dizziness | 1 | 2 | 12 | 17% (2 to 48%) | - |
| 9 | Disturbance of<br>Consciousness | 1 | 24 | 651 | 4% (2 to 5%) | - |
| 10 | Taste Impairment | 1 | 2 | 105 | 2% (0 to 7%) | - |

\*These studies did not separate myalgia from fatigue and smell from taste impairment.

### Neurological symptoms in COVID-19 cases analysed by hospitalization status

We included 129,825 hospitalized and 9,188 non-hospitalized COVID-19 cases. Importantly, the data for neurological symptoms in non-hospitalized COVID-19 cases could only be extracted from 13 studies. Smell impairment (50%), taste impairment (44%), headache (31%) and myalgia (31%) were found to be more common in non-hospitalized cases, whereas fatigue (31%) and myalgia or fatigue (30%) were found to be more common in hospitalized cases. A full comparison is available in supplementary table 8.

### Mortality in COVID-19 patients with and without neurological manifestations

Mortality in COVID-19 patients having one or more neurological manifestations was assessed in 21 studies included in the systematic review and meta-analysis. A total of 770 patients died out of 2982 experiencing neurological manifestations. The pooled prevalence of mortality in COVID-19 patients with one or more neurological manifestations was 27% (95%CI 19 to 35%).

The association between mortality and neurological manifestations was assessed in 13 studies consisting of 2706 and 10808 COVID-19 patients with and without neurological manifestations, respectively. There was a non-significant trend towards a higher odds of mortality in patients with neurological manifestations compared to those without them (OR 1.39; 95%CI 0.92 to 2.11). For patients ≥60 years, the presence of neurological manifestations was significantly associated with increased mortality (OR 1.80; 95%CI 1.11 to 2.91) (figure 3).

**Figure 3:**
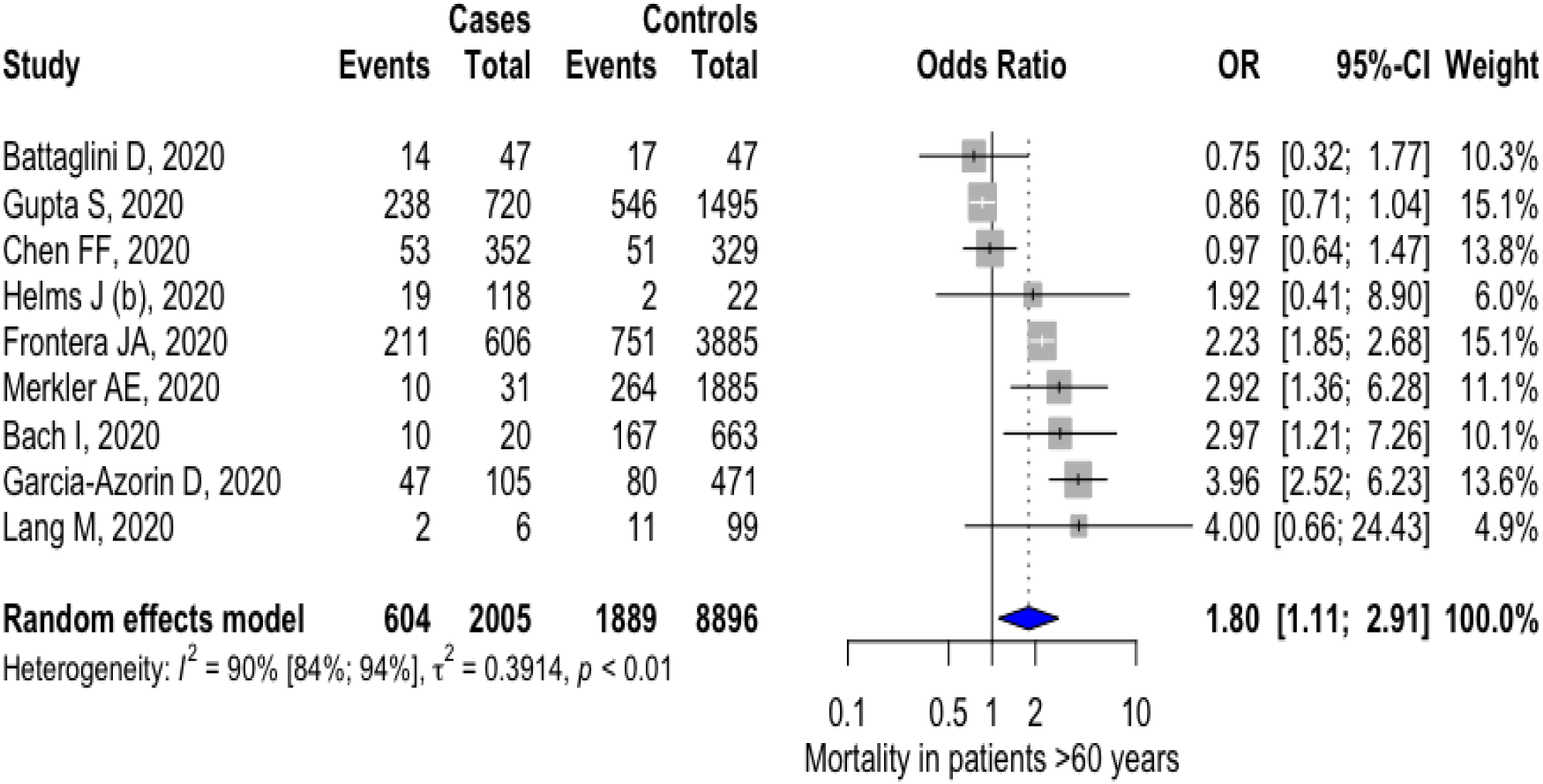
Forest plot depicting the association between presence of neurological manifestations and mortality of COVID-19 patients aged >60 years

## Discussion

It is increasingly apparent that SARS-CoV-2 can have many effects on the central and peripheral nervous system. In this review, we report on all acute neurological manifestations reported in COVID-19, comorbidities associated with COVID-19 infection, the association with acute neurological manifestations and severity or mortality, as well as sub-group analyses of neurological manifestations by age group. Our pooled prevalence estimates indicate that up to one-third of COVID-19 patients analysed in our review experienced at least one neurological manifestation and one in 50 people developed a stroke.

Our review shows that pre-existing neurological conditions are a common comorbidity associated with COVID-19. After hypertension and diabetes, the combined comorbidity of cardiovascular/cerebrovascular disease was the third most commonly reported comorbidity associated with COVID-19. Disaggregated data released from national and international COVID-19 registries similarly indicates a high prevalence of pre-existing neurological conditions and additionally an association with severe disease.^23–25^

### Acute neurological manifestations commonly reported in COVID-19

This review identified 41 neurological manifestations (24 symptoms and 17 diagnoses). The following neurological symptoms were found to be common in a large number of studies: fatigue, myalgia, smell and taste impairment, headaches and agitation. Neuropsychiatric disorders were found to affect 1 in 4 hospitalised patients, though data were only available from three studies. Critically, the pooled prevalence in our review for the diagnosis of stroke was 2%, with data extracted from 29 studies; the majority of studies reported prevalence between 1 to 3%. To our knowledge this is the first time that pooled prevalence for stroke has been reported in COVID-19 patients and indicates an alarming, enduring neurological morbidity associated with the pandemic. Additionally, we note that this pooled prevalence is far higher than has previously been reported for patients with influenza. One recent study^26^ compared the risk of stroke in COVID-19 versus influenza and found the prevalence of stroke to be 0.2% in influenza versus 1.2% in COVID-19.

### Brain involvement more common in older persons

Acute confusion/delirium is significantly more common in older people who have COVID-19, with data from five studies demonstrating 1 in 3 hospitalised older COVID-19 patients are affected (pooled prevalence 34%), compared with 5% in young adults and 12% for all ages. This bears implications for both cognitive outcomes and mortality. Delirium in older hospitalized patients has previously been demonstrated to be an independent risk factor for mortality one year after hospitalization, increasing the risk of death by over two-fold.^27^ Moreover, delirium in older adults is a risk factor for developing dementia, increasing the risk nearly nine-fold over ten years of follow-up for adults 85 years and older.^28^

### Acute neurological manifestations associated with severity and mortality in elderly

For adults with COVID-19 aged over 60 years, the presence of any neurological manifestation was associated with increased mortality (OR 1.80; 95%CI 1.11 to 2.91). We also note that the relationship between COVID-19 severity and neurological manifestations may be bidirectional, as critically ill patients are more likely to experience multi-organ system dysfunction.

### Alterations in smell and taste associated with non-severe disease

It has been posited that there may be pathophysiological basis for the association between smell and taste impairment and mild COVID-19, as inflammation in mucosal membranes may play a role in preventing migration of the virus elsewhere in the body.^29^ Moreover, patients with severe COVID-19 may not have been able to provide a clear history regarding smell or taste impairment, and we note that the majority of the data we have analysed comes from hospitalized patients.^30^

### Potential mechanisms for acute neurological manifestations of COVID-19

Multiple, potentially interacting, mechanisms have been proposed for the neurological manifestations including: hypoxia, severe cytokine storm during infection and post-infectious autoimmune responses, hypercoagulability, endotheliopathy, multiple organ failure (e.g., liver failure, leading to metabolic derangement), and possibly direct neuroinvasion. The neuro-invasive and neurotropic potential of SARS-CoV-2 is controversial but it may be explained by the expression of Angiotensin-converting Enzyme-2 (ACE-2) receptor in the brain.^31^

### Limitations of this review

There are several important limitations for this review. Firstly, there are few data from prospective studies; most data came from retrospective cohorts or case series. Additionally, standardized classifications, definitions, and diagnostic criteria of neurological manifestations were not uniformly used or reported. Moreover, studies relating to many manifestations were not suitable for pooling. Thirdly, as the vast majority of data analyzed came from hospitalized cases (89%), our pooled prevalence estimates of neurological manifestations, especially those of severe manifestations such as stroke, may not represent community prevalence. We are therefore unable to draw conclusions regarding the prevalence of neurological manifestations amongst COVID-19 patients in the community. Additionally, interpretation of data regarding pre-existing neurological conditions and COVID-19 diagnoses is limited due to the proportion of hospitalized cases studied.

Meta-analyses for most outcomes in this review had a high degree of heterogeneity, which could only partly be explained by meta-regression and subgroup analyses. It is expected that the patient populations in the included studies were clinically diverse, having presented to centers with varied referral pathways, and this may have contributed to the heterogeneity observed. For almost all symptoms, studies with higher risk of bias yielded higher pooled prevalence, indicating overestimation from bias in selection and/or measurement. Bias is known to exaggerate effect estimates in studies.^32^ Studies were varying in terms of biases in selection, measurement and reporting. Severity and mortality analyses in this review are limited by confounding bias, as given estimates were not adjusted for potential confounders such as comorbidities.

Moreover, the timing of symptom onset is not reported in the majority of studies, which bears implications for understanding causality and as such we are unable to distinguish acute neurological manifestations caused by COVID-19 from the incidence of similar symptoms and diagnoses in the general population. Additionally, this review has mainly focused on new-onset neurological symptoms. We have not analysed worsening of underlying or pre-existing neurological disorders, for which there are data emerging in relation to COVID-19 and dementia, Parkinson’s disease, epilepsy,^33^ myasthenia gravis^34^ and multiple sclerosis.^35^

And critically, the majority of the data analysed in this review are from high and middle-income countries. There is little data from low-income countries on neurological manifestations available in published studies to date.

### Implications for clinical practice and further research

It is our hope that the data provided in this review might aid clinical practice and future research. Pooled prevalence estimates reported in this paper may be used to design and/or validate clinical checklists for detecting neurological manifestations that may otherwise be missed. Additionally, neurological manifestations may be factored into risk stratification, especially for older persons with COVID-19, given the association with higher mortality for any neurological manifestation in this subpopulation.

Several questions remain as to the characteristics, timing, and severity of neurological manifestations of COVID-19. Well-designed cohort, case-control and population-based studies are required to understand risk factors and suggest underlying mechanisms, which should be confirmed by neuropathological studies. The timing of risk for acute, subacute, and long-term neurological manifestations remains unclear, and this is particularly relevant for stroke and cognitive impairment. Importantly, it is vital to have data on neurological manifestations from geographically and resource diverse settings. One year into the pandemic, it is critical to have robust, harmonised standards for methodology and reporting in order to minimize the risk of bias and enhance confidence in our understanding of the true neurological implications of this illness.

## Data Availability

The full dataset and statistical codes will be available upon reasonable request.

## Disclaimer

The authors alone are responsible for the views expressed in this article and they do not necessarily represent the views, decisions or policies of the institutions with which they are affiliated.

## Contributors

KP conceptualised the idea of the review, helped design the methodology, supervised each step of execution of the review, and invited the international authors. SM, DR, KKT, AK and AD contributed to the literature search. SM and DR conducted the data extraction. SM, DR, MP, AK, KTT and AD conducted the risk of bias analysis. SM and MP conducted the statistical analysis. SM, KK, TD, KP, MP and DR designed and drafted the figures and tables. SM, KK, TD, KP, MP and DR prepared the initial manuscript draft. TS, ASW, AG, ELF, BDM, EB, GKW, CAP, SHYC, FKH, ES, AKM, AD, AKS and AA critically reviewed the initial draft. All authors contributed to, reviewed, and approved the final draft of the paper.

## Acknowledgments

KP received funding from WHO to support this systematic review and meta-analysis. SM is a DST-INSPIRE Fellow supported by Department of Science and Technology, Government of India. SHYC receives research funding from the National Institutes of Health (NIH) National Center for Advancing Translational Sciences (NCATS) through Grant Number(s) UL1 TR001857 (University of Pittsburgh/Chou), the National Institute of Neurological Disorders and Stroke (R21-NS113037) & the University of Pittsburgh School of Medicine Dean’s Faculty Advancement Award. ELF has a research grant from Neurocritical Care Society INCLINE grant, NIH (NINDS) R01 NS096714. CAP is supported by NIH R01 NS110122 and Bart McLean Fund for Neuroimmunology Research. BDM is supported to conduct clinical COVID-19 neuroscience research by the UKRI/MRC (MR/V03605X/1); for additional neurological inflammation research due to SARS-CoV2 and associated viral infection BDM is also supported by grants from the MRC/UKRI (MR/V007181//1), MRC (MR/T028750/1) and Wellcome (ISSF201902/3). TS is supported by the National Institute for Health Research (NIHR) Health Protection Research Unit in Emerging and Zoonotic Infections (Grant Nos. IS-HPU-1112-10117 and NIHR200907), NIHR Programme Grant for Applied Research (No. RP-PG-0108-10,048), NIHR Global Health Research Group on Brain Infections (No. 17/63/110), and the European Union’s Horizon 2020 research and innovation program ZikaPLAN (Preparedness Latin America Network), grant agreement No. 734584.

## Ethics approval

Not required

## Data sharing

The full dataset and statistical codes will be available upon reasonable request.

The lead author (SM) affirms that the manuscript is an honest, accurate, and transparent account of the study being reported; that no important aspects of the study have been omitted; and that any discrepancies from the study as planned have been explained.

## Dissemination to participants and related patient and public communities

No specific plan beyond dissemination through journal publication and news media.

