## Supplementary file for "Frequency of neurological manifestations in COVID-19: a systematic review and meta-analysis of 350 studies"

**(SUPPLEMENTARY MATERIAL)**

**Supplementary Methods**

**Detailed Search strategy**

1. (("severe acute respiratory syndrome coronavirus 2"[Supplementary Concept] OR "severe acute respiratory syndrome coronavirus 2"[All Fields] OR "ncov"[All Fields] OR "2019-nCoV"[All Fields] OR "COVID-19"[All Fields] OR "SARS-CoV-2"[All Fields] OR ((coronavirus[All Fields] OR "cov"[All Fields]) AND 2019/11[PubDate] : 3000[PubDate])) AND neurological[All Fields] AND "features"[All Fields]) AND ("2019/12/31"[PubDate] : "2020/12/15"[PubDate])
2. (("severe acute respiratory syndrome coronavirus 2"[Supplementary Concept] OR "severe acute respiratory syndrome coronavirus 2"[All Fields] OR "ncov"[All Fields] OR "2019-nCoV"[All Fields] OR "COVID-19"[All Fields] OR "SARS-CoV-2"[All Fields] OR ((coronavirus[All Fields] OR "cov"[All Fields]) AND 2019/11[PubDate] : 3000[PubDate])) AND ("neurologic manifestations"[MeSH Terms] OR ("neurologic"[All Fields] AND "manifestations"[All Fields]) OR "neurologic manifestations"[All Fields] OR ("neurological"[All Fields] AND "manifestations"[All Fields]) OR "neurological manifestations"[All Fields])) AND ("2019/12/31"[PubDate] : "2020/12/15"[PubDate])
3. (("coronavirus"[MeSH Terms] OR "coronavirus"[All Fields]) AND ("neurologic manifestations"[MeSH Terms] OR ("neurologic"[All Fields] AND "manifestations"[All Fields]) OR "neurologic manifestations"[All Fields] OR ("neurological"[All Fields] AND "symptoms"[All Fields]) OR "neurological symptoms"[All Fields]) AND ("diagnosis"[Subheading] OR "diagnosis"[All Fields] OR "signs"[All Fields] OR "diagnosis"[MeSH Terms] OR "signs"[All Fields])) AND ("2019/12/31"[PubDate] : "2020/12/15"[PubDate])
4. (("severe acute respiratory syndrome coronavirus 2"[Supplementary Concept] OR "severe acute respiratory syndrome coronavirus 2"[All Fields] OR "ncov"[All Fields] OR "2019-nCoV"[All Fields] OR "COVID-19"[All Fields] OR "SARS-CoV-2"[All Fields] OR ((coronavirus[All Fields] OR "cov"[All Fields]) AND 2019/11[PubDate] : 3000[PubDate])) AND ("brain"[MeSH Terms] OR "brain"[All Fields]) AND "features"[All Fields]) AND ("2019/12/31"[PubDate] : "2020/12/15"[PubDate])
5. (("severe acute respiratory syndrome coronavirus 2"[Supplementary Concept] OR "severe acute respiratory syndrome coronavirus 2"[All Fields] OR "sars cov 2"[All Fields]) AND ("central nervous system"[MeSH Terms] OR ("central"[All Fields] AND "nervous"[All Fields] AND "system"[All Fields]) OR "central nervous system"[All Fields])) AND ("2019/12/31"[PubDate] : "2020/12/15"[PubDate])
6. (Clinical[All Fields] AND "features"[All Fields]) AND ("covid-19"[MeSH Terms] OR "covid-19"[All Fields] OR "covid19"[All Fields]) AND ("2019/12/31"[PubDate] : "2020/12/15"[PubDate])
7. (Clinical[All Fields] AND "features"[All Fields]) AND ("coronavirus"[MeSH Terms] OR "coronavirus"[All Fields]) AND ("2019/12/31"[PubDate] : "2020/12/15"[PubDate])

**Definitions**

1. Neuropsychiatric disorders were defined as depression, anxiety, psychosis, paranoia, hallucinations, irritability, insomnia or impairment of sleep.
2. Disturbance of consciousness was defined as disturbances of both level and content of consciousness and spans from confusion to coma.
3. Corticospinal tract signs were defined as diffuse hyperreflexia, ankle clonus and bilateral extensor plantar reflexes.
4. Hyposmia and anosmia were included under the symptom “Smell Impairment”.
5. Ageusia and Dysgeusia were included under the symptom “Taste Impairment”.
6. Mild and moderate cases of COVID-19 were categorized as “non-severe COVID-19”.
7. Severe and critical cases of COVID-19 were categorized as “severe COVID-19”.

**Supplementary Figures**

| **I. Prevalence of Neurological symptoms/signs and diagnosis in COVID-19** | |
| --- | --- |
| **Dizziness (N=46)**  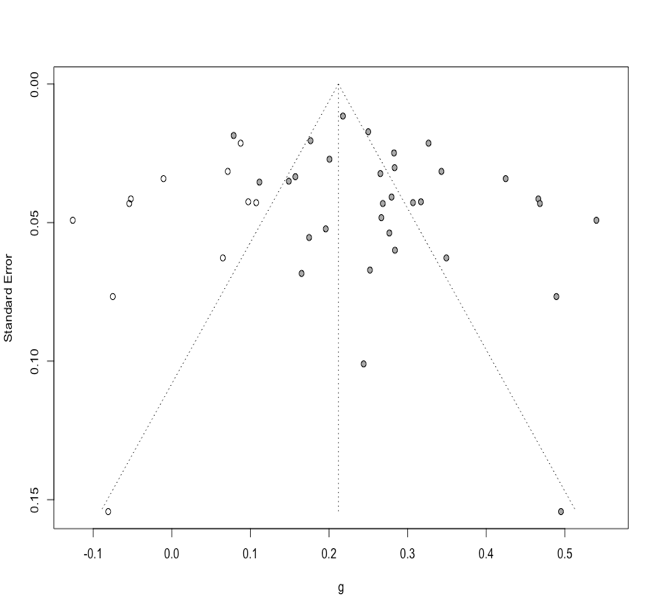  **P-value: 0.003** | **Disturbance of Consciousness (N=25)**  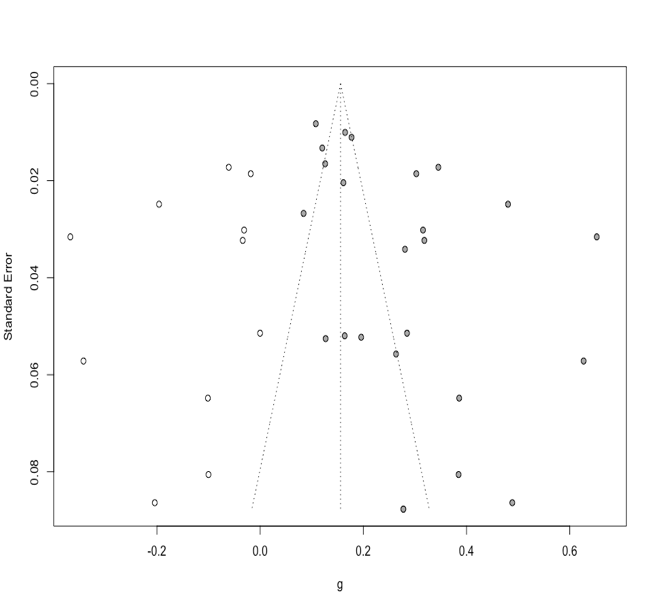  **P-value: 0.007** |
| **Myalgia or Fatigue (N=22)**  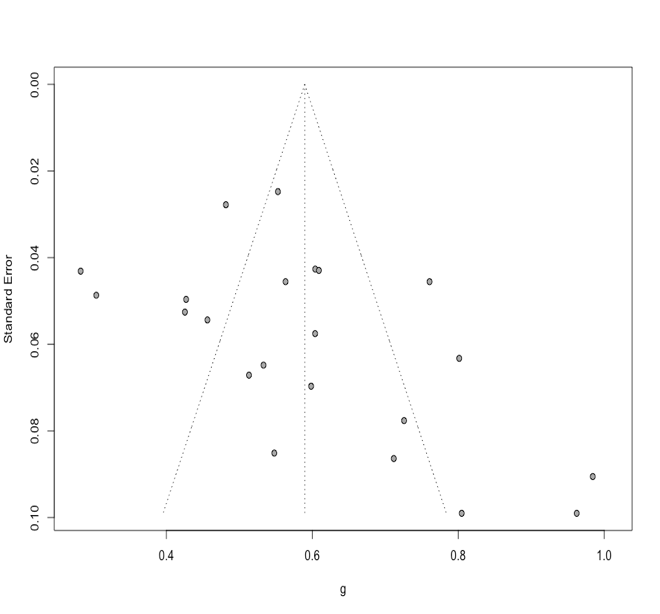  **P-value:** 0.06 | **Fatigue (N=169)**  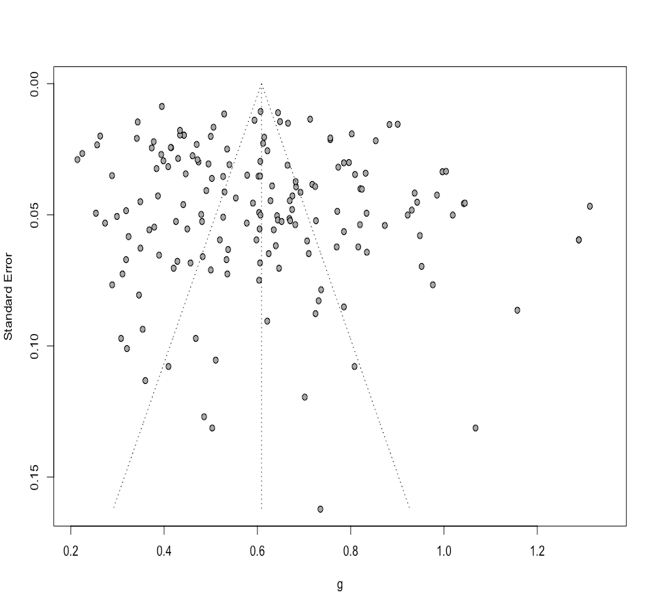  P-value: 0.07 |
| **Smell Impairment (N=51)**  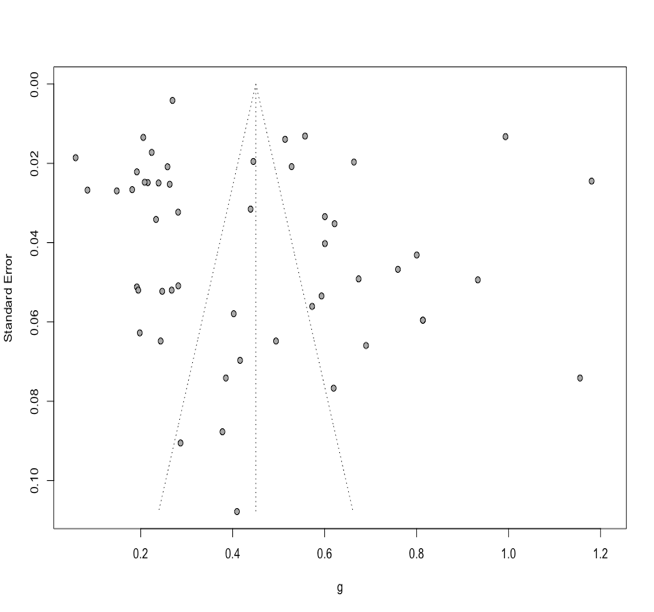  P-value: 0.07 | **Seizure (N=15)**  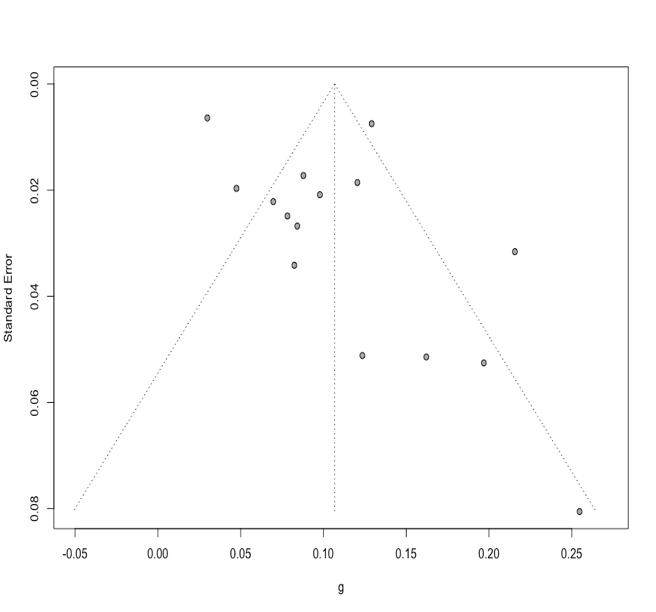  P-value: 0.14 |
| **Myalgia (N=207)**  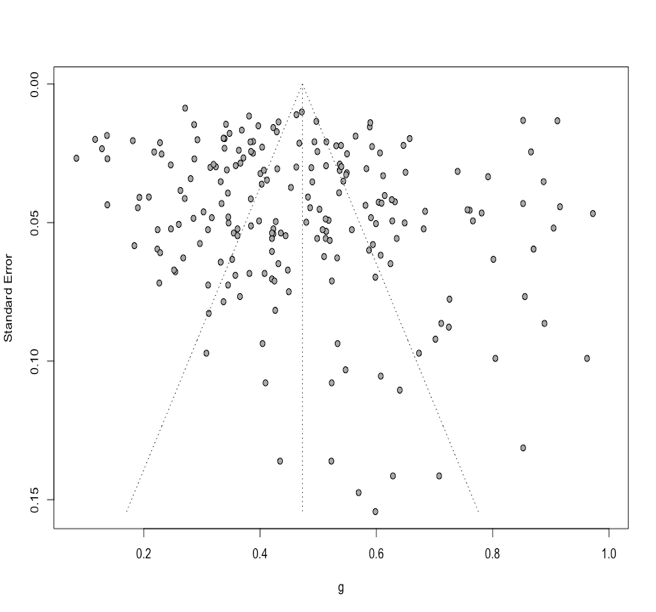  P-value: 0.18 | **Vision Impairment (N=10)**  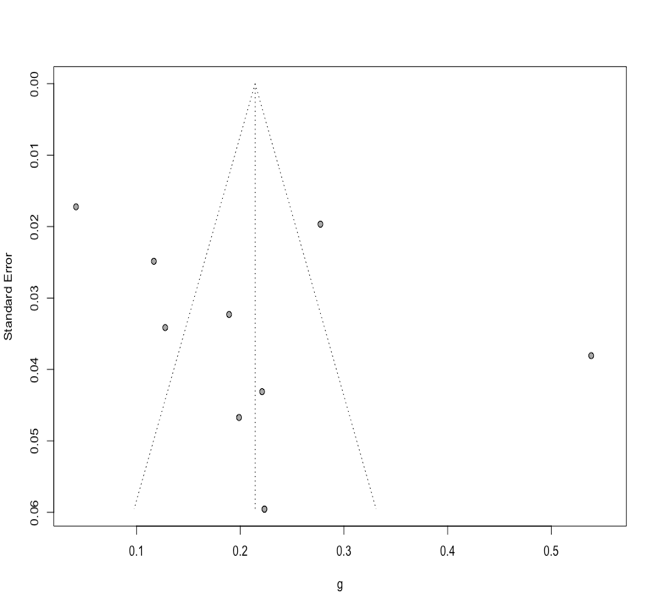  P-value: 0.26 |
| **Smell or Taste Impairment (N=14)**  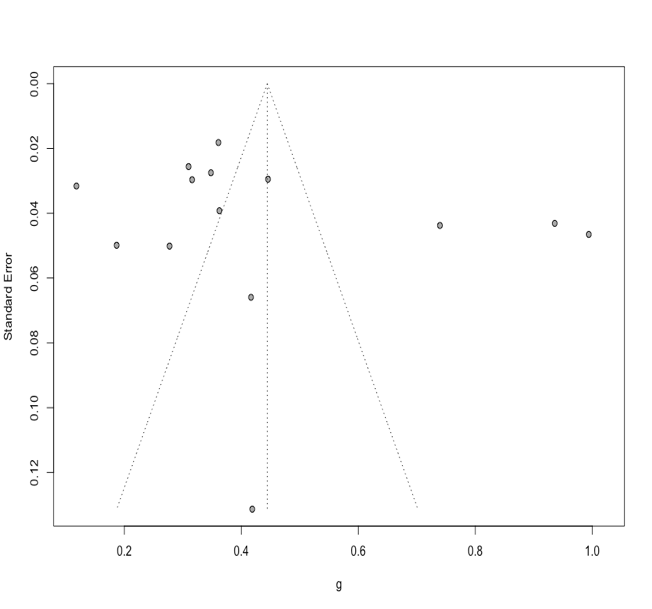  P-value: 0.29 | **Headache (N=202)**  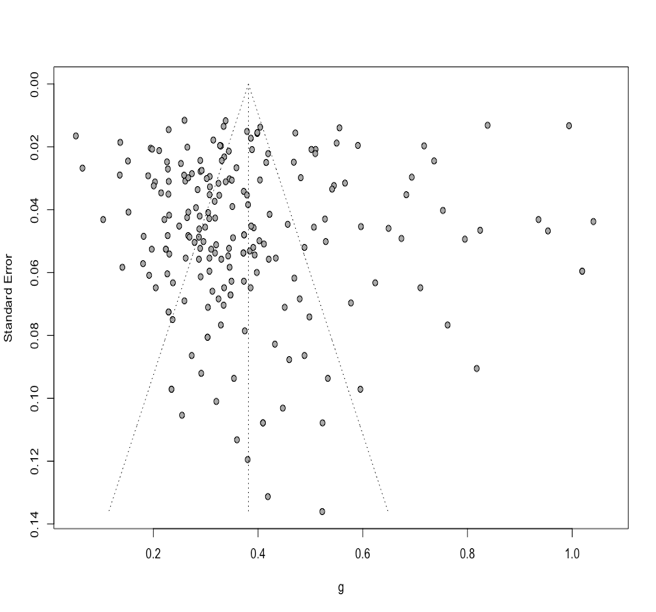  P-value: 0.29 |
| **Acute confusion/delirium (N=19)**  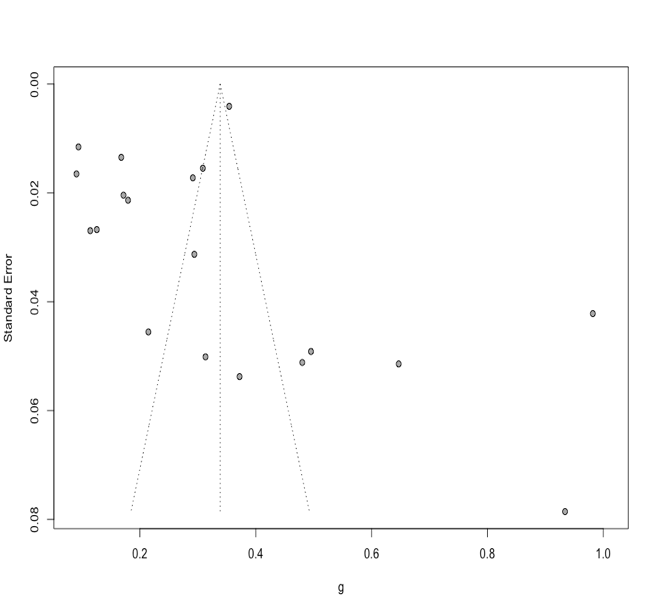  P-value: 0.63 | **Taste Impairment (N=38)**  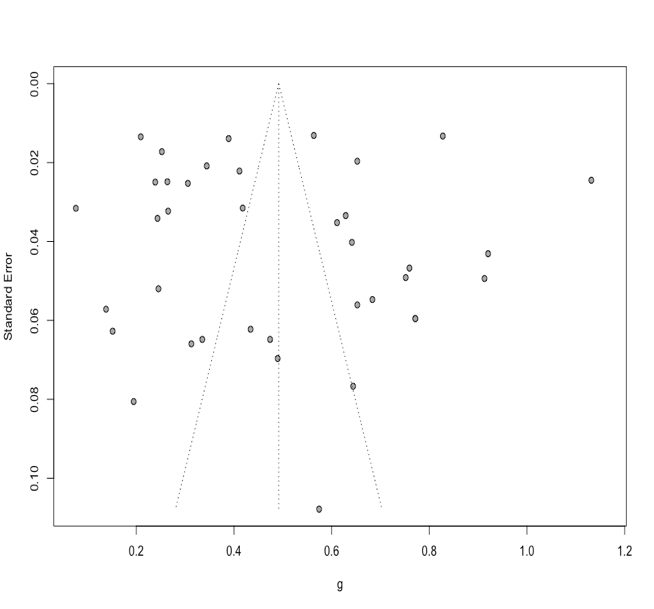  P-value: 0.82 |
| **Total Stroke (N=22)**  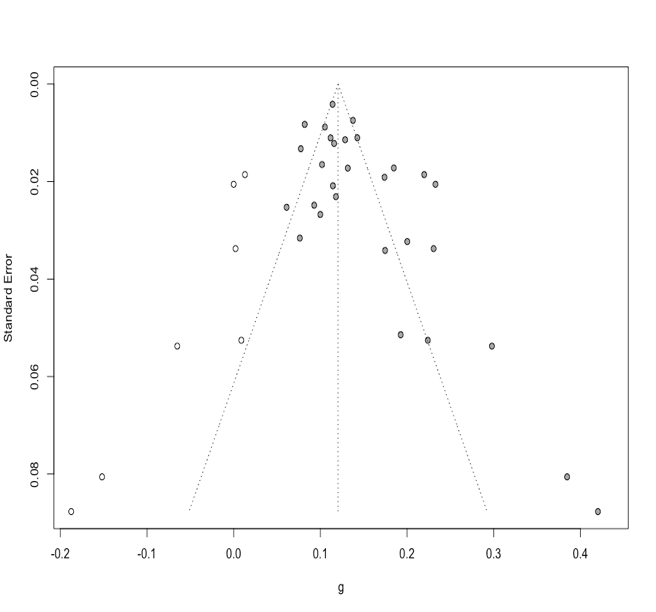  **P-value: 0.007** |  |
| **II. Neurological manifestations and severity of COVID-19** | |
| **Myalgia (N=31)**  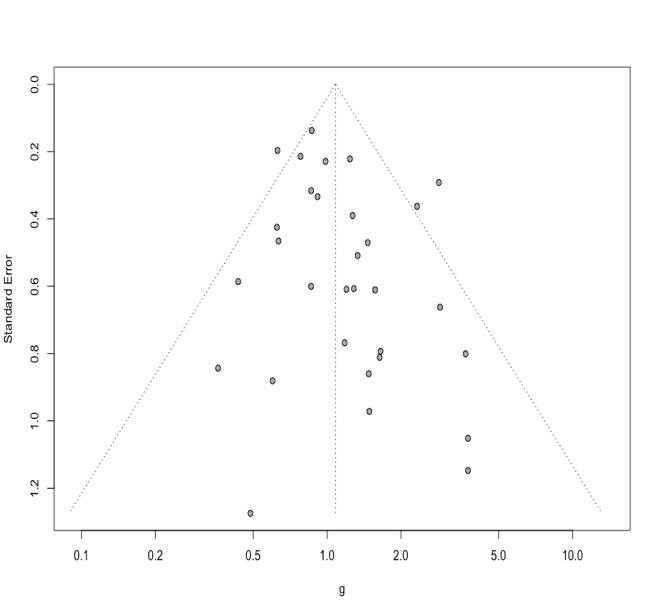  P-value: 0.09 | **Fatigue (N=33)**  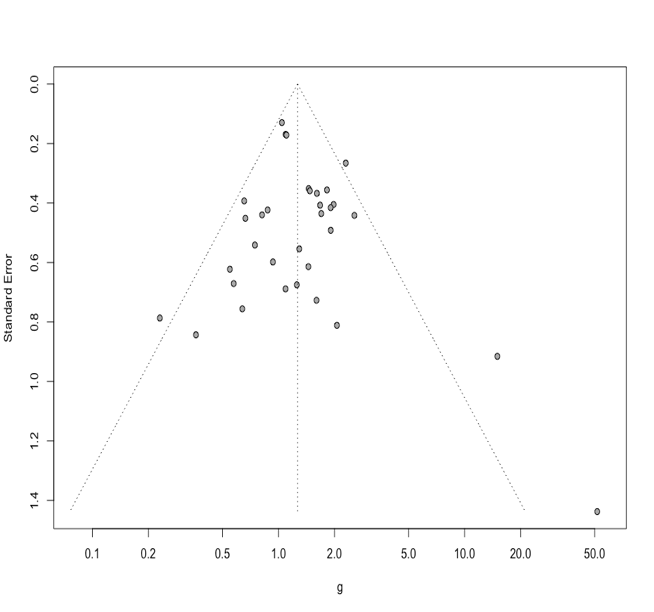  P-value: 0.39 |
| **Headache (N=32)**  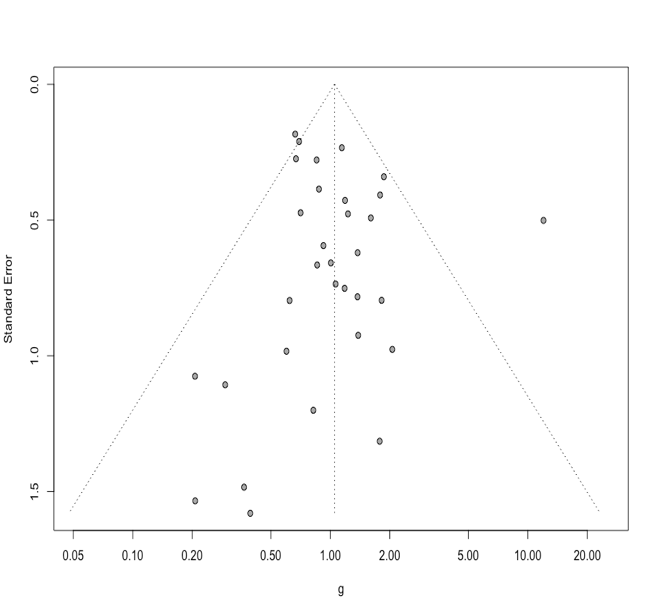  P-value: 0.44 | **Dizziness (N=11)**  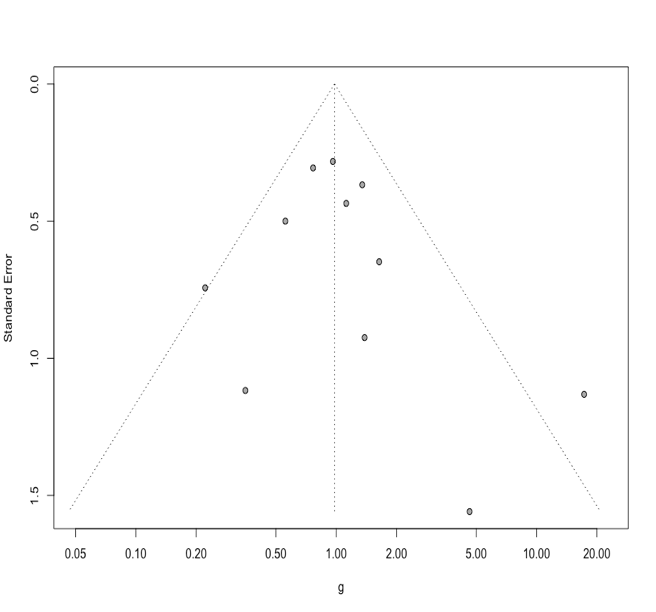  P-value: 0.49 |
| **III.** **Prevalence of Neurological symptoms/signs in COVID-19 in Elderly population** | |
| **Headache (N=10)**  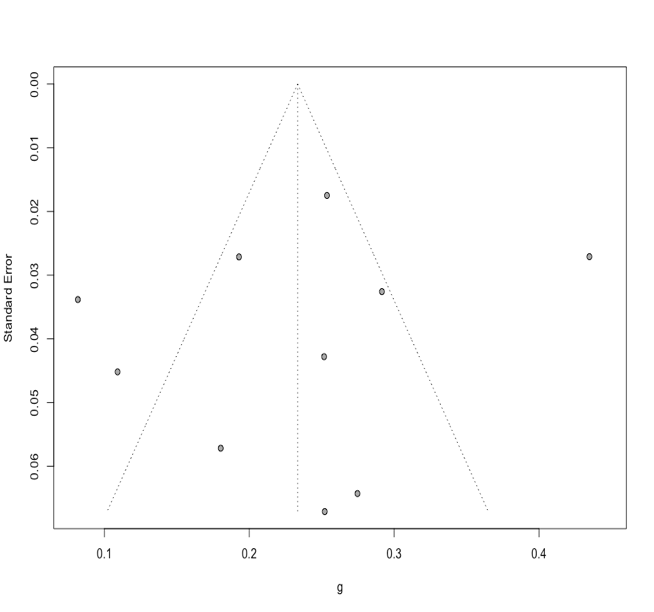  P-value: 0.55 | **Myalgia (N=10)**  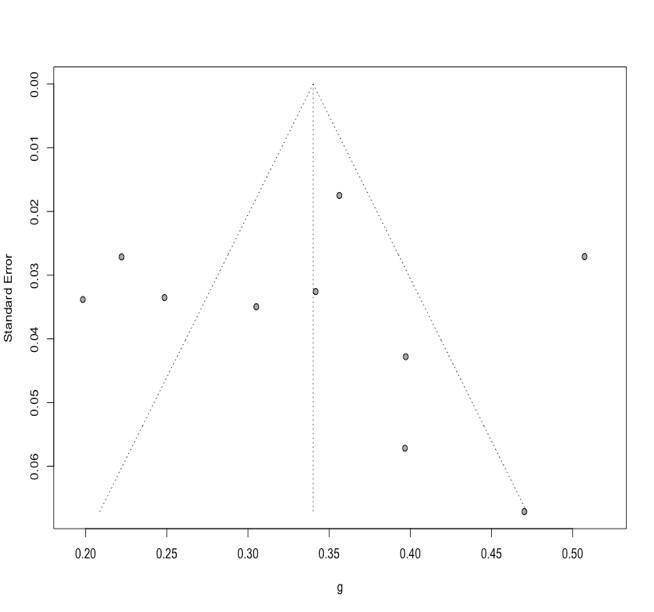  P-value: 0.97 |
| **IV. Prevalence of Neurological symptoms/signs in COVID-19 in Young population** | |
| **Headache (N=13)**  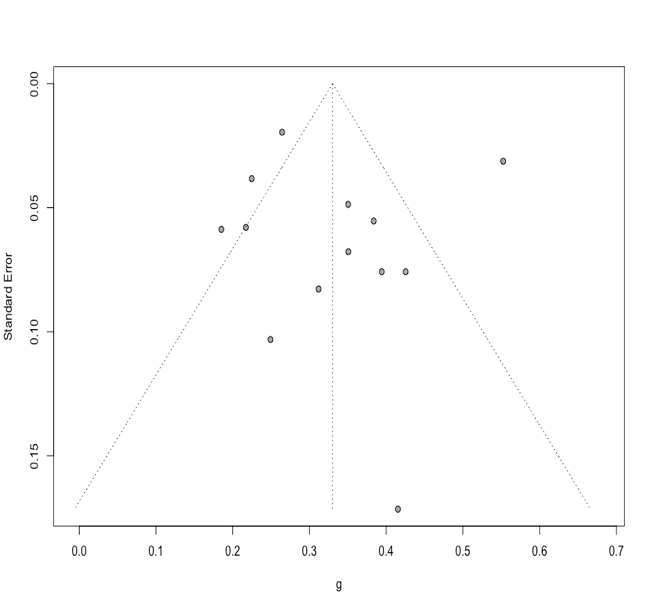  P-value: 0.78 |  |

***Bold p-values: <0.05**

**Figure 1:** Publication bias analysis using funnel plots for various neurological manifestations in COVID-19.

**Supplementary Tables**

**Table 1:** List of distinct countries included in our systematic review and meta-analysis

| **S. No** | **Country** | **World Bank Income bracket** | **Number of studies** |
| --- | --- | --- | --- |
| 1 | Australia | HIC | 3 |
| 2 | Austria | HIC | 1 |
| 3 | Belgium | HIC | 1 |
| 4 | Bolivia | LMIC | 1 |
| 5 | Brazil | UMIC | 3 |
| 6 | Bulgaria | UMIC | 1 |
| 7 | Canada | HIC | 1 |
| 8 | Chile | HIC | 1 |
| 9 | China | UMIC | 202 |
| 10 | Croatia | HIC | 1 |
| 11 | Denmark | HIC | 1 |
| 12 | Egypt | LMIC | 1 |
| 13 | Estonia | HIC | 1 |
| 14 | France | HIC | 13 |
| 15 | Germany | HIC | 3 |
| 16 | Greece | HIC | 1 |
| 17 | Hungary | HIC | 1 |
| 18 | India | LMIC | 7 |
| 19 | Indonesia | UMIC | 1 |
| 20 | Iran | UMIC | 9 |
| 21 | Iraq | UMIC | 2 |
| 22 | Ireland | HIC | 1 |
| 23 | Israel | HIC | 1 |
| 24 | Italy | HIC | 16 |
| 25 | Japan | HIC | 5 |
| 26 | Jordan | UMIC | 2 |
| 27 | Kazakhstan | UMIC | 1 |
| 28 | Lithuania | HIC | 1 |
| 29 | Madagascar | LIC | 1 |
| 30 | Moldova | LMIC | 1 |
| 31 | Morocco | LMIC | 1 |
| 32 | Netherlands | HIC | 1 |
| 33 | Nigeria | LMIC | 1 |
| 34 | Norway | HIC | 1 |
| 35 | Oman | HIC | 1 |
| 36 | Pakistan | LMIC | 2 |
| 37 | Portugal | HIC | 1 |
| 38 | Romania | HIC | 1 |
| 39 | Russia | UMIC | 1 |
| 40 | Saudi Arabia | HIC | 4 |
| 41 | Scotland | PART OF UK - IGNORE | 1 |
| 42 | Singapore | HIC | 1 |
| 43 | Slovakia | HIC | 1 |
| 44 | Slovenia | HIC | 1 |
| 45 | South Korea | HIC | 6 |
| 46 | Spain | HIC | 11 |
| 47 | Sweden | HIC | 1 |
| 48 | Switzerland | HIC | 2 |
| 49 | Tunisia | LMIC | 1 |
| 50 | Turkey | UMIC | 7 |
| 51 | UAE | HIC | 1 |
| 52 | UK | HIC | 5 |
| 53 | Ukraine | LMIC | 1 |
| 54 | USA | HIC | 28 |
| 55 | Uzbekistan | LMIC | 1 |

**Abbreviations:** HIC- High Income Countries; LMIC- Low Middle Income Countries; UMIC- Upper Middle-Income Countries; LIC- Low Income Countries

**Table 2:** Baseline characteristics of studies included in the systematic review and meta-analysis

| **S. No** | **Author & Year** | **Country** | **Sample size** | **Mean Age, SD** | **Males** |
| --- | --- | --- | --- | --- | --- |
| 1 | Mao L, 2020 ^1^ | China | 214 | 52.7, 15.5 | 87 |
| 2 | Helms J (a), 2020 ^2^ | France | 58 | Median (63) | NA |
| 3 | Hornuss D, 2020 ^3^ | Germany | 45 | 56, 16.9 | 25 |
| 4 | Levinson R, 2020 ^4^ | Israel | 42 | 43.67, 15.43 | 23 |
| 5 | Yan CH, 2020 ^5^ | USA | 59 | NA | 29 |
| 6 | Chen N, 2020 ^6^ | China | 99 | 55.5, 13.1 | 67 |
| 7 | Guan W, 2020 ^7^ | China | 1099 | 46.67, 17.07 | 637 |
| 8 | Chen P, 2020 ^8^ | China | 136 | 48.33, 17.98 | 69 |
| 9 | Chen Q, 2020 ^9^ | China | 145 | 47.5, 14.6 | 79 |
| 10 | Chen T, 2020 ^10^ | China | 274 | 58.67, 19.38 | 171 |
| 11 | Chen X, 2020 ^11^ | China | 267 | 47.67, 22.36 | 121 |
| 12 | Cummings NJ, 2020 ^12^ | USA | 257 | 61.67, 15.65 | 170 |
| 13 | Du W, 2020 ^13^ | China | 53 | 42.49, 35.53 | 26 |
| 14 | Jin X, 2020 ^14^ | China | 651 | 45.21, 14.42 | 331 |
| 15 | Kim ES, 2020 ^15^ | South Korea | 28 | 42.6, 13.4 | 15 |
| 16 | Liang WH, 2020 ^16^ | China | 1590 | 48.9, 16.3 | 904 |
| 17 | Liu K, 2020 ^17^ | China | 137 | 53.33, 47.2 | 61 |
| 18 | Ma Y, 2020 ^18^ | China | 47 | 35.67, 54.3 | 20 |
| 19 | Niu S, 2020 ^19^ | China | 141 | NA | 70 |
| 20 | Shen L, 2020 ^20^ | China | 119 | 49.33, 17.26 | 56 |
| 21 | Sun Y, 2020 ^21^ | China | 63 | 45, 62.21 | NA |
| 22 | Tian S, 2020 ^22^ | China | 262 | 47.5, 69.32 | 127 |
| 23 | Tomlins J, 2020 ^23^ | UK | 95 | 72, 17.31 | 60 |
| 24 | Wang D (a), 2020 ^24^ | China | 138 | 55.33, 19.48 | 75 |
| 25 | Wang D (b), 2020 ^25^ | China | 107 | 50.67, 21.79 | 57 |
| 26 | Wang L (a), 2020 ^26^ | China | 26 | 42.93, 15.53 | 11 |
| 27 | Romero-Sanchez C, 2020 ^27^ | Spain | 841 | 66.42, 14.96 | 473 |
| 28 | Wang X (a), 2020 ^28^ | China | 80 | 39.83, 12.46 | 31 |
| 29 | Wang X (b), 2020 ^29^ | China | 1012 | 49, 14.1 | 524 |
| 30 | Xu YH, 2020 ^30^ | China | 50 | 43.9, 16.8 | 29 |
| 31 | Yang W, 2020 ^31^ | China | 149 | 45.11, 13.35 | 81 |
| 32 | Yang X, 2020 ^32^ | China | 52 | 59.7, 13.3 | 35 |
| 33 | Yin S, 2020 ^33^ | China | 33 | 47.5, 25.96 | 16 |
| 34 | Huang C, 2020 ^34^ | China | 41 | 49.33, 13.06 | 30 |
| 35 | Zhang G, 2020 ^35^ | China | 221 | 53.5, 20.52 | 108 |
| 36 | Zhao XY, 2020 ^36^ | China | 91 | Median (46) | 49 |
| 37 | Zheng Y (a), 2020 ^37^ | China | 73 | 46.67, 41.59 | 40 |
| 38 | Wan S, 2020 ^38^ | China | 135 | 46, 14.24 | 72 |
| 39 | Qian GQ, 2020 ^39^ | China | 91 | 47.83, 15.44 | 37 |
| 40 | Hu Z, 2020 ^40^ | China | 24 | 36.17, 29.95 | 8 |
| 41 | Wu J (a), 2020 ^41^ | China | 80 | 46.1, 15.42 | 39 |
| 42 | Xu XW, 2020 ^42^ | China | 62 | 41.67, 15.18 | 35 |
| 43 | Zhang X, 2020 ^43^ | China | 645 | 45.34, 13.86 | 328 |
| 44 | Wang L(b), 2020 ^44^ | China | 339 | 70, 8.19 | 166 |
| 45 | Cai Q, 2020 ^45^ | China | 298 | 47.17, 20.86 | 145 |
| 46 | Kong I, 2020 ^46^ | South Korea | 28 | 45.2, 41.4 | 15 |
| 47 | Gupta N, 2020 ^47^ | India | 21 | 43.1, 45.32 | 14 |
| 48 | Karadas O, 2020 ^48^ | Turkey | 239 | 46.46, 15.41 | 133 |
| 49 | Beltran-Corbellini A, 2020 ^49^ | Spain | 79 | 61.6, 17.4 | 48 |
| 50 | Lechien JR (a), 2020 ^50^ | France/ Belgium | 417 | 36.9, 11.4 | 154 |
| 51 | Han R, 2020 ^51^ | China | 108 | 52, 51.84 | 38 |
| 52 | Lo IL, 2020 ^52^ | China | 10 | 48.33, 31.82 | 3 |
| 53 | Du Y, 2020 ^53^ | China | 85 | 65.8, 14.2 | 62 |
| 54 | Han X, 2020 ^54^ | China | 17 | 40, 10 | 6 |
| 55 | Lian J (a), 2020 ^55^ | China | 788 | NA | 407 |
| 56 | Shi S, 2020 ^56^ | China | 416 | 60, 55.05 | 205 |
| 57 | Lei P, 2020 ^57^ | China | 14 | 47, 19 | 8 |
| 58 | Zheng F, 2020 ^58^ | China | 161 | 45.17, 17.58 | 80 |
| 59 | Wang Z, 2020 ^59^ | China | 69 | 46.33, 20.44 | 32 |
| 60 | Bhatraju PK, 2020 ^60^ | USA | 24 | 64, 18 | 15 |
| 61 | Chang D, 2020 ^61^ | China | 13 | 38.67, 11.63 | 10 |
| 62 | Xu X (a), 2020 ^62^ | China | 90 | 51.33, 51.23 | 39 |
| 63 | Liu Y (a), 2020 ^63^ | China | 12 | 53.67, 18.01 | 8 |
| 64 | Zhou F, 2020 ^64^ | China | 191 | 56.33, 15.69 | 119 |
| 65 | Han Y, 2020 ^65^ | China | 25 | 45.33, 37.73 | 12 |
| 66 | Cao J, 2020 ^66^ | China | 102 | 52.67, 22.56 | 53 |
| 67 | Zhao W, 2020 ^67^ | China | 101 | 45.48, 43.62 | 56 |
| 68 | Li X (a), 2020 ^68^ | China | 131 | 47, 15 | 63 |
| 69 | Feng Y, 2020 ^69^ | China | 476 | 52.33, 17.84 | 271 |
| 70 | Lei Z, 2020 ^70^ | China | 20 | 43.2, 14 | 10 |
| 71 | Zhang R, 2020 ^71^ | China | 120 | 45.4, 15.6 | 43 |
| 72 | Barrasa H, 2020 ^72^ | Spain | 48 | 63, 12 | 27 |
| 73 | Li YK, 2020 ^73^ | China | 25 | 60.33, 14.15 | 12 |
| 74 | Chen TL, 2020 ^74^ | China | 203 | 54.33, 20.16 | 108 |
| 75 | Chu J, 2020 ^75^ | China | 54 | 46, 35.8 | 36 |
| 76 | Cheng Z, 2020 ^76^ | China | 11 | 50.36, 15.5 | 8 |
| 77 | Goyal P, 2020 ^77^ | USA | 393 | 61.5, 18.67 | 238 |
| 78 | Chen J (a), 2020 ^78^ | China | 249 | 50.33, 20.88 | 126 |
| 79 | Escalera-Antezana JP, 2020 ^79^ | Bolivia | 12 | 36.5, 15.78 | 6 |
| 80 | Merza MA, 2020 ^80^ | Iraq | 15 | 28.06, 16.42 | 9 |
| 81 | Wang R, 2020 ^81^ | China | 125 | 38.76, 13.79 | 71 |
| 82 | Zheng Y (b), 2020 ^82^ | China | 99 | 49.39, 18.45 | 51 |
| 83 | Garg R, 2020 ^83^ | India | 391 | 49.07, 15.74 | 241 |
| 84 | Li Y, 2020 ^84^ | China | 219 | 53.3, 15.9 | 89 |
| 85 | Chung M, 2020 ^85^ | China | 21 | 51, 14.5 | 13 |
| 86 | Du RH, 2020 ^86^ | China | 179 | 57.6, 13.7 | 97 |
| 87 | Li X (b), 2020 ^87^ | China | 548 | 59, 15.61 | 279 |
| 88 | Pan F, 2020 ^88^ | China | 21 | 40, 9 | 6 |
| 89 | Shi H, 2020 ^89^ | China | 81 | 49.5, 11 | 42 |
| 90 | Song F, 2020 ^90^ | China | 51 | 49, 16 | 25 |
| 91 | Wu J (b), 2020 ^91^ | China | 80 | 44, 11 | 42 |
| 92 | Zhang JJ, 2020 ^92^ | China | 140 | 56.33, 46.44 | 7 |
| 93 | Duanmu Y, 2020 ^93^ | USA | 100 | 47.33, 24.82 | 56 |
| 94 | Giacomelli A, 2020 ^94^ | Italy | 59 | 61.33, 18.24 | 40 |
| 95 | Li K, 2020 ^95^ | China | 83 | 45.5, 12.3 | 44 |
| 96 | Jain R, 2020 ^96^ | USA | 3218 | NA | NA |
| 97 | Qin C, 2020 ^97^ | China | 1875 | 61.33, 14.09 | 945 |
| 98 | Merkler AE, 2020 ^98^ | USA | 1916 | 63.67, 18.55 | 1111 |
| 99 | Radmanesh A, 2020 ^99^ | USA | 242 | 68.7, 16.5 | 150 |
| 100 | Xiong W, 2020 ^100^ | China | 917 | 48.7, 17.1 | 506 |
| 101 | Mahammedi A, 2020 ^101^ | Italy | 108 | 69, 15 | 69 |
| 102 | Agarwal P, 2020 ^102^ | USA | 404 | NA | 197 |
| 103 | Notz Q, 2020 ^103^ | Germany | 38 | 64.5, 19.5 | 25 |
| 104 | Helms J (b), 2020 ^104^ | France | 140 | 61.33, 13.48 | 100 |
| 105 | Ora J, 2020 ^105^ | Italy | 21 | 68.4, 13.9 | 13 |
| 106 | Duployez N, 2020 ^106^ | France | 122 | 60.67, 54.77 | 93 |
| 107 | Fu J, 2020 ^107^ | China | 75 | 46.6, 14 | 45 |
| 108 | Gao Y, 2020 ^108^ | China | 44 | 52.17, 19.54 | 15 |
| 109 | Gu Q, 2020 ^109^ | China | 50 | 44.84, 16.26 | 25 |
| 110 | Guo T, 2020 ^110^ | China | 98 | 49, 12 | 43 |
| 111 | Hafiz M, 2020 ^111^ | Indonesia | 30 | 53.9, 16.4 | 16 |
| 112 | He F, 2020 ^112^ | China | 288 | 48.27, 20.64 | 131 |
| 113 | Hu HT, 2020 ^113^ | China | 55 | 63.67, 15.22 | 34 |
| 114 | Huang R, 2020 ^114^ | China | 280 | 43.67, 17.88 | 146 |
| 115 | Jiang H, 2020 ^115^ | China | 59 | 64, 12.16 | 29 |
| 116 | Kato H, 2020 ^116^ | Japan | 70 | 66.67, 6.81 | 47 |
| 117 | Khan M, 2020 ^117^ | Pakistan | 121 | 47, 64.52 | 85 |
| 118 | Klopfenstein T, 2020 ^118^ | France | 70 | 57, 19 | 29 |
| 119 | Lapostolle F, 2020 ^119^ | France | 1452 | 44.33, 18.55 | 700 |
| 120 | Liu D (a), 2020 ^120^ | China | 2044 | 61, 14.09 | 1000 |
| 121 | Liu J, 2020 ^121^ | China | 1190 | 57, 14.84 | 635 |
| 122 | Mishra P, 2020 ^122^ | India | 74 | NA | 43 |
| 123 | Qu J, 2020 ^123^ | China | 246 | 53.63, 15.27 | 115 |
| 124 | Ren D, 2020 ^124^ | China | 150 | 51.33, 19.46 | 82 |
| 125 | Trigo J, 2020 ^125^ | Spain | 576 | 67.2, 14.7 | 326 |
| 126 | Wang Y, 2020 ^126^ | China | 90 | 45, 14 | 33 |
| 127 | Wang ZH, 2020 ^127^ | China | 59 | 67.4, 11.3 | 38 |
| 128 | Wei Y, 2020 ^128^ | China | 276 | 50, 12.67 | 155 |
| 129 | Wu G, 2020 ^129^ | China | 299 | NA | 137 |
| 130 | Xie Y, 2020 ^130^ | China | 62 | 64.1, 14.95 | 27 |
| 131 | Xiong Y, 2020 ^131^ | China | 86 | NA | 43 |
| 132 | Xu X (b), 2020 ^132^ | China | 88 | 57.11, 15.39 | 36 |
| 133 | Yang L, 2020 ^133^ | China | 200 | 55, 17.1 | 98 |
| 134 | Yang Q, 2020 ^134^ | China | 136 | 54.67, 14.98 | 66 |
| 135 | Zhao D, 2020 ^135^ | China | 19 | 46.67, 23.23 | 11 |
| 136 | Zhao G, 2020 ^136^ | China | 47 | 50, 21.41 | 19 |
| 137 | Zheng Y (c), 2020 ^137^ | China | 238 | NA | 141 |
| 138 | Guilmot A, 2020 ^138^ | Belgium | 15 | 61, 38.44 | 12 |
| 139 | Liguori C, 2020 ^139^ | Italy | 103 | 55, 14.65 | 59 |
| 140 | Romagnolo A, 2020 ^140^ | Italy | 344 | 61.5, 17.8 | 204 |
| 141 | Asai N, 2020 ^141^ | Japan | 92 | 58, 57.24 | 50 |
| 142 | Cavalleri M, 2020 ^142^ | Italy | 172 | 64.2, 13.4 | 117 |
| 143 | Davarpanah AH, 2020 ^143^ | Iran | 228 | 56.1, 15.9 | 148 |
| 144 | Faury H, 2020 ^144^ | France | 100 | 61, 17.3 | 58 |
| 145 | Feng G, 2020 ^145^ | China | 134 | 44.67, 17.24 | 65 |
| 146 | Kang MK, 2020 ^146^ | South Korea | 118 | 60.33, 15.01 | 52 |
| 147 | Liu D (b), 2020 ^147^ | China | 599 | 52.67, 67.62 | 325 |
| 148 | Marvisi M, 2020 ^148^ | Italy | 90 | 66, 15 | 60 |
| 149 | Meiler S, 2020 ^149^ | Germany | 64 | 57.2, 14.5 | 40 |
| 150 | Salepci E, 2020 ^150^ | Turkey | 223 | 49.67, 19.4 | 113 |
| 151 | Shah SJ, 2020 ^151^ | USA | 33 | 62.67, 19.37 | 21 |
| 152 | Wang W (a), 2020 ^152^ | China | 123 | 67.5, 16.13 | 60 |
| 153 | Xiang C, 2020 ^153^ | China | 53 | 53, 16 | 31 |
| 154 | Zandkarimi E, 2020 ^154^ | Iran | 1831 | 52.74, 22.16 | 1019 |
| 155 | Zhang J, 2020 ^155^ | China | 211 | 51.67, 16.42 | 95 |
| 156 | Zhou J, 2020 ^156^ | China | 201 | 45.93, 18.82 | 102 |
| 157 | Abbas HM, 2020 ^157^ | Iraq | 284 | 48.6, 14.79 | 188 |
| 158 | Campioli CC, 2020 ^158^ | USA | 251 | 53, 27 | 103 |
| 159 | Chen X (b), 2020 ^159^ | China | 70 | 42.93, 13.32 | 41 |
| 160 | Chon YJ, 2020 ^160^ | South Korea | 281 | 61.17, 16.39 | 75 |
| 161 | Coppola A, 2020 ^161^ | Italy | 73 | 69.75 | 52 |
| 162 | Gatti M, 2020 ^162^ | Italy | 260 | 62.8, 15.8 | 159 |
| 163 | Gu J, 2020 ^163^ | China | 155 | 42.97, 14.85 | 87 |
| 164 | Kuzan TY, 2020 ^164^ | Turkey | 120 | 50.9, 16.04 | 67 |
| 165 | Li J (a), 2020 ^165^ | China | 74 | 64.33, 12.85 | 44 |
| 166 | Li J (b), 2020 ^166^ | China | 94 | 58.7, 16 | 51 |
| 167 | Lian J (b), 2020 ^167^ | China | 465 | 46, 61.72 | 243 |
| 168 | Liu Y (b), 2020 ^168^ | China | 107 | 54.83, 16.16 | 45 |
| 169 | Moon SS, 2020 ^169^ | South Korea | 352 | 55.67, 25.31 | 144 |
| 170 | Tabata S, 2020 ^170^ | Japan | 104 | 63.33, 21.05 | 54 |
| 171 | Tan JY, 2020 ^171^ | Singapore | 287 | NA | 233 |
| 172 | Wang H, 2020 ^172^ | China | 13 | 49.54, 17.42 | 7 |
| 173 | Wang W (b), 2020 ^173^ | China | 421 | 50.67, 16.36 | 214 |
| 174 | Wang X (c), 2020 ^174^ | China | 131 | 49, 19.49 | 59 |
| 175 | Xu H, 2020 ^175^ | China | 102 | NA | 50 |
| 176 | Ye H, 2020 ^176^ | China | 117 | 48.2, 13.5 | 65 |
| 177 | Zha L, 2020 ^177^ | China | 88 | 56.17, 13.94 | 45 |
| 178 | Zhang L, 2020 ^178^ | China | 134 | 60.78, 12.98 | 87 |
| 179 | Zhou H, 2020 ^179^ | China | 42 | 38.93, 11.67 | 17 |
| 180 | Zhuang Y, 2020 ^180^ | China | 22 | 40.7, 10.3 | 11 |
| 181 | Lechien JR (b), 2020 ^181^ | France/ Belgium | 1420 | 39.17, 12.09 | 458 |
| 182 | Rothstein A, 2020 ^182^ | USA | 844 | 59, 18 | 405 |
| 183 | Chary E, 2020 ^183^ | France | 115 | 50, 47.3 | 34 |
| 184 | Scullen T, 2020 ^184^ | USA | 76 | 61.4, 11.7 | 39 |
| 185 | Aggarwal A, 2020 ^185^ | India | 32 | 53.58, 10.67 | 19 |
| 186 | Alsofayan YM, 2020 ^186^ | Saudi Arabia | 1519 | 36 | 825 |
| 187 | Carignan A, 2020 ^187^ | Canada | 134 | 54.27, 17.46 | 64 |
| 188 | Chen FF, 2020 ^188^ | China | 681 | 63.67, 13.37 | 362 |
| 189 | Chen J (c), 2020 ^189^ | China | 3309 | 60, 14.83 | 1642 |
| 190 | Chen Y, 2020 ^190^ | China | 904 | 54, 20.79 | 421 |
| 191 | Duan X, 2020 ^191^ | China | 25 | 52, 19.3 | 15 |
| 192 | Easom N, 2020 ^192^ | UK | 68 | 39.67, 57.18 | 32 |
| 193 | Fu Y, 2020 ^193^ | China | 482 | 54.27, 21.04 | 243 |
| 194 | Gao X, 2020 ^194^ | China | 36 | 44.1, 17.6 | 17 |
| 195 | Gaur A, 2020 ^195^ | India | 26 | 41.3, 14.14 | 16 |
| 196 | Gupta S, 2020 ^196^ | USA | 2215 | 60.5, 14.5 | 1436 |
| 197 | Jalessi M, 2020 ^197^ | Iran | 92 | 52.94, 13.25 | 62 |
| 198 | Li G, 2020 ^198^ | China | 199 | 62.67, 18.67 | 89 |
| 199 | Li W, 2020 ^199^ | China | 97 | 45, 18 | 45 |
| 200 | Martin-Sanchez FJ, 2020 ^200^ | Spain | 1379 | 62, 18 | 739 |
| 201 | Meng H, 2020 ^201^ | China | 58 | 42.6, 16.56 | 26 |
| 202 | O'Reilly GM (a), 2020 ^202^ | Australia | 14 | 50, 18 | 11 |
| 203 | Samrah SM, 2020 ^203^ | Jordan | 81 | 39.95, 16.59 | 37 |
| 204 | Khamis F, 2020 ^204^ | Oman | 63 | 48, 16 | 53 |
| 205 | Saleemi S, 2020 ^205^ | Saudi Arabia | 51 | 48.33, 27.46 | 23 |
| 206 | Shang Y, 2020 ^206^ | China | 307 | 44.67, 16.39 | 164 |
| 207 | Sun D, 2020 ^207^ | China | 84 | 45.33, 12.07 | 37 |
| 208 | Wang D (c), 2020 ^208^ | China | 143 | 54.67, 20.97 | 73 |
| 209 | Wang F, 2020 ^209^ | China | 323 | 46, 19.36 | 154 |
| 210 | Wang S, 2020 ^210^ | China | 605 | 58, 15.6 | 322 |
| 211 | Yang J, 2020 ^211^ | China | 37 | 32.32, 15.71 | 24 |
| 212 | Yue H, 2020 ^212^ | China | 86 | 42.1, 17.57 | 38 |
| 213 | Zhang SY, 2020 ^213^ | China | 788 | NA | 407 |
| 214 | Zhang Y, 2020 ^214^ | China | 258 | 63.33, 10.44 | 138 |
| 215 | Zhao K, 2020 ^215^ | China | 619 | 58.2, 13.1 | 309 |
| 216 | Fan S, 2020 ^216^ | China | 86 | 66.6, 11.1 | 54 |
| 217 | Hernández-Fernández F, 2020 ^217^ | Spain | 23 | 66.9, 11.8 | 18 |
| 218 | García-Azorín D, 2020 ^218^ | Spain | 576 | 67.18, 14.75 | 326 |
| 219 | Liu M, 2020 ^219^ | China | 30 | 35, 8 | 10 |
| 220 | Qin W, 2020 ^220^ | China | 582 | 63, 12.63 | 293 |
| 221 | Sun C, 2020 ^221^ | China | 150 | 45, 16 | 67 |
| 222 | Teich VD, 2020 ^222^ | Brazil | 510 | 39.9, 13.6 | 290 |
| 223 | Xie W, 2020 ^223^ | China | 75 | 58.1, 15.9 | 41 |
| 224 | Xu M, 2020 ^224^ | China | 23 | 46.17, 9.09 | 15 |
| 225 | Zeng YL, 2020 ^225^ | China | 49 | 52.82, 18.12 | 26 |
| 226 | Zhang MQ, 2020 ^226^ | China | 9 | 33, 28.86 | 5 |
| 227 | Chen L, 2020 ^227^ | China | 29 | 53.67, 41.33 | 21 |
| 228 | Liu C, 2020 ^228^ | China | 278 | 48.1, 17 | 130 |
| 229 | Lu XF, 2020 ^229^ | China | 141 | 48.33, 58.42 | 77 |
| 230 | Zhu Q, 2020 ^230^ | China | 64 | 67.33, 7.58 | 36 |
| 231 | Zou W, 2020 ^231^ | China | 63 | 46.71, 16 | 32 |
| 232 | Casas-Rojo JM, 2020 ^232^ | Spain | 15111 | 63.13, 62.27 | 8643 |
| 233 | Fakiri K El, 2020 ^233^ | Morocco | 74 | 8.06, 12.72 | 34 |
| 234 | Gotzinger F, 2020 ^234^ | Multicentric | 582 | 5.83, 8.55 | 311 |
| 235 | Guo CX, 2020 ^235^ | China | 341 | 7, 10.41 | 183 |
| 236 | Korkmaz MF, 2020 ^236^ | Turkey | 81 | 9.25, 9 | 48 |
| 237 | Mamishi S, 2020 ^237^ | Iran | 24 | 6.33, 4.73 | 11 |
| 238 | Qui H, 2020 ^238^ | China | 36 | 8.3, 3.5 | 23 |
| 239 | Romani L, 2020 ^239^ | Italy | 43 | 8.01, 13.02 | 24 |
| 240 | Song X, 2020 ^240^ | USA | 54 | 10.99, 17.62 | 25 |
| 241 | Sun D, 2020 ^241^ | China | 8 | 6.62, 6.34 | 6 |
| 242 | Swann OV, 2020 ^242^ | UK | 651 | 6.2, 9.95 | 367 |
| 243 | Zhao Y, 2020 ^243^ | China | 23 | 5.7, 3.8 | 9 |
| 244 | Dang JZ, 2020 ^244^ | China | 17 | 88.2, 2.75 | 12 |
| 245 | Knopp P, 2020 ^245^ | UK | 217 | 80, 6.8 | 134 |
| 246 | Li P, 2020 ^246^ | China | 204 | 74.33, 26.13 | 100 |
| 247 | Al-Mutair A, 2020 ^247^ | Saudi Arabia | 401 | 38.16 (13.43) | 321 |
| 248 | Almalki ZS, 2020 ^248^ | Saudi Arabia | 458 | 38.82 (12.86) | 398 |
| 249 | An P, 2020 ^249^ | China | 205 | 54 (22-77) | 122 |
| 250 | Ayaz A, 2020 ^250^ | Pakistan | 66 | 50.6 (19.1) | 40 |
| 251 | Baghaei P, 2020 ^251^ | Iran | 127 | 55 (45-63) | 97 |
| 252 | Bouzid D, 2020 ^252^ | France | 268 | 59 (49-73) | 190 |
| 253 | Buckner FS, 2020 ^253^ | USA | 105 | 69 (23-97) | 53 |
| 254 | Caronna E, 2020 ^254^ | Spain | 130 | 53.9 (16.4) | 64 |
| 255 | Chen HJ, 2020 ^255^ | China | 34 | 54.5 (11.8) | 21 |
| 256 | Chen Y (b), 2020 ^256^ | China | 208 | 64 (55-69) | 101 |
| 257 | Chi Q, 2020 ^257^ | China | 17 | 53.5 (13.4) | 9 |
| 258 | Cobb NL, 2020 ^258^ | USA | 65 | 60.4 (15.7) | 46 |
| 259 | Deng LS, 2020 ^259^ | China | 83 | 53 (3-80) | 35 |
| 260 | Du H, 2020 ^260^ | China | 164 | 61.8 (13.6) | 84 |
| 261 | Emami A, 2020 ^261^ | Iran | 6147 |  |  |
| 262 | Fu F, 2020 ^262^ | China | 55 | 45 (20-67) | 33 |
| 263 | Gao J, 2020 ^263^ | China | 96 | 43 (40-46.6) | 44 |
| 264 | He S, 2020 ^264^ | China | 420 | 56 (43-63.75) | 207 |
| 265 | Higuchi T, 2020 ^265^ | Japan | 57 | 52 (35-69.5) | 32 |
| 266 | Homayounieh F, 2020 ^266^ | Iran | 90 | 59.58 (15.98) | 58 |
| 267 | Hong JM, 2020 ^267^ | China | 41 | 39.9 (15-83) |  |
| 268 | Hong L, 2020 ^268^ | China | 67 | 45 (15.2) | 36 |
| 269 | Ibrahim OR, 2020 ^269^ | Nigeria | 45 | 43 (16) | 39 |
| 270 | Jiang J, 2020 ^270^ | China | 36 | 41 (30-55) | 18 |
| 271 | Jin XH, 2020 ^271^ | China | 146 | 47 (4-86) | 77 |
| 272 | Jourdes A, 2020 ^272^ | France | 263 | 65 (54-76) | 155 |
| 273 | Kadiane-Oussou NJ, 2020 ^273^ | France | 114 | 56.7 (20) | 48 |
| 274 | Kaeuffer C, 2020 ^274^ | France | 1045 | 66.3 (16) | 612 |
| 275 | Khraise NW, 2020 ^275^ | Jordan | 108 | 36.4 (17.1) | 45 |
| 276 | Kim KH, 2020 ^276^ | Uzbekistan | 1030 | 36 (26-47) | 588 |
| 277 | Kumar A, 2020 ^277^ | India | 91 | 44 (15-82) | 61 |
| 278 | Li X (c), 2020 ^278^ | China | 52 | 36 (19-66), 6.33 (0-15) | 28 |
| 279 | Lima MA, 2020 ^279^ | Brazil | 57 | 41.4 (10.4) | 26 |
| 280 | Lin L, 2020 ^280^ | China | 78 | 66.5 (53.75-70) | 38 |
| 281 | Liu H, 2020 ^281^ | China | 40 | 40 (30-47) | 7 |
| 282 | Liu J (b), 2020 ^282^ | China | 214 | 68 (59-76) | 119 |
| 283 | Liu M, 2020 ^283^ | China | 122 | 48 (15) | 61 |
| 284 | Liu W, 2020 ^284^ | China | 140 | 36 (30.75-43.25) | 44 |
| 285 | de Magalhaes JJF, 2020 ^285^ | Brazil | 557 | 47 (32-68) | 253 |
| 286 | Mirfazeli FS, 2020 ^286^ | Iran | 201 | 51.84 (16.57) | 119 |
| 287 | Morikawa M, 2020 ^287^ | Japan | 154 | 44 (32-56) | 94 |
| 288 | Novelli L, 2020 ^288^ | Italy | 508 | 66.3 (15.8) | 368 |
| 289 | Pugliese L, 2020 ^289^ | Italy | 92 | 63 (52.3-78.5) | 66 |
| 290 | Qui Z, 2020 ^290^ | China | 64 | 48.5 (10-77) | 38 |
| 291 | Raberahona M, 2020 ^291^ | Madagascar | 1288 | 38 (26-52) | 626 |
| 292 | Ren C, 2020 ^292^ | China | 422 | 47 (33-60) | 201 |
| 293 | Shi JC, 2020 ^293^ | China | 105 | 42 (22-64) | 57 |
| 294 | Sun JT, 2020 ^294^ | China | 99 | 52 (42-62), 70.5 (61.25-80.75) | 60 |
| 295 | Sun L, 2020 ^295^ | China | 55 | 44 (34-56) | 31 |
| 296 | Tian J, 2020 ^296^ | China | 120 | 52.3 (17.4) | 60 |
| 297 | Vial MR, 2020 ^297^ | Chile | 381 | 39 (31-49) | 183 |
| 298 | Vilke GM, 2020 ^298^ | USA | 330 |  |  |
| 299 | Wang C, 2020 ^299^ | China | 169 | 45 (34.5-55) | 86 |
| 300 | Wang J (a), 2020 ^300^ | China | 297 | 38 (31.25-54), 48 (36.5-57), 47 (34.25-57.75) | 164 |
| 301 | Wang J (b), 2020 ^301^ | China | 342 | 44.5 (32-57) | 112 |
| 302 | Wang Z (b), 2020 ^302^ | China | 293 | 59.2 (42.8-73.1) | 138 |
| 303 | Wei Y (b), 2020 ^303^ | China | 628 | 53 (14.8) | 296 |
| 304 | Wu B, 2020 ^304^ | China | 91 | 50 (14) | 52 |
| 305 | Xiao J, 2020 ^305^ | China | 243 | 47 (20-89) | 105 |
| 306 | Xie L, 2020 ^306^ | China | 49 | 41 (33-56) | 26 |
| 307 | Zayet S, 2020 ^307^ | France | 70 | 56.7 (19.3) | 29 |
| 308 | Zhang L (b), 2020 ^308^ | China | 34 | 49.21 (7-88) | 19 |
| 309 | Zhang N, 2020 ^309^ | China | 60 | 64.4 (11) | 43 |
| 310 | Bach I, 2020 ^310^ | USA | 683 | 61.13 (15.39) | 397 |
| 311 | Battaglini D, 2020 ^311^ | Italy | 94 | 61.6 (11.1) | 74 |
| 312 | Cantador E, 2020 ^312^ | Spain | 1419 |  |  |
| 313 | Chachkhiani D, 2020 ^313^ | USA | 250 | 60 (15) | 113 |
| 314 | Foresti C, 2020 ^314^ | Italy | 1832 |  |  |
| 315 | Frontera JA, 2020 ^315^ | USA | 4491 | 70.39 (14.29) | 2607 |
| 316 | John S, 2020 ^316^ | UAE | 591 |  |  |
| 317 | Kacem I, 2020 ^317^ | Tunisia | 646 | 42.17 (17.58) | 348 |
| 318 | Keller E, 2020 ^318^ | Switzerland | 32 |  |  |
| 319 | Lang M, 2020 ^319^ | USA | 468 |  |  |
| 320 | Lin E, 2020 ^320^ | USA | 2054 | 64 (50-75) | 1171 |
| 321 | Liotta EM, 2020 ^321^ | USA | 509 | 58.5 (16.9) | 281 |
| 322 | Requena M, 2020 ^322^ | Spain | 2050 |  |  |
| 323 | Salahuddin H, 2020 ^323^ | USA | 574 | 62.8 (17.6) | 276 |
| 324 | Shekhar R, 2020 ^324^ | USA | 90 |  |  |
| 325 | Siegler JE, 2020 ^325^ | USA, Spain, Egypt, Romania | 14483 |  |  |
| 326 | Yao X, 2020 ^326^ | China | 2474 | 61 (15.7) | 1235 |
| 327 | Wang YX, 2020 ^327^ | China | 90 | 53.9 (16.92) | 54 |
| 328 | O'Reilly GM (b), 2020 ^328^ | Australia | 255 | 58 (22) | 130 |
| 329 | Bellan M, 2020 ^329^ | Italy | 407 | 71 (58-80) | 239 |
| 330 | Chen SL, 2020 ^330^ | China | 1168 | 43.5 (32-57) | 560 |
| 331 | Horvath L, 2020 ^331^ | Australia | 102 | 45 (17-87) |  |
| 332 | Huang H, 2020 ^332^ | China | 125 | 44.87 (18.55) | 63 |
| 333 | Rokni M, 2020 ^333^ | Iran | 233 | 49.8 | 149 |
| 334 | Shah NN, 2020 ^334^ | India | 655 | 32.7 (10.1) | 414 |
| 335 | Bayesheva D, 2020 ^335^ | Kazakhstan | 650 | 7.1 (6.1) | 366 |
| 336 | Bhumbra S, 2020 ^336^ | USA | 19 | 5 (0.8-16) | 14 |
| 337 | Cai J, 2020 ^337^ | China | 49 | 11.5 (5.12) | 28 |
| 338 | Chen J, 2020 ^338^ | China | 12 | 14.5 (9.25-15.75) | 6 |
| 339 | Duramaz BB, 2020 ^339^ | Turkey | 43 | - | 17 |
| 340 | Li K, 2020 ^340^ | China | 72 | 2 (0-11) |  |
| 341 | Parri N, 2020 ^341^ | Italy | 170 | 3.75 (0.33-10.7) | 95 |
| 342 | Yilmaz K, 2020 ^342^ | Turkey | 105 | 9.05 (5.47) | 54 |
| 343 | BaĞ SoytaŞ R, 2020 ^343^ | Turkey | 218 | 75.3 (70-81) | 112 |
| 344 | Davis P, 2020 ^344^ | Scotland | 222 | 82 (56-99) | 74 |
| 345 | Gan JM, 2020 ^345^ | UK | 122 | 81 (8) | 51 |
| 346 | Hwang J, 2020 ^346^ | South Korea | 340 | 77.32 (7.25), 74.39 (6.70) | 129 |
| 347 | Kennedy M, 2020 ^347^ | USA | 817 | 77.7 (8.2) | 386 |
| 348 | Mendes A, 2020 ^348^ | Switzerland | 235 | 86.3 (6.5) | 102 |
| 349 | Vrillon A, 2020 ^349^ | France | 76 | 90 (86-92) | 34 |
| 350 | Zhou J, 2020 ^350^ | China | 118 | 70.6 (6.9), 73.1 (7.3) | 53 |

**Abbreviations:** NA- Not available; USA- United States of America; UK- United Kingdom

**Table 3:** Benjamini-Hochberg procedure for controlling the false discovery rate

| **Association of severity of disease and mortality with neurological manifestations** | **P-values** | **Benjamini-Hochberg significance** | **Adjusted Benjamini-Hochberg P-value** |
| --- | --- | --- | --- |
| Skeletal Muscle Injury & severity of COVID-19 | 0.0001 | significant | 0.00115 |
| Myopathy & severity of COVID-19 | 0.0001 | significant | 0.00115 |
| Smell Impairment & severity of COVID-19 | 0.0003 | significant | 0.0023 |
| Disturbance of consciousness & severity of COVID-19 | 0.0007 | significant | 0.004025 |
| Fatigue & severity of COVID-19 | 0.009 | significant | 0.038333333 |
| Taste Impairment & severity of COVID-19 | 0.01 | significant | 0.038333333 |
| Acute Confusion/Delirium & severity of COVID-19 | 0.09 | not significant | 0.276 |
| Neuralgia & severity of COVID-19 | 0.11 | not significant | 0.276 |
| Total Stroke & severity of COVID-19 | 0.12 | not significant | 0.276 |
| Neurological manifestations & mortality | 0.12 | not significant | 0.276 |
| Seizure & severity of COVID-19 | 0.2 | not significant | 0.418181818 |
| Dysautonomia & severity of COVID-19 | 0.32 | not significant | 0.558571429 |
| Encephalopathy & severity of COVID-19 | 0.32 | not significant | 0.558571429 |
| Headache & Dizziness & severity of COVID-19 | 0.34 | not significant | 0.558571429 |
| Myalgia & severity of COVID-19 | 0.4 | not significant | 0.613333333 |
| Rhabdomyolysis & severity of COVID-19 | 0.46 | not significant | 0.635882353 |
| Movement disorder & severity of COVID-19 | 0.47 | not significant | 0.635882353 |
| Myalgia or Fatigue & severity of COVID-19 | 0.6 | not significant | 0.766666667 |
| Vision Impairment & severity of COVID-19 | 0.65 | not significant | 0.7705 |
| Headache & severity of COVID-19 | 0.67 | not significant | 0.7705 |
| Encephalitis & severity of COVID-19 | 0.79 | not significant | 0.865238095 |
| Dizziness & severity of COVID-19 | 0.92 | not significant | 0.961818182 |
| Neuropsychiatric disorder & severity of COVID-19 | 0.99 | not significant | 0.99 |

***False Discovery Rate: 5%**

**Table 4:** Subgroup analysis based on risk of bias for the prevalence of neurological symptoms and diagnoses

| **S. No** | **Neurological symptom** | **No. of studies (N); Prevalence of studies with a low risk of bias; I-square** | **No. of studies (N); Prevalence of studies with moderate related to risk of bias; I-square** | **No. of studies (N); Prevalence of studies with high risk of bias; I-square** | **p-value for test for subgroup differences** |
| --- | --- | --- | --- | --- | --- |
| **Neurological symptoms** | | | | |  |
| 1. | Acute confusion/ delirium | N=17; 11% (95%CI 7 to 16%); I^2^= 98.7% | N=2; 42% (95%CI 7 to 83%); I^2^= 95.5% | - | 0.06 |
| 2. | Myalgia or Fatigue | N=15; 24% (95%CI 19 to 29%); I^2^= 83.3% | N=6; 43% (95%CI 33 to 53%); I^2^= 78.8% | N=1; 68% (95%CI 48 to 85%) | **<0.001** |
| 3. | Headache | N=172; 13% (95%CI 11 to 15%); I^2^= 97.1% | N=27; 18% (95%CI 11 to 27%); I^2^=98.4% | N=3; 20% (95%CI 2 to 48%); I^2^=91.9% | 0.32 |
| 4. | Dizziness | N= 38; 7% (95%CI 5 to 8%); I^2^= 91.3% | N=6; 6% (95%CI 2 to 13%); I^2^=91.5% | N=2; 16% (95%CI 9 to 25%); I^2^=28% | **0.02** |
| 5. | Disturbance of Consciousness | N=18; 9% (95%CI 6 to 13%); I^2^= 97.1% | N=6; 4% (95%CI 1 to 8%); I^2^= 91.3% | N=1; 1% (95%CI 0 to 5%) | **0.009** |
| 6. | Smell Impairment | N=41; 18%; (95%CI 12 to 24%); I^2^= 99.1% | N=9; 21% (95%CI 6 to 42%); I^2^= 99.3% | N=1; 33% (95%CI 20 to 48%) | 0.11 |
| 7. | Taste Impairment | N=30; 19% (95%CI 13 to 27%); I^2^= 98.7% | N=7; 29% (95%CI 12 to 50%); I^2^= 99% | N=1; 36% (95%CI 22 to 51%) | 0.10 |
| 8. | Vision Impairment | N=8; 4% (95%CI 1 to 10%); I^2^= 95.4% | N=2; 5% (95%CI 2 to 10%); I^2^=81.6% | - | 0.70 |
| 9. | Seizure | N=12; 1% (95%CI 0.37 to 2%); I^2^= 92% | N=3; 0.41% (95%CI 0 to 2%); I^2^=70.7% | - | 0.64 |
| 10. | Myalgia | N=171; 20% (95%CI 18 to 22%); I^2^= 97.1% | N=31; 22% (95%CI 14 to 30%); I^2^= 98% | N=5; 31% (95%CI 11 to 55%); I^2^= 92.8% | 0.54 |
| 11. | Fatigue | N=148; 32% (95%CI 29 to 35%); I^2^= 97.6% | N=18; 32% (95%CI 25 to 40%); I^2^= 92.9% | N=3; 40% (95%CI 15 to 68%); I^2^= 93% | 0.87 |
| 12. | Smell or Taste Impairment | N=13; 19% (95%CI 10 to 30%); I^2^= 97.6% | N=1; 7% (95%CI 3 to 13%) | - | **0.02** |
| **Neurological diagnosis** | | | | |  |
| 13. | Total Stroke | N=24; 2% (95%CI 1 to 2%); I^2^= 84.1% | N=5; 3% (95%CI 1 to 8%); I^2^= 90% | - | 0.17 |

N= Number of studies

**Table 5:** Subgroup analysis based on risk of bias for the association between severity and neurological symptoms

| **S. No** | **Neurological symptom** | **OR and 95% CI in studies with a low risk of bias; I-square** | **OR and 95% CI in studies with moderate with risk of bias; I-square** | **OR and 95% CI in studies with high risk of bias; I-square** | **p-value for test for subgroup differences** |
| --- | --- | --- | --- | --- | --- |
| 1. | Headache | N=28; (OR 0.88; 95%CI 0.75 to 1.03); I^2^= 0% | N=4; (OR 2.46; 95%CI 0.74 to 8.15); I^2^= 80% | - | 0.1 |
| 2. | Myalgia | N=28; (OR 1.06; 0.88 to 1.29); I^2^= 36% | N=2; (OR 1.83; 95%CI 0.73 to 4.60); I^2^= 12% | N=1; (OR 0.49; 95% CI 0.04-5.93) | 0.43 |
| 3. | Fatigue | N=30; (OR 1.24; 95%CI 1.06 to 1.46); I^2^= 22% | N=2; (OR 4.20; 95%CI 0.48 to 36.65); I^2^= 80% | N=1; (OR 0.23; 95%CI 0.05 to 1.08) | 0.055 |
| 4. | Dizziness | N=10; (OR 1.05; 95%CI 0.68 to 1.62); I^2^= 38% | N=1; (OR 0.56; 95%CI 0.21 to 1.48) | - | 0.24 |

N= Number of studies

**Table 6:** Inter-rater agreement for analyzing the hospitalization status of COVID-19 patients in 35 studies.

|  | **Rater 2** | | **No. of studies= 35** | |
| --- | --- | --- | --- | --- |
| **Rater 1** | 0 (No) | 1 (Yes) | **Kappa* (95%CI)** | **Agreement** |
| **Hospitalized**  0 (No)  1 (Yes)  **Non-Hospitalized**  0 (No)  1 (Yes)  **Combined**  0 (No)  1 (Yes)  **Unclear**  0 (No)  1 (Yes) | 2  2  32  0  34  1  33  1 | 1  30  2  1  -  -  1  0 | 0.525 (0.055 to 0.995)  0.478 (-0.122 to 1.000)  Not Estimable  -0.029 (-0.099 to 0.040) | 91.43%  94.29%  97.14%  94.29% |

**Table 7:** Inter-rater agreement for analyzing the risk of bias in 35 studies.

|  | **Rater 2** | | | **No. of studies= 35** | |
| --- | --- | --- | --- | --- | --- |
| **Rater 1** | 0 (No) | 1 (Unclear) | 2 (Yes) | **Kappa* (95%CI)** | **Agreement** |
| **Item 1**  0 (No)  1 (Unclear)  2 (Yes)  **Item 2**  0 (No)  1 (Unclear)  2 (Yes)  **Item 3**  0 (No)  1 (Unclear)  2 (Yes)  **Item 4**  0 (No)  1 (Unclear)  2 (Yes)  **Item 5**  0 (No)  1 (Unclear)  2 (Yes)  **Item 6**  0 (No)  1 (Unclear)  2 (Yes)  **Item 7**  0 (No)  1 (Unclear)  2 (Yes)  **Item 8**  0 (No)  1 (Unclear)  2 (Yes) | 0  0  0  0  0  0  0  0  0  0  0  0  3  0  2  0  0  1  5  1  0  0  0  1 | 0  0  0  0  0  0  0  1  0  0  1  4  0  6  8  0  0  0  1  9  0  0  0  0 | 3  0  32  3  0  32  0  1  33  0  1  29  0  1  15  1  0  33  3  5  11  0  2  32 | Not Estimable  Not Estimable  0.653 (0.026 to 1.000)  0.222 (-0.222 to 0667)  0.486 (0.111 to 0.597)  Not Estimable  0.566 (0.309 to 0.740)  Not Estimable | 91.43%  91.43%  97.14%  85.71%  68.57%  94.29%  71.43%  91.43% |

**Key:** Item 1- Clear Inclusion criteria in case series; Item 2- Condition measured in a standard, reliable way; Item 3- Valid methods used for identification of the condition; Item 4- Consecutive inclusion of participants; Item 5- Complete inclusion of participants; Item 6- Clear reporting of demographics; Item 7- Clear reporting of clinical information; Item 8- Clear reporting of presenting site(s)/clinic(s) demographic information

**Table 8:** Subgroup analysis based on hospitalization status of the patients

| **S. No** | **Neurological symptom** | **N=Number of studies; Pooled prevalence (95%CI) in hospitalized cases** | **N=Number of studies; Pooled prevalence (95%CI) in non-hospitalized cases** | **p-value for test for subgroup differences** |
| --- | --- | --- | --- | --- |
| 1 | Headache | N=176; 11% (95%CI 10 to 12%) | N=8; 30% (95%CI 23-38%) | **<0.0001** |
| 2 | Dizziness | N=40; 7% (95%CI 6 to 9%) | N=2; 3% (95%CI 0 to 22%) | 0.57 |
| 3 | Smell Impairment | N=35; 11% (95%CI 8 to 15%) | N=4; 50% (95%CI 13 to 86%) | **0.03** |
| 4 | Taste Impairment | N=22; 13% (95%CI 8 to 19%) | N=4; 44% (95%CI 7 to 86%) | 0.13 |
| 5 | Myalgia | N=177; 18% (95%CI 16 to 19%) | N=11; 31% (95%CI 17 to 47%) | 0.06 |
| 6 | Fatigue | N=157; 31% (95%CI 28 to 34%) | N=6; 21% (95%CI 11 to 34%) | 0.12 |
| 7 | Myalgia or Fatigue | N=22; 30% (95%CI 24 to 36%) | N=1; 75% (95%CI 46 to 96%) | **0.002** |
| 8 | Smell or Taste Impairment | N=12; 13% (95%CI 7 to 21%) | N=2; 5% (95%CI 1 to 15%) | 0.14 |

**Table 9:** Risk of bias (quality) assessment using the JBI tool for the studies included in the systematic review and meta-analysis

| **S. No** | **Author** | **Point 1** | **Point 2** | **Point 3** | **Point 4** | **Point 5** | **Point 6** | **Point 7** | **Point 7.1** | **Point 7.2** | **Point 7.3** | **Point 7.4** | **Point 8** | **Overall** |
| --- | --- | --- | --- | --- | --- | --- | --- | --- | --- | --- | --- | --- | --- | --- |
| 1 | Mao L, 2020 | Low | Low | Low | Low | Low | Moderate | Low | Low | Moderate | Low | Low | Low | Low |
| 2 | Helms J, 2020a | Low | Low | Low | Low | High | High | Moderate | High | High | Low | Low | Low | Moderate |
| 3 | Hornuss D, 2020 | Low | Low | Low | Low | Low | Moderate | High | High | High | High | Low | Low | Low |
| 4 | Levinson R, 2020 | Low | High | High | Moderate | Moderate | High | High | High | High | High | High | High | High |
| 5 | Yan CH, 2020 | High | Moderate | Moderate | Moderate | Moderate | Low | Moderate | Low | High | High | High | High | Moderate |
| 6 | Chen N, 2020 | Low | Low | Moderate | Low | Low | Low | Low | Low | Low | Low | Low | Low | Low |
| 7 | Guan W, 2020 | Low | Low | Low | Low | Low | Low | Low | Low | Low | Low | Low | Low | Low |
| 8 | Chen P, 2020 | Low | Moderate | Moderate | Moderate | Moderate | Low | Moderate | High | Moderate | High | Moderate | Moderate | Moderate |
| 9 | Chen Q, 2020 | Low | Moderate | Low | High | High | Moderate | Moderate | Low | Moderate | High | Moderate | Low | Moderate |
| 10 | Chen T, 2020 | Low | Low | Low | Low | Low | Low | Low | Low | Low | Low | Low | Low | Low |
| 11 | Chen X, 2020a | Low | Low | Low | Low | Low | Low | Low | Low | Low | High | Low | Low | Low |
| 12 | Cummings MJ, 2020 | Low | Low | Low | Low | Moderate | Low | Low | Low | Low | High | Low | Low | Low |
| 13 | Du W, 2020 | Low | Low | Low | Low | Moderate | Low | Low | Low | Low | Low | High | Low | Low |
| 14 | Jin X, 2020 | Low | Low | Low | Low | Moderate | Moderate | Low | Low | Low | Low | Low | Low | Low |
| 15 | Kim ES, 2020 | Low | Low | Low | Low | Low | Low | Low | Low | Low | Low | High | Low | Low |
| 16 | Liang WH, 2020 | Low | Low | Low | Low | Moderate | Low | Low | Low | Low | Low | Low | Low | Low |
| 17 | Liu K, 2020 | Low | Low | Low | Low | Moderate | Low | Low | Low | Low | Low | High | Low | Low |
| 18 | Ma Y, 2020 | Low | Low | Low | Low | Low | Moderate | Low | Low | Low | Low | Low | Low | Low |
| 19 | Niu S, 2020 | Low | Low | Low | Low | Low | Moderate | Low | Low | Low | Low | Low | Low | Low |
| 20 | Shen L, 2020 | Low | Low | Low | Low | Moderate | Low | Low | Low | Low | High | Low | Low | Low |
| 21 | Sun Y, 2020 | Low | Low | Low | Moderate | Moderate | Low | Low | Low | Low | Low | High | Low | Low |
| 22 | Tian S, 2020 | Low | Low | Low | Low | Moderate | Low | Low | High | Low | High | Low | Low | Low |
| 23 | Tomlins J, 2020 | Low | Low | Low | Low | Low | Low | Low | Low | Low | Low | High | Low | Low |
| 24 | Wang D, 2020a | Low | Low | Low | Low | Low | Low | Low | Low | Low | Low | Low | Low | Low |
| 25 | Wang D, 2020b | Low | Low | Low | Low | Low | Low | Low | Low | Low | Low | Low | Low | Low |
| 26 | Wang L, 2020a | Low | Low | Low | Low | Moderate | Low | Low | Low | Low | Low | Low | Low | Low |
| 27 | Romero-Sanchez C, 2020 | Low | Low | Low | Low | Low | Low | Low | Low | Low | Low | High | Low | Low |
| 28 | Wang X, 2020a | Low | Low | Low | Low | Moderate | Moderate | Low | Low | High | Low | Low | Low | Low |
| 29 | Wang X, 2020b | Low | Low | Low | Low | Low | Low | Low | Low | Low | Low | Low | Low | Low |
| 30 | Xu YH, 2020 | Low | Low | Low | Low | Moderate | High | High | High | High | Low | High | Low | Moderate |
| 31 | Yang W, 2020 | Low | Low | Low | Low | Moderate | Low | Low | Low | Low | Low | High | Low | Low |
| 32 | Yang X, 2020 | Low | Low | Low | Low | Low | Moderate | Low | Low | Low | Low | Low | Low | Low |
| 33 | Yin S, 2020 | Low | Low | Low | Low | Moderate | Low | Low | High | Low | Low | Low | Low | Low |
| 34 | Huang C, 2020 | Low | Low | Low | Low | Moderate | Low | Low | Low | Low | Low | Low | Low | Low |
| 35 | Zhang G, 2020 | Low | Low | Low | Low | Moderate | Low | Low | Low | Low | Low | Low | Low | Low |
| 36 | Zhao XY, 2020 | High | Moderate | Low | Moderate | High | Low | Low | Low | High | Low | Low | Low | Moderate |
| 37 | Zheng Y, 2020a | High | Moderate | Low | Low | Low | Low | Low | Low | High | Low | Low | Low | Low |
| 38 | Wan S, 2020 | High | High | Low | Moderate | Low | Low | Low | Low | Low | Low | Low | Low | Moderate |
| 39 | Qian GQ, 2020 | High | Moderate | Low | Moderate | High | Low | Low | Low | High | Low | Low | Low | Moderate |
| 40 | Hu Z, 2020 | High | Moderate | Low | Moderate | Moderate | Low | Low | Low | High | Low | Low | Low | Moderate |
| 41 | Wu J, 2020a | High | High | Low | Low | Moderate | Low | Low | Low | High | Low | Low | Low | Moderate |
| 42 | Xu XW, 2020 | High | High | Low | Moderate | High | Low | Low | Low | High | Low | Low | Low | Moderate |
| 43 | Zhang X, 2020 | High | High | Moderate | Moderate | Moderate | Low | Moderate | Low | High | High | Low | Low | Moderate |
| 44 | Cai Q, 2020 | High | Low | Low | Low | Low | Low | Low | Low | Low | Low | Low | Low | Low |
| 45 | Kong I, 2020 | High | High | High | Moderate | High | Low | High | High | High | High | Low | High | High |
| 46 | Gupta N, 2020 | Low | Moderate | Low | Moderate | Moderate | Low | Low | Low | Low | High | Low | Low | Low |
| 47 | Karadas O, 2020 | High | High | Low | Low | Low | Low | Low | Low | Low | Low | Low | Low | Moderate |
| 48 | Beltran-Corbellini A, 2020 | Low | Moderate | Low | Low | Low | Low | Moderate | Low | Moderate | High | Low | Low | Low |
| 49 | Lechien JR, 2020a | Low | Moderate | Low | Moderate | Moderate | High | High | High | High | High | Low | Low | Moderate |
| 50 | Han R, 2020 | Low | Low | Low | Moderate | Moderate | Low | Moderate | High | Low | Low | High | Low | Low |
| 51 | Lo IL, 2020 | Low | Low | Low | Moderate | Moderate | Low | Low | Low | Low | Low | Low | Low | Low |
| 52 | Du Y, 2020 | Low | Low | Low | Low | Low | Low | Low | Low | High | Low | Low | Low | Low |
| 53 | Han X, 2020 | Low | Low | Low | Moderate | Moderate | Low | Moderate | Low | High | Low | High | Low | Low |
| 54 | Lian J, 2020a | Low | Low | Low | Low | Moderate | Moderate | Low | Low | Low | Low | Low | Low | Low |
| 55 | Shi S, 2020 | Low | Low | Low | Low | Low | Low | Moderate | Low | High | Low | High | High | Low |
| 56 | Lei P, 2020 | Low | Low | Low | Low | Moderate | Low | Low | Low | High | Low | Low | Low | Low |
| 57 | Zheng F, 2020 | Low | Low | Low | Low | Moderate | Low | Low | Low | Low | Low | High | Low | Low |
| 58 | Wang Z, 2020a | Low | Low | Low | Low | Low | Low | High | Low | High | Moderate | Moderate | Low | Low |
| 59 | Bhatraju PK, 2020 | Low | Low | Low | Low | Moderate | Moderate | Low | Low | High | Low | Low | Low | Low |
| 60 | Chang D, 2020 | Low | Low | Low | Low | Moderate | Low | High | High | High | Moderate | High | Low | Low |
| 61 | Xu X, 2020a | Low | Low | Low | Low | Low | Low | Moderate | Low | High | Low | High | Low | Low |
| 62 | Liu Y, 2020 | Low | Low | Low | Low | Moderate | Moderate | Low | Low | Low | Low | Low | Low | Low |
| 63 | Zhou F, 2020 | Low | Low | Low | Low | Low | Low | Low | Low | Low | Low | Low | Low | Low |
| 64 | Han YN, 2020 | Low | Low | Low | Moderate | Moderate | Low | Moderate | Low | High | Low | High | Low | Low |
| 65 | Cao J, 2020 | Low | Low | Low | Low | Low | Low | Low | Low | High | Low | Low | Low | Low |
| 66 | Zhao W, 2020 | Low | Low | Low | Low | Moderate | Low | Moderate | Low | High | Low | High | Low | Low |
| 67 | Li X, 2020a | Low | Low | Low | Moderate | Moderate | Low | High | High | High | Low | High | Low | Moderate |
| 68 | Feng Y, 2020 | Low | Low | Low | Low | Moderate | Low | Low | Low | Low | Low | Low | Low | Low |
| 69 | Lei Z, 2020 | Low | Low | Low | Low | Low | Low | Low | Low | Low | Low | High | Low | Low |
| 70 | Zhang R, 2020 | High | Moderate | Low | Low | Low | Low | Moderate | Low | High | Low | Moderate | Low | Moderate |
| 71 | Barrasa H, 2020 | Low | Low | Low | Low | Low | Low | Low | Low | Low | High | Low | Low | Low |
| 72 | Li YK, 2020 | High | High | High | Moderate | High | Moderate | Low | Low | Low | Low | Low | Low | High |
| 73 | Chen TL, 2020 | Low | Moderate | Low | High | Moderate | Low | Low | Low | Low | Low | Low | Low | Moderate |
| 74 | Chu J, 2020 | High | High | High | High | Moderate | High | Moderate | High | Low | Low | Moderate | Low | High |
| 75 | Cheng Z, 2020 | Low | Moderate | Low | High | Moderate | High | Moderate | High | High | Low | Low | Low | Moderate |
| 76 | Goyal P, 2020 | Low | Low | Low | Low | Low | Low | Low | Low | Moderate | Low | Low | Low | Low |
| 77 | Chen J, 2020a | Low | Moderate | Low | High | Moderate | Low | Low | Low | High | Low | Low | Low | Moderate |
| 78 | Escalera-Antezana JP, 2020 | Low | Moderate | Low | High | Moderate | Moderate | High | High | High | High | Low | Low | Moderate |
| 79 | Merza MA, 2020 | Low | Low | Low | Moderate | Moderate | Moderate | Low | Low | Low | Low | Low | Low | Low |
| 80 | Wang R, 2020 | Low | Low | Low | Moderate | Moderate | Low | Moderate | Low | High | Moderate | Low | High | Moderate |
| 81 | Zheng Y, 2020b | Low | Low | Low | High | Moderate | Low | Moderate | Low | Low | Moderate | Moderate | Low | Moderate |
| 82 | Garg R, 2020 | Low | Low | Low | Low | Low | Moderate | Moderate | Low | High | High | Low | Low | Low |
| 83 | Li Y, 2020 | Low | Low | Low | Low | Low | Low | Low | Low | Low | Low | Low | Low | Low |
| 84 | Chung M, 2020 | Low | Moderate | Moderate | Low | Moderate | Moderate | Moderate | High | High | Low | Low | Low | Moderate |
| 85 | Du RH, 2020 | High | High | High | Low | Low | Low | High | High | High | High | Moderate | Low | Moderate |
| 86 | Li X, 2020b | Moderate | Moderate | Moderate | Low | Low | Low | Low | Low | Low | Low | Low | Low | Low |
| 87 | Pan F, 2020 | Low | Low | Low | Moderate | Moderate | Moderate | Moderate | High | Moderate | Low | Low | Low | Moderate |
| 88 | Shi H, 2020 | Low | Low | Low | Low | High | Low | Low | Low | High | Low | Low | Low | Low |
| 89 | Song F, 2020 | Low | Low | Low | Moderate | High | Moderate | Low | Low | High | Low | Low | Low | Moderate |
| 90 | Wu J, 2020b | Low | Low | Low | High | High | Moderate | Moderate | Low | High | Low | Moderate | High | Moderate |
| 91 | Zhang JJ, 2020 | Low | Low | Low | Low | Low | Low | Low | Low | Low | Low | Low | Low | Low |
| 92 | Duanmu Y, 2020 | Low | Low | Low | Low | Moderate | Low | Low | Low | High | Low | Low | Low | Low |
| 93 | Giacomelli A, 2020 | Low | Low | Moderate | Moderate | Moderate | Low | High | High | High | High | High | Low | Moderate |
| 94 | Li K, 2020 | Low | Low | Low | Low | Moderate | Low | Low | Low | Low | Low | High | Low | Low |
| 95 | Jain R, 2020 | Low | Low | Low | Low | Low | Moderate | High | High | High | Low | High | Low | Low |
| 96 | Qin C, 2020 | Low | Low | Low | Low | Low | Low | Low | Low | Low | Low | Low | Low | Low |
| 97 | Merkler AE, 2020 | Low | Low | Low | Low | Moderate | Low | Low | Low | High | Low | Low | Low | Low |
| 98 | Radmanesh A, 2020 | Low | Low | Low | Moderate | Moderate | High | High | High | High | Low | High | Low | Moderate |
| 99 | Xiong W, 2020 | Low | Low | Low | Low | Moderate | Low | Low | Low | Low | Low | Low | Low | Low |
| 100 | Mahammedi A, 2020 | Low | Low | Low | Low | Moderate | Low | Low | Low | High | Low | Low | Low | Low |
| 101 | Agarwal P, 2020 | Low | Low | Low | Low | Moderate | Low | High | Low | High | Moderate | High | Low | Low |
| 102 | Notz Q, 2020 | Low | Moderate | Moderate | Low | Moderate | Moderate | Low | High | Low | Low | Low | Low | Moderate |
| 103 | Helms J, 2020b | Low | Low | Low | Low | High | Low | Low | Low | Low | Low | Low | Low | Low |
| 104 | Ora J, 2020 | Low | Low | Moderate | Low | Moderate | Low | High | High | Low | High | High | Low | Moderate |
| 105 | Duployez N, 2020 | Low | Low | Low | Low | Low | Low | Moderate | Low | Low | High | High | Low | Low |
| 106 | Fu J, 2020 | Low | Low | Low | Low | Moderate | Low | Low | Low | Low | Low | Low | Low | Low |
| 107 | Gao Y, 2020 | Low | Low | Low | Low | High | Moderate | Low | Low | Low | Low | Low | Low | Low |
| 108 | Gu Q, 2020 | Low | Low | Low | Low | High | Low | High | High | High | Low | High | Low | Moderate |
| 109 | Guo T, 2020 | Low | Low | Low | Moderate | Moderate | Moderate | High | High | High | Low | High | Low | Moderate |
| 110 | Hafiz M, 2020 | Low | Low | Low | Low | Low | Low | Moderate | Low | High | Low | High | Low | Low |
| 111 | He F, 2020 | Low | Low | Low | Low | Moderate | Moderate | Low | Low | Low | Low | High | Low | Low |
| 112 | Hu HT, 2020 | Low | Low | Low | Low | Low | Low | Low | Low | Low | Low | Low | Low | Low |
| 113 | Huang R, 2020 | Low | Low | Low | Low | Moderate | Moderate | Low | Low | High | Low | Low | Low | Low |
| 114 | Jiang H, 2020 | Low | Low | Low | Low | High | Low | Moderate | Low | High | Low | High | Low | Low |
| 115 | Kato H, 2020 | Moderate | Low | Low | Moderate | High | Moderate | Low | Low | Moderate | Low | Low | Moderate | Moderate |
| 116 | Khan M, 2020 | Low | Low | Low | Low | Low | Low | High | Low | High | High | High | Low | Low |
| 117 | Klopfenstein T, 2020 | Low | Low | Low | Low | Low | Low | High | Low | High | High | High | Low | Low |
| 118 | Lapostolle F, 2020 | Moderate | Low | Moderate | Low | Low | Low | High | High | High | High | High | Low | Moderate |
| 119 | Liu D, 2020a | Low | Low | Moderate | Low | High | Low | Low | Low | Low | Low | Moderate | Low | Low |
| 120 | Liu J, 2020a | Moderate | Low | Low | Low | Low | Low | Low | Low | Low | Low | Low | Low | Low |
| 121 | Mishra P, 2020 | Low | Moderate | High | Moderate | Low | Low | High | High | High | High | Moderate | Moderate | Moderate |
| 122 | Qu J, 2020 | Moderate | Low | Low | Low | Low | Low | Low | Low | Low | Low | Moderate | Low | Low |
| 123 | Ren D, 2020 | Moderate | Moderate | High | Low | Low | Low | Moderate | Low | High | High | Moderate | Low | Moderate |
| 124 | Trigo J, 2020 | Low | Low | Low | Low | Low | Low | Moderate | Low | High | High | Low | Low | Low |
| 125 | Wang Y, 2020 | Low | Low | Low | Low | Low | Low | Moderate | High | High | Low | Moderate | Low | Low |
| 126 | Wang ZH, 2020 | Low | Low | Low | Low | Low | Low | Low | Low | Low | Moderate | Low | Low | Low |
| 127 | Wei Y, 2020a | Low | Low | Low | Low | Low | Low | Low | Low | Low | Low | Moderate | Low | Low |
| 128 | Wu G, 2020 | Low | Low | Low | Low | Low | Low | Low | Low | Low | Low | Low | Low | Low |
| 129 | Xie Y, 2020 | Low | Low | Low | Low | Low | Low | Low | Low | Low | Low | High | Low | Low |
| 130 | Xiong Y, 2020 | Low | Low | Low | Low | Low | Moderate | Low | Low | Low | Low | High | Low | Low |
| 131 | Xu X, 2020b | Moderate | Low | Low | Low | Low | Low | Low | Low | Low | High | Low | Low | Low |
| 132 | Yang L, 2020 | Moderate | Low | Low | Low | Low | Low | Moderate | Low | High | Moderate | Low | Low | Low |
| 133 | Yang Q, 2020 | Low | Low | Low | Moderate | Low | Low | Low | Low | Low | High | Low | Low | Low |
| 134 | Zhao D, 2020 | Low | Low | Low | Low | Low | Low | Low | Low | High | Low | Low | Low | Low |
| 135 | Zhao G, 2020 | Low | Low | Low | Low | Low | Low | High | High | Low | High | High | Low | Low |
| 136 | Zheng Y, 2020c | Low | Low | Low | Low | Low | Low | Low | Low | High | Low | Low | Low | Low |
| 137 | Guilmot A, 2020 | Low | Low | Low | Low | High | Moderate | Low | Low | Low | High | Low | Moderate | Moderate |
| 138 | Liguori C, 2020 | Low | Low | Moderate | Low | Low | Low | High | High | High | High | High | Low | Low |
| 139 | Romagnolo A, 2020 | Low | Low | Low | Low | Low | Low | Moderate | Low | High | High | Low | Low | Low |
| 140 | Asai N, 2020 | Moderate | Low | Low | Moderate | Low | Low | Moderate | Low | High | Low | High | Low | Low |
| 141 | Cavalleri M, 2020 | Low | Low | Low | Low | Low | Low | High | High | Low | High | High | Low | Low |
| 142 | Davarpanah AH, 2020 | Low | Low | Moderate | Low | Low | Low | Moderate | Low | High | Low | Moderate | Low | Low |
| 143 | Faury H, 2020 | Low | Moderate | High | Low | High | Low | Moderate | Low | High | Low | Moderate | Low | Moderate |
| 144 | Feng G, 2020 | Low | Low | Low | Low | Low | Low | Low | Low | Low | Low | Low | Low | Low |
| 145 | Kang MK, 2020 | Low | Low | Low | Low | Low | Low | Low | Low | High | Low | Low | Low | Low |
| 146 | Liu D, 2020b | Low | Low | Low | Moderate | Low | Low | Low | Low | High | Low | Low | Moderate | Low |
| 147 | Marvisi M, 2020 | Low | Low | Low | Low | Low | Low | Moderate | Low | High | Low | Moderate | Low | Low |
| 148 | Meiler S, 2020 | Low | Low | Low | Low | Low | Low | Low | Low | High | Low | Low | Low | Low |
| 149 | Salepci E, 2020 | Low | Low | Low | Low | Low | Low | Low | Low | Low | High | Low | Low | Low |
| 150 | Shah SJ, 2020 | Low | Low | Low | Low | Low | Low | Low | Low | High | Low | Low | Low | Low |
| 151 | Wang W, 2020a | Low | Low | Low | Low | Low | Low | Low | Low | Low | High | Low | Low | Low |
| 152 | Xiang C, 2020 | Low | Low | Low | Low | Low | Low | High | High | High | Low | High | Low | Low |
| 153 | Zandkarimi E, 2020 | Low | Low | Low | Low | Low | Low | High | Low | High | High | Moderate | Low | Low |
| 154 | Zhang J, 2020 | Moderate | Low | Moderate | Low | Low | Low | Moderate | Low | High | Low | Moderate | Low | Low |
| 155 | Zhou J, 2020 | Low | Low | Low | Low | Low | Low | Moderate | High | High | Low | Low | Low | Low |
| 156 | Abbas HM, 2020 | Low | Low | Low | Low | Low | Low | Low | Low | Low | High | Low | Low | Low |
| 157 | Campioli CC, 2020 | Low | Low | Low | Low | Low | Low | High | Low | High | High | Moderate | Low | Low |
| 158 | Chen X, 2020b | Low | Low | Low | Low | Low | Low | High | High | High | Low | High | Low | Low |
| 159 | Chon YJ, 2020 | Low | Low | Low | Low | Low | Low | Low | Low | Low | Low | Moderate | Low | Low |
| 160 | Coppola A, 2020 | Moderate | Low | Moderate | Moderate | Low | High | High | High | High | High | High | High | Moderate |
| 161 | Gatti M, 2020 | Low | Low | Low | Low | Low | Low | Low | Low | High | Low | Low | Low | Low |
| 162 | Gu J, 2020 | Low | Low | Low | Low | Low | Low | Moderate | High | High | Low | Low | Low | Low |
| 163 | Kuzan TY, 2020 | Low | Low | Moderate | Low | Low | Low | Moderate | High | High | Low | Low | Low | Low |
| 164 | Li J, 2020a | Low | Low | Low | Low | Low | Low | Low | Low | Low | Low | Low | Low | Low |
| 165 | Li J, 2020b | Low | Low | Moderate | Low | Low | Low | Moderate | Low | Low | High | Moderate | Low | Low |
| 166 | Lian J, 2020b | Low | Low | Low | Low | Low | Low | Low | Low | Low | Low | Low | Low | Low |
| 167 | Liu Y, 2020b | Low | Low | Low | Low | Low | Low | Moderate | Low | Low | High | Moderate | Low | Low |
| 168 | Moon SS, 2020 | Low | Moderate | Moderate | Low | Low | Low | Moderate | Low | High | Low | Moderate | Low | Low |
| 169 | Tabata S, 2020 | Low | Low | Low | Low | Low | Low | Low | Low | Low | Low | Low | Low | Low |
| 170 | Tan JY, 2020 | Low | Low | Low | Low | High | Moderate | Low | Low | High | Low | Low | Low | Low |
| 171 | Wang H, 2020 | Low | Low | Low | Low | Low | Low | Moderate | High | High | Low | Low | Low | Low |
| 172 | Wang W, 2020b | Low | Low | Low | Low | Low | Low | Low | Low | High | Low | Low | Low | Low |
| 173 | Wang X, 2020c | Low | Low | Moderate | Low | Low | Low | Moderate | Low | Low | High | Moderate | Low | Low |
| 174 | Xu H, 2020 | Low | Moderate | Moderate | Low | Low | Low | Moderate | Low | High | High | Low | Low | Low |
| 175 | Ye H, 2020 | Low | Low | Low | Low | Low | Low | Moderate | Low | High | High | Low | Low | Low |
| 176 | Zha L, 2020 | Moderate | Low | Low | Low | Low | Low | Moderate | Low | Low | High | Moderate | Low | Low |
| 177 | Zhang L, 2020a | Low | Low | Low | Low | Low | Low | Moderate | Low | High | High | Low | Low | Low |
| 178 | Zhou H, 2020 | Low | Low | Low | Low | Low | Low | Low | Low | Low | Low | Low | Low | Low |
| 179 | Zhuang Y, 2020 | Low | Low | Moderate | Low | Low | Low | High | High | High | Low | High | Low | Low |
| 180 | Lechien JR, 2020b | Low | Low | Low | Low | Low | Low | Moderate | Low | Low | High | Moderate | Low | Low |
| 181 | Rothstein A, 2020 | Moderate | Low | Low | Low | Low | Low | Moderate | Low | High | High | Low | Low | Low |
| 182 | Chary E, 2020 | Low | Low | Low | Low | Low | Low | High | High | High | High | Moderate | Low | Low |
| 183 | Scullen T, 2020 | Low | Low | Low | Low | Low | Low | Moderate | Low | High | Low | Moderate | Low | Low |
| 184 | Aggarwal A, 2020 | Low | Low | Low | Low | Low | Low | Low | Low | Low | Low | Low | Low | Low |
| 185 | Alsofayan YM, 2020 | Low | Low | Low | Low | Low | Low | High | Low | High | High | Moderate | Low | Low |
| 186 | Carignan A, 2020 | Low | Low | Low | Low | Low | Low | High | Moderate | High | High | Low | Low | Low |
| 187 | Chen FF, 2020 | Low | Low | Moderate | Low | Low | Low | High | Low | High | High | High | Low | Low |
| 188 | Chen J, 2020c | Low | Low | Moderate | Low | Low | Low | Low | Low | Low | Low | Low | Low | Low |
| 189 | Chen Y, 2020a | Low | Low | Moderate | Low | Low | Low | Moderate | Low | Low | High | Moderate | Low | Low |
| 190 | Duan X, 2020 | Low | Low | Low | Low | Low | Low | Low | Low | Low | Low | Low | Low | Low |
| 191 | Easom N, 2020 | Low | Low | Low | Low | Low | Low | High | High | High | High | Low | Low | Low |
| 192 | Fu Y, 2020 | Low | Low | Low | Low | Low | Low | Moderate | Low | Low | Moderate | Moderate | Low | Low |
| 193 | Gao X, 2020 | Low | Low | Low | Low | Low | Low | Moderate | Low | High | Low | Moderate | Low | Low |
| 194 | Gaur A, 2020 | Low | Low | Low | Low | Low | Moderate | Moderate | Low | High | Low | Moderate | Low | Low |
| 195 | Gupta S, 2020 | Low | Low | Moderate | Low | Low | Low | Moderate | Low | High | High | Low | Low | Low |
| 196 | Jalessi M, 2020 | Low | Low | Low | Low | High | Low | Moderate | Low | High | Low | Moderate | Low | Low |
| 197 | Li G, 2020 | Low | Low | Moderate | Low | Low | Low | High | High | High | High | Low | Low | Low |
| 198 | Li W, 2020 | Low | Low | Low | Low | Low | Low | Low | Low | Low | Low | Low | Low | Low |
| 199 | Martin-Sanchez FJ, 2020 | Low | Low | Low | Low | Low | Low | Moderate | Low | High | Moderate | Low | Low | Low |
| 200 | Meng H, 2020 | Low | Low | Low | Low | Low | Low | Low | Low | Low | Low | Moderate | Low | Low |
| 201 | O'Reilly GM, 2020a | Low | Low | Low | Low | Low | Low | Moderate | Low | High | Moderate | Low | Low | Low |
| 202 | Samrah SM, 2020 | Low | Low | Low | Low | Low | Low | High | Low | High | High | Moderate | Low | Low |
| 203 | Khamis F, 2020 | Low | Low | Low | Low | Low | Low | Moderate | Low | High | Moderate | Low | Low | Low |
| 204 | Saleemi S, 2020 | Low | Low | Low | Low | Low | Low | Moderate | Low | Low | Moderate | High | Low | Low |
| 205 | Shang Y, 2020 | Low | Low | Low | Low | Low | Low | High | High | High | Low | High | Low | Low |
| 206 | Sun D, 2020 | Low | Low | Moderate | Low | Low | Low | Low | Low | Low | Low | Moderate | Low | Low |
| 207 | Wang D, 2020c | Low | Moderate | Moderate | Low | Low | Low | Low | Low | Low | High | Low | Low | Low |
| 208 | Wang F, 2020 | Low | Low | Low | Low | Low | Low | Low | Low | Low | Moderate | Low | Low | Low |
| 209 | Wang S, 2020 | Low | Low | Low | Low | Low | Low | High | Low | High | High | Moderate | Low | Low |
| 210 | Yang J, 2020 | Low | Low | Low | Low | Low | Moderate | High | High | High | Low | High | Moderate | Moderate |
| 211 | Yue H, 2020 | Low | Low | Low | Low | Low | Low | Moderate | Low | Low | Moderate | High | Low | Low |
| 212 | Zhang SY, 2020 | Low | Low | Low | Low | Low | Low | Low | Low | Low | High | Low | Low | Low |
| 213 | Zhang Y, 2020 | Low | Low | Low | Low | Low | Low | Low | Low | Low | Moderate | Low | Low | Low |
| 214 | Zhao K, 2020 | Low | Low | Low | Low | Low | Low | Moderate | Low | High | High | Low | Low | Low |
| 215 | Fan S, 2020 | Low | Low | Low | Low | Low | Low | Moderate | Low | Low | High | Moderate | Low | Low |
| 216 | Hernández-Fernández F, 2020 | Low | Low | Low | Low | Low | Low | Moderate | Low | High | High | Low | Low | Low |
| 217 | García-Azorín D, 2020 | Low | Low | Low | Low | Low | Low | Moderate | Low | High | High | Low | Low | Low |
| 218 | Liu M, 2020a | Moderate | Low | Low | Low | Low | Moderate | Moderate | High | Low | Low | Moderate | Moderate | Moderate |
| 219 | Qin W, 2020 | Low | Moderate | Moderate | Low | Moderate | Low | High | Low | High | High | Moderate | Low | Moderate |
| 220 | Sun C, 2020 | Low | Low | Low | Low | Low | Low | Low | Low | High | Low | Low | Low | Low |
| 221 | Teich VD, 2020 | Moderate | Low | Low | Low | Low | Low | Low | Low | Low | Low | Moderate | Low | Low |
| 222 | Xie W, 2020 | Moderate | Moderate | Moderate | Low | Moderate | Low | Low | Low | Low | Low | Moderate | Low | Moderate |
| 223 | Xu M, 2020 | Low | Low | Low | Low | Low | Low | Low | Low | Low | High | Low | Low | Low |
| 224 | Zeng YL, 2020 | Low | Moderate | Moderate | Low | Low | Low | High | High | High | Low | Moderate | Low | Moderate |
| 225 | Zhang MQ, 2020 | Moderate | Low | Low | Low | Low | Moderate | Moderate | High | High | Low | Low | Low | Low |
| 226 | Chen L, 2020 | Low | Low | Low | Low | Low | Low | Moderate | Low | High | Low | Moderate | Low | Low |
| 227 | Liu C, 2020 | Low | Low | Moderate | Low | Low | Low | Moderate | High | High | Low | Low | Low | Low |
| 228 | Lu XF, 2020 | Low | Low | Low | Low | Low | Low | High | High | High | Low | Moderate | Low | Low |
| 229 | Zhu Q, 2020 | Low | Low | Moderate | Low | Low | Low | Low | Low | Low | High | Low | Low | Low |
| 230 | Zou W, 2020 | Moderate | Low | Low | Low | Low | Low | Low | Low | Low | Low | Moderate | Low | Low |
| 231 | Casas-Rojo JM, 2020 | Low | Low | Low | Low | High | Low | Moderate | Low | Low | High | Moderate | Low | Low |
| 232 | Al-Mutair A, 2020 | Low | Low | Low | Moderate | Moderate | Low | Low | Low | Low | Low | High | Low | Low |
| 233 | Almaki ZS, 2020 | Low | Low | Low | Low | Low | Low | High | Low | High | High | High | Low | Low |
| 234 | An P, 2020 | Low | Low | Low | Low | Low | Low | Low | Low | Low | Low | High | Low | Low |
| 235 | Ayaz A, 2020 | Low | Low | Low | Low | Low | Low | Low | Low | Low | Low | Low | Low | Low |
| 236 | Baghaei P, 2020 | Low | Low | Low | Low | Low | Low | Low | Low | Low | Low | High | Low | Low |
| 237 | Bouzid D, 2020 | Low | Low | Low | Low | Low | Low | Moderate | Low | High | High | Low | Low | Low |
| 238 | Buckner FS, 2020 | Low | Low | Low | Low | Low | Low | Low | Low | Low | Low | High | Low | Low |
| 239 | Caronna E, 2020 | Low | Low | Low | Low | Low | Low | Low | Low | High | Low | Low | Low | Low |
| 240 | Chen HJ, 2020 | Low | Low | Low | Low | High | Low | Low | Low | High | Low | Low | Low | Low |
| 241 | Chen Y, 2020b | Low | Low | Low | Low | Low | Low | Low | Low | High | Low | Low | Low | Low |
| 242 | Chi Q, 2020 | Low | Low | Low | Low | Low | Low | Moderate | Low | High | Low | High | Low | Low |
| 243 | Cobb NL, 2020 | Low | Low | Low | Low | Moderate | Low | Low | Low | Low | Low | Low | Low | Low |
| 244 | Deng LS, 2020 | Low | Low | Low | Moderate | Moderate | Low | Low | Low | Moderate | Low | Low | Low | Low |
| 245 | Du H, 2020 | Low | Low | Low | Low | Moderate | Low | Moderate | Low | High | Low | High | Low | Low |
| 246 | Emami A, 2020 | Low | Low | Low | Low | Low | Low | Low | Low | High | Low | Low | Low | Low |
| 247 | Fu F, 2020 | Low | Low | Low | Low | Low | Low | High | High | High | Low | High | Low | Low |
| 248 | Gao J, 2020 | Low | Low | Low | Moderate | Moderate | Low | Low | Low | Low | Low | High | Low | Low |
| 249 | He S, 2020 | Low | Low | Low | Low | Low | Low | High | Low | High | High | High | Low | Low |
| 250 | Higuchi T, 2020 | Low | Low | Low | Low | Low | Low | Low | Low | Low | High | Low | Low | Low |
| 251 | Homayounieh F, 2020 | Low | Low | Low | Low | Moderate | Low | Low | Low | High | Low | Low | Low | Low |
| 252 | Hong JM, 2020 | Low | Low | Low | Low | Low | Low | Low | Low | Low | Low | Low | Low | Low |
| 253 | Hong L, 2020 | Low | Low | Low | Low | Low | Low | Low | Low | Low | Low | Low | Low | Low |
| 254 | Ibrahim OR, 2020 | Low | Low | Low | Low | Low | Low | Low | Low | Low | Low | High | Low | Low |
| 255 | Jiang J, 2020 | Low | Low | Low | Low | Moderate | Low | Moderate | Low | High | Low | High | Low | Low |
| 256 | Jin XH, 2020 | Low | Low | Low | yea | Low | Low | Moderate | Low | Low | High | High | Low | Low |
| 257 | Jourdes A, 2020 | Low | Low | Low | Low | Low | Low | Low | Low | Low | Low | Low | Low | Low |
| 258 | Kadiane-Oussou NJ, 2020 | Low | Low | Low | Low | Low | Low | Low | Low | Low | High | Low | Low | Low |
| 259 | Kaeuffer C, 2020 | Low | Low | Low | Low | Moderate | Low | Low | Low | Low | High | Low | Low | Low |
| 260 | Khraise NW, 2020 | Low | Low | Low | Low | Low | Low | Moderate | Low | Low | High | High | Low | Low |
| 261 | Kim KH, 2020 | Low | Low | Low | Low | High | Low | Low | Low | Low | High | Low | Low | Low |
| 262 | Kumar A, 2020 | Low | Low | Low | Low | Moderate | Low | Moderate | Low | Low | High | High | Low | Low |
| 263 | Li X, 2020c | Low | Low | Low | Low | Moderate | Low | Low | Low | Low | Low | High | Low | Low |
| 264 | Lima MA, 2020 | Low | Low | Low | Low | Moderate | Low | High | High | Low | High | High | Low | Low |
| 265 | Lin L, 2020 | Low | Low | Low | Low | Moderate | Low | Low | Low | Low | Low | High | Low | Low |
| 266 | Liu H, 2020 | Low | Low | Low | Moderate | Moderate | Moderate | Low | Low | Low | Low | High | Low | Low |
| 267 | Liu J, 2020b | Low | Low | Low | Low | Moderate | Low | Low | Low | Low | Low | Low | Low | Low |
| 268 | Liu M, 2020 | Low | Low | Low | Low | Moderate | Low | Moderate | Low | High | Low | High | Low | Low |
| 269 | Liu W, 2020 | Low | Low | Low | Low | Low | Moderate | Low | Low | Low | Low | High | Low | Low |
| 270 | de Magalhaes JJF, 2020 | Low | Low | Low | Low | Low | Low | Low | Low | Low | High | Low | Low | Low |
| 271 | Mirfazeli FS, 2020 | Low | Low | Low | Low | Low | Low | Low | Low | Low | Low | High | Low | Low |
| 272 | Morikawa M, 2020 | Low | Low | Low | Low | Moderate | Low | Low | Low | Low | High | Low | Low | Low |
| 273 | Novelli L, 2020 | Low | Low | Low | Low | Moderate | Low | Moderate | Low | Low | High | High | Low | Low |
| 274 | Pugliese L, 2020 | Low | Low | Low | Moderate | Moderate | Low | Low | Low | Low | Low | High | Low | Low |
| 275 | Qui Z, 2020 | Low | Low | Low | Low | Moderate | Low | Moderate | High | Low | Low | High | Low | Low |
| 276 | Raberahona M, 2020 | Low | Low | Low | Moderate | Moderate | Low | Moderate | Low | High | High | Low | Low | Low |
| 277 | Ren C, 2020 | Low | Low | Low | Low | Low | Low | Low | Low | Low | High | Low | Low | Low |
| 278 | Shi JC, 2020 | Low | Low | Low | Low | Moderate | Low | Low | High | Low | Low | Low | Low | Low |
| 279 | Sun JT, 2020 | Low | Low | Low | Low | Moderate | Low | Low | Low | Low | High | Low | Low | Low |
| 280 | Sun L, 2020 | Low | Low | Low | Low | Low | Low | Low | Low | Low | Low | Low | Low | Low |
| 281 | Tian J, 2020 | Low | Low | Low | Low | Moderate | Low | Low | Low | Low | Low | High | Low | Low |
| 282 | Vial MR, 2020 | Low | Low | Low | Low | Low | Low | Moderate | Low | Low | High | High | Low | Low |
| 283 | Vilke GM, 2020 | Low | Low | Low | Low | Low | Low | High | High | High | High | High | Low | Low |
| 284 | Wang C, 2020 | Low | Low | Low | Low | Moderate | Low | Low | Low | Low | Low | High | Low | Low |
| 285 | Wang J, 2020a | Low | Low | Low | Low | Moderate | Low | Low | Low | High | Low | Low | Low | Low |
| 286 | Wang J, 2020b | Low | Low | Low | Low | Moderate | Low | Low | Low | Low | Low | Low | Low | Low |
| 287 | Wang Z, 2020b | Low | Low | Low | Low | Moderate | Low | Moderate | Low | High | Low | High | Low | Low |
| 288 | Wei Y, 2020b | Low | Low | Low | Low | Low | Low | Moderate | Low | High | Low | High | Low | Low |
| 289 | Wu B, 2020 | Low | Low | Low | Low | Low | Low | Low | Low | Low | Low | Low | Low | Low |
| 290 | Xiao J, 2020 | Low | Low | Low | Low | High | Low | Low | Low | Low | Low | Low | Low | Low |
| 291 | Xie L, 2020 | Low | Low | Low | Low | Moderate | Low | Low | Low | Low | Low | Low | Low | Low |
| 292 | Zayet S, 2020 | Low | Low | Low | Low | Low | Low | Moderate | Low | High | High | Low | Low | Low |
| 293 | Zhang L, 2020b | Low | Low | Low | Low | Moderate | Low | Low | Low | Low | Low | High | Low | Low |
| 294 | Zhang N, 2020 | Low | Low | Low | Low | Moderate | Low | Low | Low | Low | Low | High | Low | Low |
| 295 | Bach I, 2020 | Low | Low | Low | Low | Low | Low | Moderate | Low | High | Low | High | Low | Low |
| 296 | Battaglini D, 2020 | Low | Low | Low | Moderate | Moderate | Low | Moderate | Low | High | High | Low | Low | Low |
| 297 | Cantador E, 2020 | Low | Low | Low | Low | Moderate | Low | Low | Low | High | Low | Low | Low | Low |
| 298 | Chachkhiani D, 2020 | Low | Low | Low | Low | Moderate | Low | High | Low | High | High | High | Low | Low |
| 299 | Foresti C, 2020 | Low | Low | Moderate | Moderate | Moderate | Low | High | High | Moderate | High | Low | Low | Moderate |
| 300 | Frontera JA, 2020 | Low | Low | Low | Low | Moderate | Low | Low | Low | High | Low | Low | Low | Low |
| 301 | John S, 2020 | Low | Low | Low | Low | Low | Low | Moderate | Low | High | Low | High | Low | Low |
| 302 | Kacem I, 2020 | Low | Low | Low | High | High | Low | High | High | High | High | High | Low | Moderate |
| 303 | Keller E, 2020 | Low | Moderate | Low | Low | Moderate | Moderate | High | High | High | Low | High | Low | Moderate |
| 304 | Lang M, 2020 | Low | Low | Low | Moderate | Moderate | Low | Moderate | Low | High | Low | High | Low | Low |
| 305 | Lin E, 2020 | Low | Low | Low | Low | Low | Low | Moderate | Low | High | Low | High | Low | Low |
| 306 | Liotta EM, 2020 | Low | Low | Low | Low | Moderate | Low | Low | Low | Low | High | Low | Low | Low |
| 307 | Requena M, 2020 | Low | Low | Low | Low | Low | Low | Low | Low | Low | Low | Low | Low | Low |
| 308 | Salahuddin H, 2020 | Low | Low | Low | Low | Low | Low | High | Low | High | High | High | Low | Low |
| 309 | Shekhar R, 2020 | Low | Low | Low | Low | Low | Low | Low | Low | High | Low | Low | Low | Low |
| 310 | Siegler JE, 2020 | Low | Low | Low | Low | Low | Low | Moderate | Low | High | Low | High | Low | Low |
| 311 | Yao X, 2020 | Low | Low | Low | Low | Moderate | Low | Low | Low | High | Low | Low | Low | Low |
| 312 | Wang YX, 2020 | Moderate | High | High | Low | Low | Moderate | High | High | Low | High | High | High | High |
| 313 | O'Reilly GM, 2020b | Low | Low | Low | Low | Low | Low | Low | Low | Moderate | Low | Low | Low | Low |
| 314 | Bellan M, 2020 | Low | Low | Low | Low | Low | Low | Moderate | Low | Low | High | High | Low | Low |
| 315 | Chen SL, 2020 | Low | Low | Low | Low | Low | Low | Low | Low | Low | Low | Low | Low | Low |
| 316 | Horvath L, 2020 | Low | Low | Low | Low | High | Low | Moderate | Low | Moderate | High | Low | Low | Low |
| 317 | Huang H, 2020 | Low | Low | Low | Low | High | Low | Moderate | Low | Low | High | High | Low | Low |
| 318 | Rokni M, 2020 | Low | Low | Low | Low | Moderate | Low | Low | Low | Low | Low | High | Low | Low |
| 319 | Shah NN, 2020 | Low | Low | Low | Low | Moderate | Low | Moderate | Low | Low | High | High | Low | Low |
| 320 | Dang JZ, 2020 | Low | Low | Low | Low | Low | Low | Low | Low | Low | Low | Moderate | Low | Low |
| 321 | Knopp P, 2020 | Low | Low | Low | Low | Low | Low | High | High | High | High | Moderate | Low | Low |
| 322 | Li P, 2020 | Low | Low | Low | Low | Low | Low | Low | Low | Low | High | Low | Low | Low |
| 323 | BaĞ SoytaŞ R, 2020 | Low | Low | Low | Low | High | Low | Low | Low | Low | Low | High | Low | Low |
| 324 | Davis P, 2020 | Low | Low | Low | Low | High | Low | Low | Low | Low | High | Low | Low | Low |
| 325 | Gan JM, 2020 | Low | Low | Low | Low | Low | Low | Moderate | Low | Low | High | High | Low | Low |
| 326 | Hwang J, 2020 | Low | Low | Low | Low | High | Low | Low | Low | Low | Low | Low | Low | Low |
| 327 | Kennedy M, 2020 | Low | Low | Low | Low | Moderate | Low | Moderate | Low | High | High | Low | Low | Low |
| 328 | Mendes A, 2020 | Low | Low | Low | Low | Low | Low | Low | Low | Low | Low | Low | Low | Low |
| 329 | Vrillon A, 2020 | Low | Low | Low | Low | Low | Low | Low | Low | Low | Low | Low | Low | Low |
| 330 | Zhou J, 2020 | Low | Low | Low | Low | High | Low | Low | Low | Low | High | Low | Low | Low |
| 331 | Wang L, 2020b | High | High | Low | Low | Low | Low | Low | Low | Low | Low | Low | Low | Moderate |
| 332 | Fakiri K El, 2020 | Low | Low | Low | Low | Low | Low | Moderate | Low | Low | High | Moderate | Low | Low |
| 333 | Gotzinger F, 2020 | Low | Low | Low | Low | Low | Low | High | Low | High | Moderate | High | Low | Low |
| 334 | Guo CX, 2020 | Low | Low | Moderate | Low | Low | Low | Moderate | Low | Low | High | Moderate | Low | Low |
| 335 | Korkmaz MF, 2020 | Low | Low | Low | Low | Low | Low | Moderate | Low | Moderate | Low | Moderate | Low | Low |
| 336 | Mamishi S, 2020 | Low | Low | Low | Low | Low | Low | Moderate | High | High | Low | Low | Low | Low |
| 337 | Qui H, 2020 | Low | Low | Low | Low | Low | Low | Low | High | Low | Low | Low | Low | Low |
| 338 | Romani L, 2020 | Moderate | Low | Low | Low | Low | Low | Moderate | Low | High | Low | Moderate | Low | Low |
| 339 | Song X, 2020 | Low | Low | Moderate | Low | Low | Low | High | Low | High | High | Moderate | Low | Low |
| 340 | Sun D, 2020 | Low | Low | Low | Low | Low | Moderate | Moderate | High | High | Low | Low | Moderate | Low |
| 341 | Swann OV, 2020 | Low | Low | Low | Low | Low | Low | Moderate | Low | High | High | Low | Low | Low |
| 342 | Zhao Y, 2020 | Low | Low | Low | Low | Low | Low | High | High | High | High | Moderate | Low | Low |
| 343 | Bayesheva D, 2020 | Low | Low | Low | Low | Moderate | Low | Low | Low | Low | Low | High | Low | Low |
| 344 | Bhumbra S, 2020 | Low | Low | Low | Low | Low | Low | Low | Low | Low | Low | Low | Low | Low |
| 345 | Cai J, 2020 | Low | Low | Low | Low | Low | Low | Moderate | High | Low | Low | High | Low | Low |
| 346 | Chen J, 2020 | Low | Low | Low | Low | Low | Low | Low | Low | Low | Low | High | Low | Low |
| 347 | Duramaz BB, 2020 | Low | Moderate | Moderate | Low | Low | Moderate | Low | Low | Low | Low | High | Low | Low |
| 348 | Li K, 2020 | Low | Low | Low | Low | Moderate | Low | Moderate | High | Low | Low | High | Low | Low |
| 349 | Parri N, 2020 | Low | Low | Low | Low | Low | Low | Low | Low | Low | Low | High | Low | Low |
| 350 | Yilmaz K, 2020 | Low | Low | Low | Low | Moderate | Low | Low | Low | Low | Low | High | Low | Low |

**Key:** Point 1- Clear Inclusion criteria in case series; Point 2- Condition measured in a standard, reliable way; Point 3- Valid methods used for identification of the condition; Point 4- Consecutive inclusion of participants; Point 5- Complete inclusion of participants; Point 6- Clear reporting of demographics; Point 7- Clear reporting of clinical information; Point 7.1- Comorbidities; Point 7.2- Stage of disease; Point 7.3- Imaging Information; Point 7.4- Entry point of patient in study; Point 8- Clear reporting of presenting site(s)/clinic(s) demographic information.

**Table 10:** Neurological symptoms & signs in the form of case reports and case series (excluded from systematic review and meta-analysis)

| **S. No** | **Neurological symptoms & signs** | **Number of case series (>1 case)** | **Number of case reports** | **Number of cases** | **References** |
| --- | --- | --- | --- | --- | --- |
| 1. | Headache | 46 | 3 | 2224 | ^1–28^ ^29–48^ |
| 2. | Fatigue | 41 | 0 | 2036 | ^1,2,5,6,9,13–16,19–22,49–59^ ^31–36,40–43,60–66^ |
| 3. | Smell or Taste Impairment | 3 | 0 | 1082 | ^9,13,67^ |
| 4. | Myalgia | 43 | 4 | 864 | ^1,9,10,14–24,26,27,49,52,54,55,57,68–73^ ^31–34,36,38–42,47,60,62–65,74,75^ |
| 5. | Smell Impairment | 25 | 8 | 856 | ^4,7,9,10,14,16,18–20,23–27,53,67,76–87^ ^34,38,39,44,46^ |
| 6. | Disturbance of Consciousness | 6 | 4 | 232 | ^4,25,27,72,88–90^ ^44,46,47^ |
| 7. | Taste Impairment | 14 | 3 | 201 | ^4,7,9,14,18–20,24,26,27,78,80,81,84,85^ ^39,46^ |
| 8. | Dizziness | 14 | 0 | 155 | ^2,3,8,15,16,21,22,51,54,57^ ^31,46,47,91^ |
| 9. | Seizure | 15 | 7 | 91 | ^4,7,11,25,87–89,92–100^ ^44–47,101,102^ |
| 10. | Acute Confusion/ Delirium | 9 | 1 | 83 | ^4,10,12,19,27,103–105^ ^43,106^ |
| 11. | Myalgia or Fatigue | 1 |  | 75 | ^48^ |
| 12. | Focal neurological deficits | 1 | 0 | 34 | ^25^ |
| 13. | Hemiplegia | 2 | 1 | 30 | ^107 44,46^ |
| 14. | Cranial Nerve Palsy | 7 | 5 | 29 | ^7,92,108–111^ ^112–115^ |
| 15. | Corticospinal tract signs | 2 | 2 | 16 | ^4,45,72,116^ |
| 16. | Myoclonus | 2 | 1 | 14 | ^12,117–120^ |
| 17. | Short term memory loss | 1 | 0 | 12 | ^7^ |
| 18. | Ataxia | 4 | 2 | 10 | ^7,118^ ^46,106,120^ |
| 19. | Agitation | 1 | 0 | 9 | ^4^ |
| 20. | Pseudo Bulbar Palsy | 1 | 1 | 9 | ^88,115^ |
| 21. | Behavioral abnormalities | 1 | 0 | 4 | ^25^ |
| 22. | Language impairment | 1 | 0 | 4 | ^106^ |
| 23. | Parasthesia | 1 | 0 | 1 | ^7^ |

**Table 11:** Neurological diagnosis in the form of case reports and case series (excluded from systematic review and meta-analysis)

| **S. No** | **Neurological diagnosis** | **Number of case series (>1 case)** | **Number of case reports** | **Number of cases** | **References** |
| --- | --- | --- | --- | --- | --- |
| 1. | Total stroke (TIA, IS, ICH, SAH & CVST) | 54 | 30 | 1090 | ^7,25,92,95,121–152,153–166 44,46,135,167–196^ |
| 2. | Encephalopathy  (Including ANE and Leukoencephalopathy) | 16 | 8 | 250 | ^7,92,95,197,198,11,153,199–206 45,46,106,207–212^ |
| 3. | GBS (AIDP, AMAN, AMSAN) | 13 | 32 | 106 | ^25,92,95,213–232,11,233–238 44–46,239–250^ |
| 4. | Meningoencephalitis | 4 | 3 | 34 | ^4,44,251–255^ |
|  | Encephalitis (Including LE, AHNE, BE,  Para Infectious AIE,  Rhombencephalitis) | 5 | 16 | 71 | ^92,95,256–268 46,269–273^ |
| 5. | Neuropsychiatric disorders  - Associated with NMDAR Encephalitis | 3 | 0 | 32 | ^92,274,275^ |
|  |  | 0 | 1 | 1 | ^276^ |
| 6. | Posterior reversible encephalopathy syndrome (PRES) | 8 | 2 | 18 | ^7,25,151,162,163,277,278 279–281^ |
| 7. | CIM | 3 | 1 | 16 | ^282,283 47,275^ |
|  | CIN | 3 | 0 | 13 | ^11 46,275^ |
|  | CINM (Critical Illness neuro-myopathy) | 1 | 0 | 4 | ^284^ |
| 8. | ADEM (Including AHL & AHLE) | 1 | 4 | 13 | ^95,285,286 287,288^ |
| 9. | Muscle Injury & Rhabdomyolysis | 2 | 3 | 12 | ^7,289,290 47,291^ |
| 10. | Acute Myelitis  (Including ATM, ANM, Post/Para Infectious Myelitis) | 1 | 7 | 9 | ^85,95,227,292–294 248,295^ |
| 11. | Cytotoxic lesion of the Corpus Callosum (CLOCCs) | 3 | 2 | 8 | ^25,163,296–298^ |
| 12. | Meningitis | 2 | 1 | 8 | ^11,46,84^ |
| 13. | Movement disorder | 1 |  | 8 | ^46^ |
| 14. | Dysautonomia | 1 | 1 | 7 | ^7,225^ |
| 15. | GBS-MFS overlap &  MFS | 3 | 1 | 5 | ^231,232,299,300^ |
| 16. | Epilepsy | 1 | 1 | 5 | ^121,301^ |
|  | NCSE | 3 | 3 | 6 | ^95,302,303 210,304,305^ |
|  | NORSE  -Associated with NMDAR Encephalitis | 1 | 0 | 1 | ^303^ |
|  |  | 0 | 1 | 1 | ^306^ |
|  | SE | 1 | 0 | 1 | ^210^ |
| 17. | Myositis/Inflammatory Myopathy | 1 | 2 | 4 | ^47,307,308^ |
| 18. | CNS microangiopathy/ Vasculitis | 1 | 2 | 3 | ^92,147,309^ |
| 19. | Brachial Plexopathy/ Brachial Neuritis | 2 | 1 | 3 | ^92,95,310^ |
| 20. | Opsoclonus-Myoclonus Syndrome (OMS) | 1 | 1 | 2 | ^92,311^ |
| 21. | Neuropathy | 1 | 1 | 2 | ^46,312^ |
| 22. | Myasthenia Gravis |  | 2 | 2 | ^313,314^ |
| 23. | CNS Abscess with ventriculitis | 1 | 0 | 1 | ^95^ |
| 24. | Acute Cerebellitis | 0 | 1 | 1 | ^315^ |
| 25. | Optic Neuritis | 1 | 0 | 1 | ^25^ |
| 26. | Vestibulo Cochlear Neuritis | 1 | 0 | 1 | ^25^ |
| 27. | Pseudo tumor cerebri | 1 | 0 | 1 | ^95^ |
| 28. | Myelopathy/ cord ischemia | 1 | 0 | 1 | ^95^ |
| 29. | Postural Tachycardia  Syndrome (POTS) | 0 | 1 | 1 | ^316^ |
| 30. | Acute Tumefactive demyelination | 1 | 0 | 1 | ^163^ |
|  | Demyelinating lesions of Brain & Spinal Cord  -Associated with Aquaporin Antibody  -Associated with MOG antibody | 0 | 2 | 2 | ^317,318^ |
|  |  | 0 | 1 | 1 | ^319^ |
|  |  | 0 | 2 | 2 | ^320,321^ |
| 31. | Multiple Sclerosis |  | 1 | 1 | ^322^ |
| 32. | Pansinusitis | 1 |  | 1 | ^184^ |
| 33. | Cerebellar Syndrome | 1 |  | 1 | ^45^ |

**Abbreviations:** ADEM- Acute disseminated encephalomyelitis, AHL- Acute Hemorrhagic Leukoencephalomyelitis, AHLE- Acute Hemorrhagic leukoencephalitis, AHNE- Acute Hemorrhagic, Necrotizing Encephalitis, AIE- Autoimmune encephalitis, AIDP- Acute inflammatory demyelinating polyneuropathy, AMAN- Acute motor axonal neuropathy, AMSAN- Acute motor sensory axonal neuropathy, ANM- Acute Necrotizing Myelitis, ANE-Acute Necrotizing Encephalopathy, ATM- Acute Transverse Myelitis, BE-Bickerstaff Encephalitis, CLOCC- Cytotoxic lesion of the Corpus Callosum, CIM-Critical Illness Myopathy, CIN- Critical illness Neuropathy, CINM- Critical Illness Neur-myopathy, CNS- Central Nervous System, CVST- Cerebral venous sinus thrombosis, GBS- Guillain Barre Syndrome, ICH- Intra Cerebral Hemorrhage, IS- Ischemic Stroke, LE-Limbic Encephalitis, MFS- Miller Fisher syndrome, MOG- Myelin Oligodendrocyte Glycoprotein, NCSE- Non-Convulsive Status Epilepticus, NORSE- New Status Onset Refractory Status Epilepticus, SE-Status Epilepticus, OMS- Opsoclonus-Myoclonus Syndrome, POTS- Postural Tachycardia Syndrome, PRES- Posterior reversible encephalopathy syndrome, SAH- Sub Arachnoid Hemorrhage, TIA- Transient Ischemic Attack.
